# Optimal tracheal tube rotation patterns for navigating through the glottis: an in-silico quantification

**DOI:** 10.1101/2023.08.22.23294387

**Authors:** Erich B Schulz, Lucy Carra Schulz

## Abstract

Seeking to unpack some of the anaesthetists’ “knack” for intubation, this study examines the effect of various orientations of the tracheal tube on the anterior movement of the tube tip using a computerised 3D model of intubation.

The model used sets of coordinates for the upper incisor tip, lower incisor tip and vallecula extracted from mean values reported in a study of 16 volunteers predicted to have easy laryngoscopy and 16 predicted to have difficult laryngoscopy during both gentle laryngoscopy and laryngoscopy under 50N of lifting force, yielding a total of four sets of airway geometry.

Tube orientation was specified with the standard aviation terms pitch, roll and yaw. Observations were repeated across permutations of tube roll (0° to 45°) and yaw (0° to 15°) in all four geometric configurations.

Across all four geometries, the most favourable tip location was observed with close to 15° of yaw and 0° roll with an anterior tip movement at the level of the glottis observed between 19.2 and 26.6mm. Unsurprisingly given the curved shapes of the objects involved, incremental movement of the tip was greatest at extreme values of roll and yaw.

Both yaw and roll caused posterolateral movement of the maxillary teeth contact point. The posterior motion at the mouth enables the entire tube to pitch tip up. However, rolling the tube caused the tube tip to move posteriorly and the pivot point on the laryngoscope blade to move cephalad, nearly always negating what should be a favourable change in pitch allowed by the posterolateral maxillary dentition contact point.

Our analysis suggests that avoiding tube roll while maximising yaw at the time of glottic entrance may be a previously unrecognised manoeuvre to improve tracheal intubation success rate having implications for intubation teaching and simulation. Understanding the importance of posterolateral movement of the tube at the oral cavity also may provide new insights into the cause of some difficult intubations.

## Introduction

Anaesthetists are universally adept at intubating, able to orient the tube in “just the right way” so that the tip arches anteriorly around the tongue and through the glottis. Unfortunately, outside of operating theatres, even with video laryngoscopy or a full view of the larynx, the first-attempt failure rate of tracheal intubation sits around 13%^1^, frequently blamed on a perceived “anterior larynx”^2^. Opportunities to develop this skill are rare^3^, but it has been estimated that it takes 50 attempts in order to achieve even basic proficiency^4^.

Better explanations to trainees may hasten learning, leading to lifesaving skill improvement in those called upon to intubate in an emergency. Seeking to unpack some of the anaesthetists’ “knack”, this study measures the effect of various orientations of the tracheal tube on the anterior movement of the tube tip.

## Method

We developed a computerised 3D model of intubation (https://bitbucket.org/erichbschulz/babairway), including a tracheal tube, maxillary dentition and a laryngoscope blade (Figure 1) using the Babylon JS library (https://www.babylonjs.com/).

**Figure 1.**
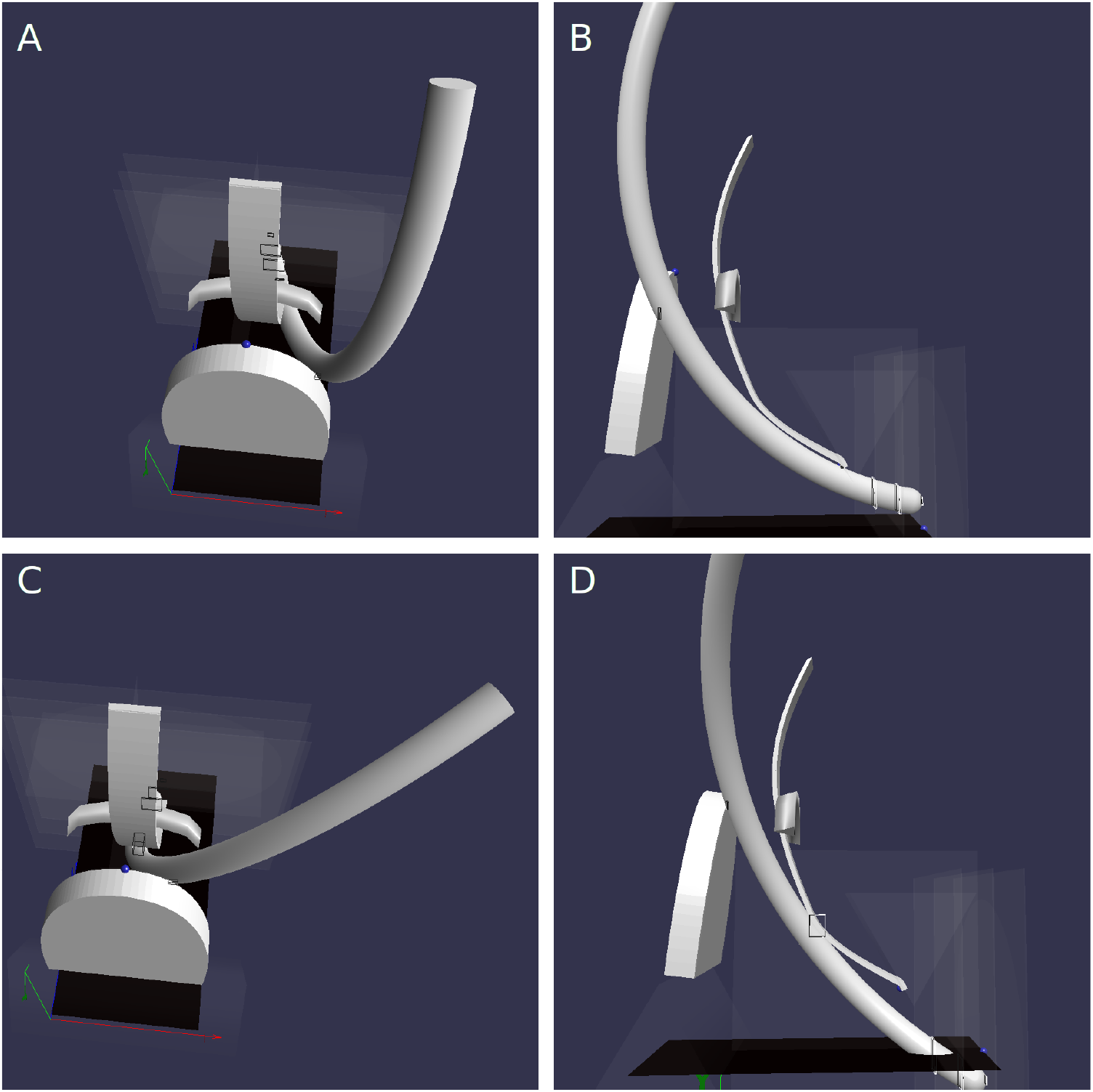
Renders of the simulated upper airway during intubation showing the impact of 15° of yaw (A and B) compared to 45° of roll (C and D). Geometry derived from mean vallecula locations in 16 patients with anticipated easy intubation during a 50N laryngoscopy. Shown are the maxillary dental arch, front mandibular teeth, laryngoscope blade and tracheal tube, from the perspective of intubation (A and C) and from the right side (B and C). Panels B and D demonstrate the similar appearance of yaw and roll when viewed from the side despite the dramatic impact on tube tip location at the level of the glottis.

**Figure 2.**
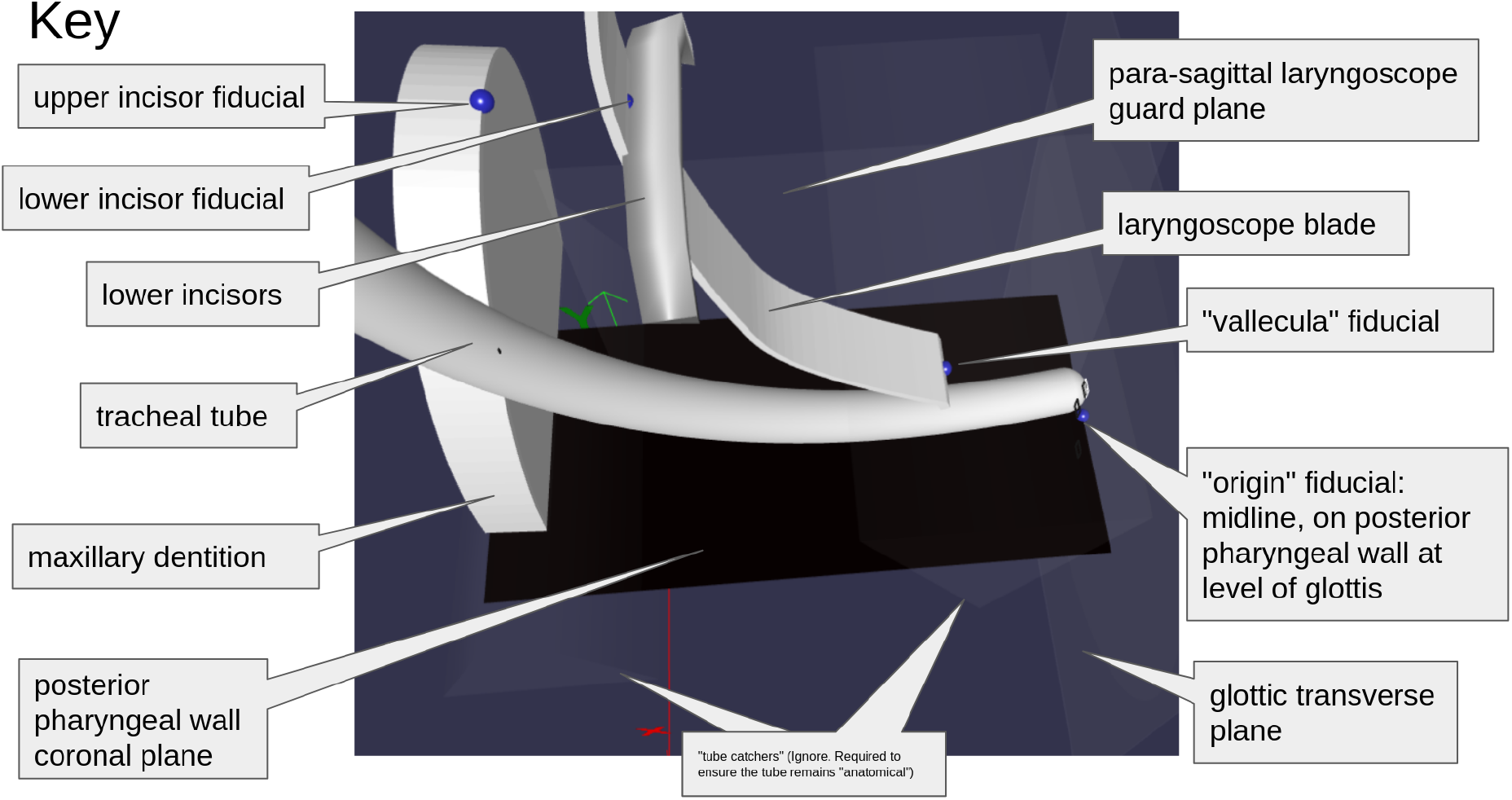
Labelled render.

Sets of coordinates for the upper incisor tip, lower incisor tip and vallecula were extracted from mean values reported in a study of 16 volunteers with predicted easy laryngoscopy and 16 with predicted difficult laryngoscopy^5^. This study used lateral photographs taken during gentle laryngoscopy and laryngoscopy under 50N of lifting force, yielding a total of four sets of airway geometry.

Lower incisor position was derived by applying the mouth opening vector calculated by relative movements of the mental process to upper incisor. The distance between the transverse plane of the glottis and vallecula was taken from a computerised tomography study of 54 healthy volunteers^6^. The maxillary dentition was placed with an approximate 7 degrees occlusal angle relative to the perpendicular from the facial plane defined in the reference study^5^.

The simulated tracheal tube had a curve radius of 140mm and external diameter of 10mm. A simulated size 4 Macintosh blade was placed so that the tip sits in the vallecula, abutting the lower incisors.

The position of the tube is specified with the standard aviation terms pitch, roll and yaw describing rotation around the lateral, longitudinal and vertical axes respectively. For a given roll and yaw angle the software precisely positioned the tube against the maxillary dentition, the anterior blade surface, the parasagittal guard surface of the laryngoscope blade and the transverse plane at the level of the glottis. This position was found by making a series of translational moves and pitch angle adjustments of decreasing size directed by detection of 3D overlaps between the mesh structure representing the tube, blade and dentition.

These observations were repeated across permutations of tube roll (0° to 45°) and yaw (0° to 15°) in all four geometric configurations. With each permutation the point of contact between the tube and its constraints was recorded, together with a set of five automated screenshots. This information is available in the supplemental materials.

## Results

Visual inspection of the 3D renders illustrates the subtle visual difference of tube yawing versus rolling when viewed from the side (Figure 1.B and 1.D).

In all four geometries, the most favourable tip location was observed with close to 15° of yaw and 0° roll (Table 1). Across the four geometries, moving from 45° of roll to 15° yaw caused an anterior tip movement of between 19.2 and 26.6mm. Overall, each 15° of roll led to a mean 1.8mm (range -0.8 to 5.6mm) posterior tip movement, whereas each 5° of yaw caused a mean 5.5mm (range -1.2 to 11.0mm) anterior tip movement. Unsurprisingly given the curved shapes of the objects involved, incremental movement of the tip was greatest at extreme values of roll and yaw.

**Table 1:**
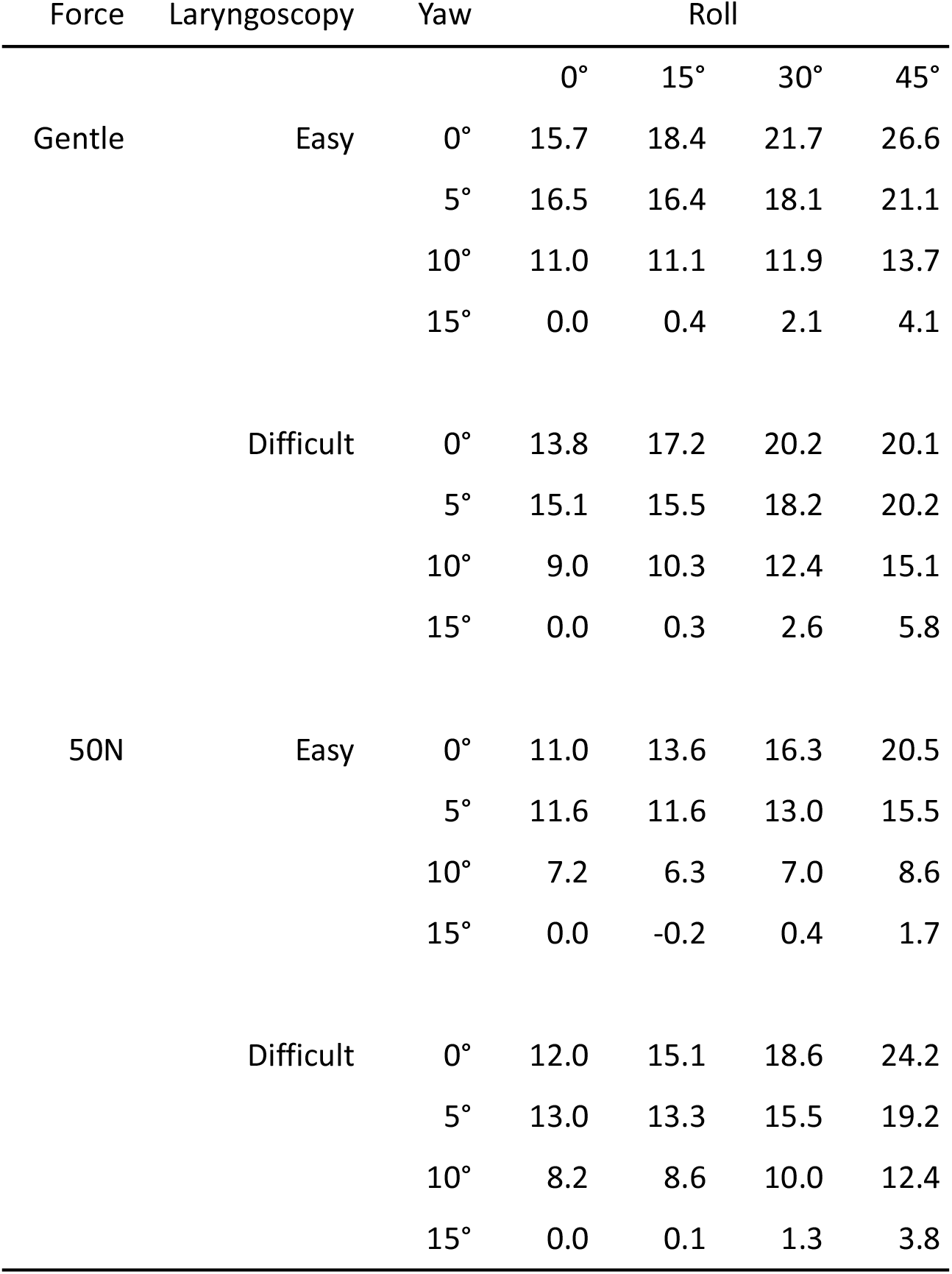
Observed posterior displacement (mm) of the tracheal tube tip at the level of the glottis during tube roll and yaw, during gentle and vigorous (50N) laryngoscopy in intubation geometry from patients with anticipated easy and difficult intubation. Positions are relative to the 15° yaw and 0° roll orientation.

Both yaw and roll caused posterolateral movement of the maxillary teeth contact point. The posterior motion at the mouth enables the entire tube to pitch tip up.

However, rolling the tube caused the tube tip to move posteriorly, nearly always negating what should be a favourable change in pitch allowed by the posterolateral maxillary dentition contact point.

## Conclusion

Our analysis suggests that avoiding tube roll while maximising yaw at the time of glottic entrance may be a previously unrecognised manoeuvre to improve intubation success rate. This has implications for intubation teaching and simulation. Understanding the importance of posterolateral movement of the tube at the oral cavity also may provide new insights into the cause of some difficult intubations.

Our simulation is limited by being based on geometry observed in a population of only 32 patients, presumably drawn from a single ethnicity. Future work should clarify normal and pathological soft tissue limitations on extreme tube yaw, expand the simulation to use geometry from other populations and validate our findings in a clinical environment.

## Supporting information

Abbreviated Renders

Full Renders

## Data Availability

All data screen renders are included the supplemental material. The software used is available here:
https://bitbucket.org/erichbschulz/babairway/src/main/

https://bitbucket.org/erichbschulz/babairway

## Acknowledgements

No competing interests declared. We wish to acknowledge valuable peer-support and advice from anaesthetists at Mater Hospital in Brisbane; Dr Robert Read, Public Invention, Austin, Texas; and Leanne Petty for her assistance modelling the maxillary arch and laryngoscope.

## Supplementary Material

**Table 2:** Glottic plane movement sensitivity analysis

**Table 2:**
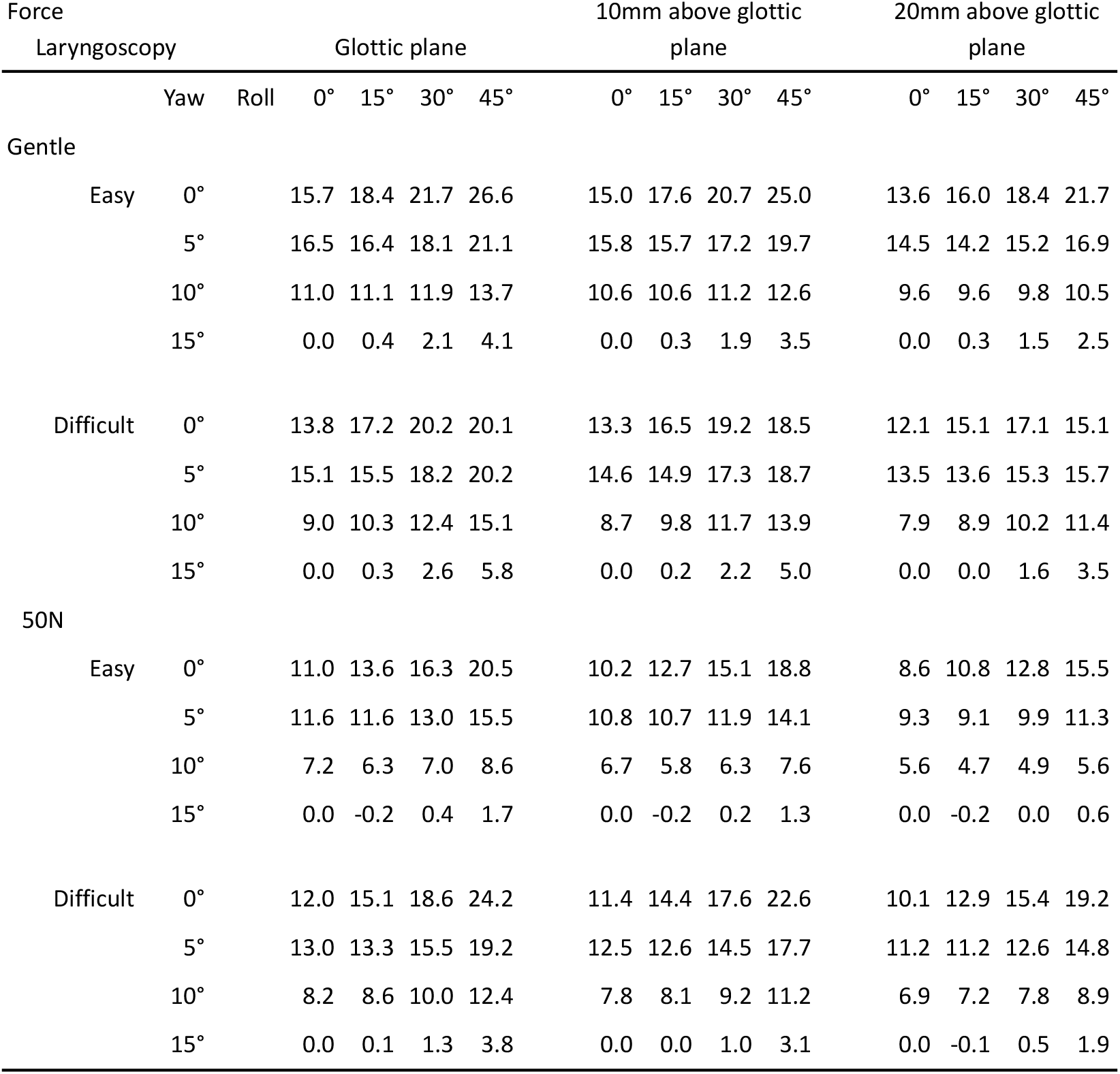
Posterior displacement (mm) of the tracheal tube tip at various transverse planes, expanding on the findings in Table 1 and illustrated in Figure 1. This supplemental analysis documents the sensitivity of the conclusions to changes in the distance from the vallecula to the transverse plane at the level of the glottis. This analysis confirms that while the magnitude of the observed movements decreases, the overall pattern is unchanged.

**Raw Observations** suitable for importing into spreadsheet or other analysis software

**Abbreviated renders:** a small file showing a selection of renders

**Full renders:** complete set of observations for all sixteen orientation permutations, all four geometries in five views

### Raw Observations

This is the settings and data collected by model software during each scenario.

(This data is presented in a tiny font to prevent PDF conversion inserting line breaks. Please cut and paste without formatting or resize after pasting into your spreadsheet or analysis software.)

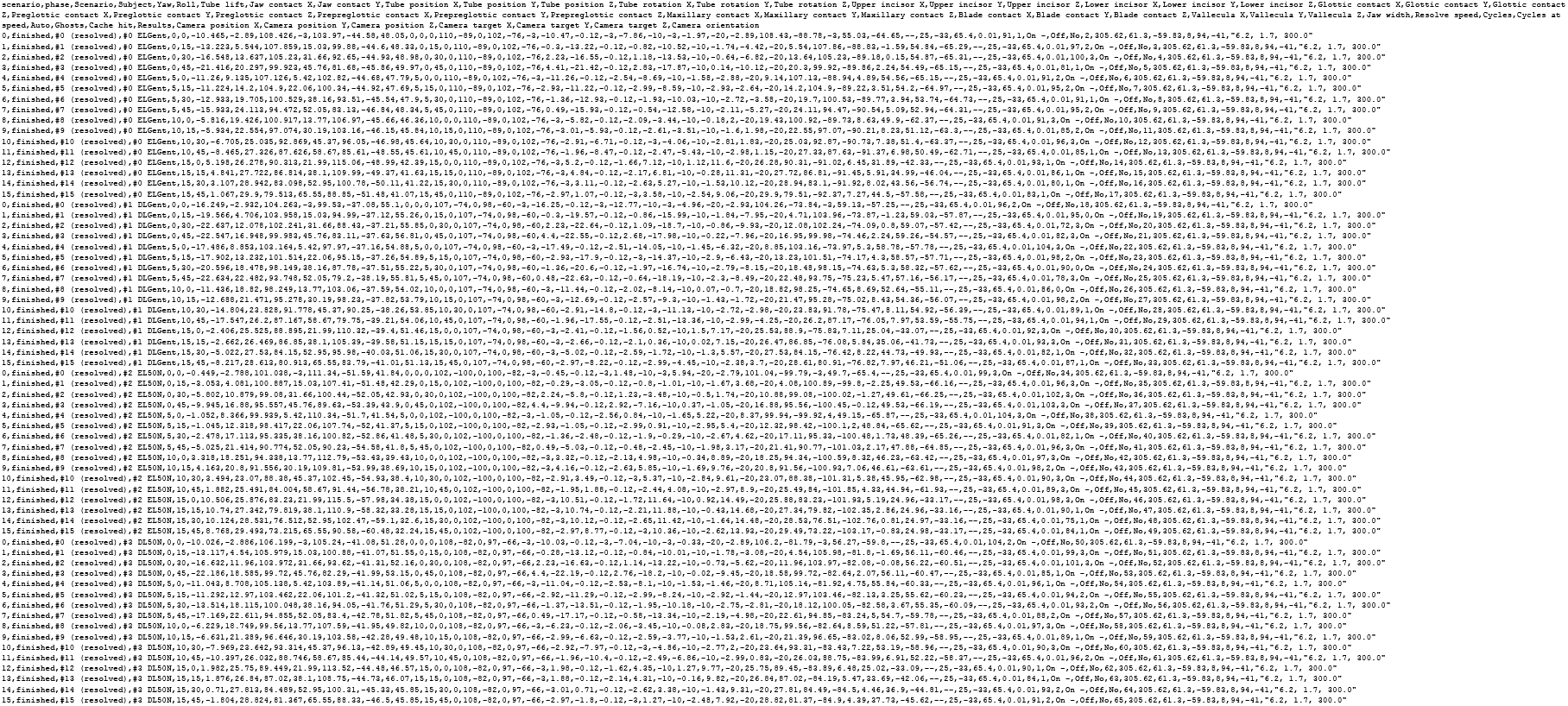

