## Supplementary material for "Optimal tracheal tube rotation patterns for navigating through the glottis: an in-silico quantification": Abbreviated Renders

### Model

### Gallery

#### ELGent Yaw: 0.0° Roll: 0.0°

```
Scenario: #0 (resolved)
Subject: #0 ELGent
Yaw: 0.0°
Roll: 0.0°
Tube lift: -11.986 mm
Jaw contact X: -2.905 mm
Jaw contact Y: 108.262 mm
Tube position: (-3.00, 102.46, -44.57)
Tube rotation: (48.07, 0.00, 0.00)
Upper incisor: (0.00, 110.00, -89.00)
Lower incisor: (0.00, 102.00, -76.00)
Glottic contact: (-3.00, -11.99, -0.12)
Preglottic contact: (-3.00, -9.38, -10.00)
Prepreglottic contact: (-3.00, -3.49, -20.00)
Maxillary contact: (-2.90, 108.26, -88.78)
Blade contact: (-3.00, 54.76, -65.25)
```

##### intubation view

##### side view

##### top view

##### above view

##### glottis view

Vallecula: (--, 25.00, -33.00)
Jaw width: 65.4mm
Resolve speed: 0.01
Cycles: 91
Cycles at speed: 1
Auto: On
Ghosts: Off
Cache hit: No
Results: 0
Camera position: (414.71, 49.32, -66.73)
Camera target: (8.00, 94.00, -41.00)
Camera orientation: 6.2, 1.7, 410.0

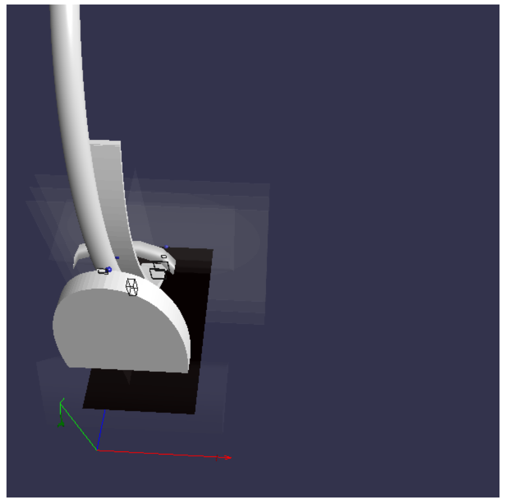

Camera position: (72.19, 330.13, -225.53)
Camera target: (60.00, 0.00, 0.00)
Camera orientation: -7.8, 0.6, 400.0

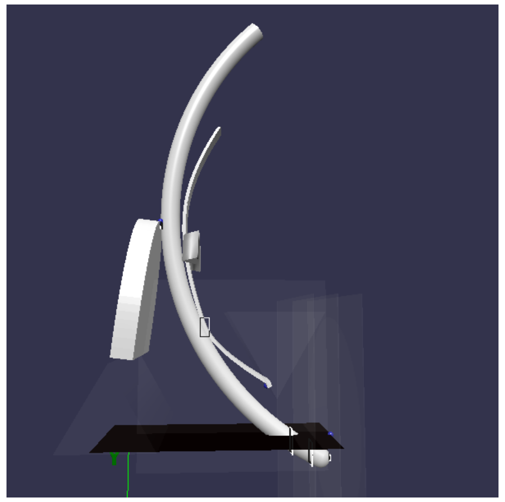

Camera position: (305.62, 61.30, -59.83)
Camera target: (8.00, 94.00, -41.00)
Camera orientation: 6.2, 1.7, 300.0

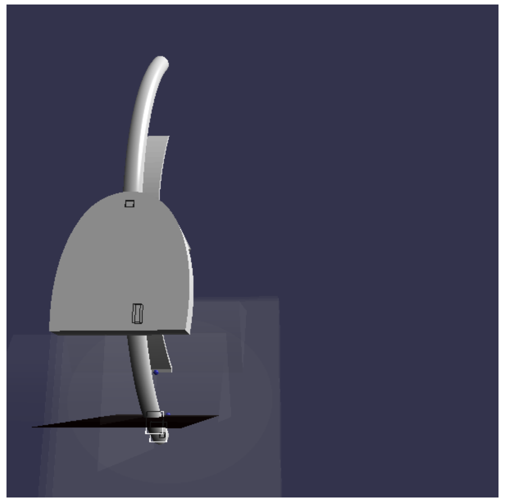

Camera position: (84.94, 16.85, -367.75)
Camera target: (50.00, 110.00, 30.00)
Camera orientation: 4.8, 1.8, 410.0

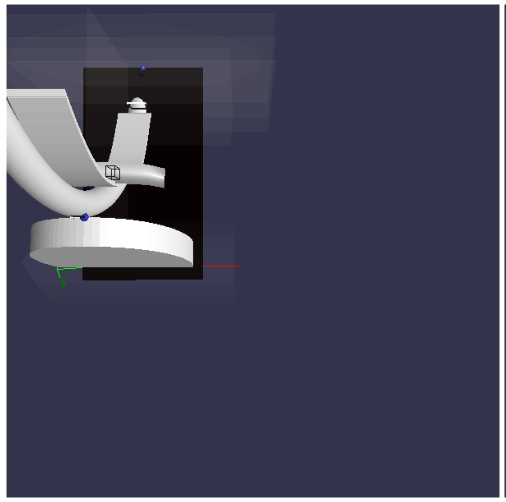

Camera position: (59.99, 316.98, -103.17)
Camera target: (60.00, 0.00, -100.00)
Camera orientation: 4.7, 0.0, 317.0

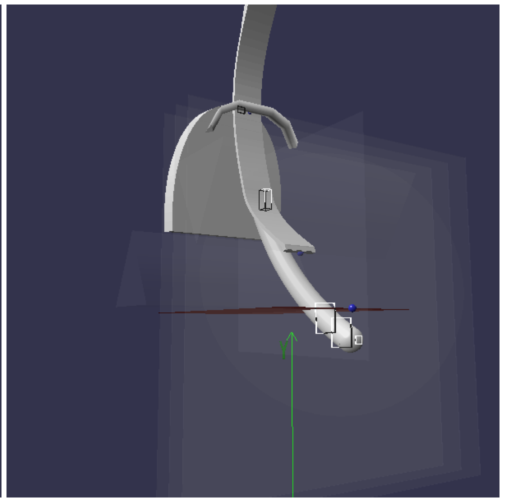

Camera position: (149.73, -6.98, 170.04)
Camera target: (0.00, 30.00, -72.00)
Camera orientation: 7.3, 1.7, 287.0

#### ELGent Yaw: 0.0° Roll: 45.0°

Scenario: #1 (resolved)
Subject: #0 ELGent
Yaw: 0.0°
Roll: 45.0°
Tube lift: -21.416 mm
Jaw contact X: 20.297 mm
Jaw contact Y: 99.923 mm
Tube position: (45.76, 81.68, -45.86)
Tube rotation: (49.97, 0.00, 45.00)
Upper incisor: (0.00, 110.00, -89.00)
Lower incisor: (0.00, 102.00, -76.00)
Glottic contact: (4.41, -21.42, -0.12)
Preglottic contact: (2.83, -17.87, -10.00)
Prepreglottic contact: (0.14, -10.12, -20.00)
Maxillary contact: (20.30, 99.92, -89.86)
Blade contact: (2.24, 54.49, -65.15)
Vallecula: (--, 25.00, -33.00)
Jaw width: 65.4mm
Resolve speed: 0.01
Cycles: 81
Cycles at speed: 1
Auto: On
Ghosts: Off
Cache hit: No
Results: 1
Camera position: (149.73, -6.98, 170.04)
Camera target: (0.00, 30.00, -72.00)
Camera orientation: 7.3, 1.7, 287.0

##### intubation view

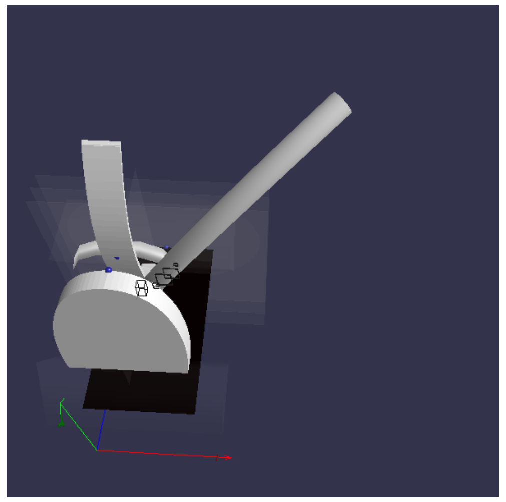

Camera position: (72.19, 330.13, -225.53)
Camera target: (60.00, 0.00, 0.00)
Camera orientation: -7.8, 0.6, 400.0

##### side view

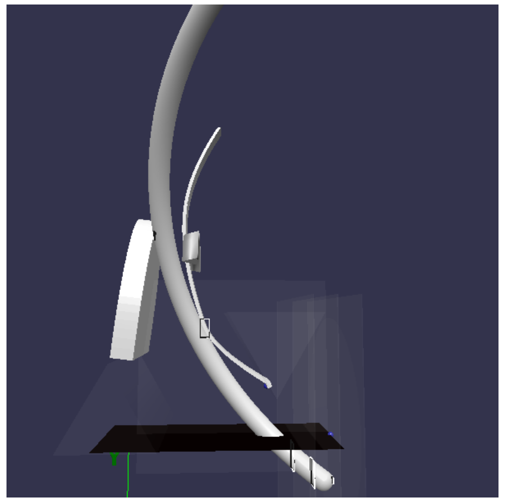

Camera position: (305.62, 61.30, -59.83)
Camera target: (8.00, 94.00, -41.00)
Camera orientation: 6.2, 1.7, 300.0

##### top view

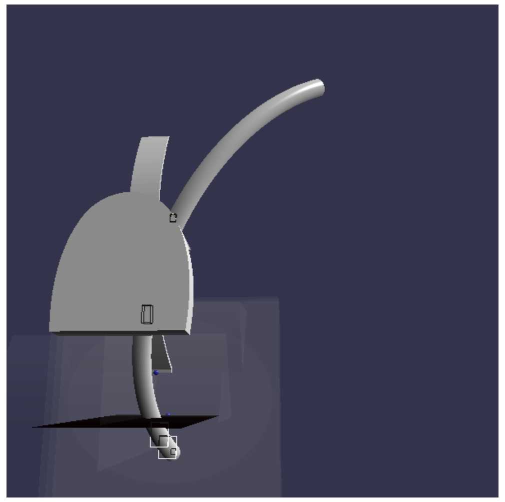

Camera position: (84.94, 16.85, -367.75)
Camera target: (50.00, 110.00, 30.00)
Camera orientation: 4.8, 1.8, 410.0

##### above view

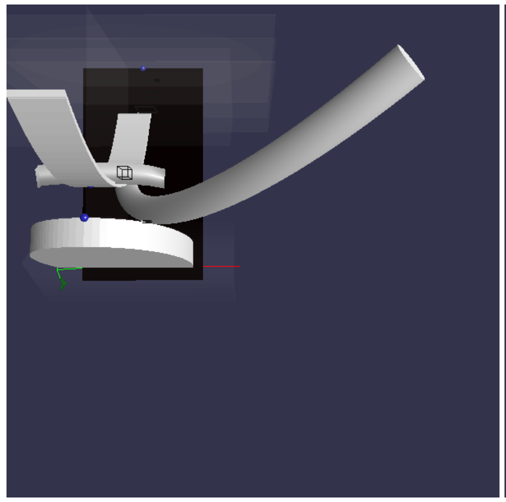

Camera position: (59.99, 316.98, -103.17)
Camera target: (60.00, 0.00, -100.00)
Camera orientation: 4.7, 0.0, 317.0

##### glottis view

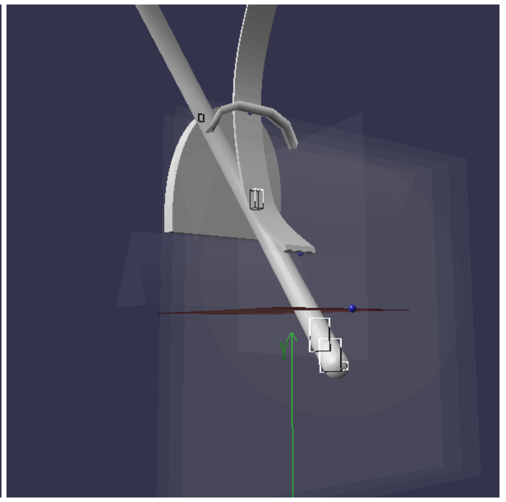

Camera position: (149.73, -6.98, 170.04)
Camera target: (0.00, 30.00, -72.00)
Camera orientation: 7.3, 1.7, 287.0

#### ELGent Yaw: 15.0° Roll: 0.0°

Scenario: #2 (resolved)
Subject: #0 ELGent
Yaw: 15.0°
Roll: 0.0°
Tube lift: 5.198 mm
Jaw contact X: 26.278 mm
Jaw contact Y: 90.313 mm
Tube position: (21.99, 115.06, -48.99)
Tube rotation: (42.39, 15.00, 0.00)
Upper incisor: (0.00, 110.00, -89.00)
Lower incisor: (0.00, 102.00, -76.00)
Glottic contact: (-3.00, 5.20, -0.12)
Preglottic contact: (-1.66, 7.12, -10.00)
Prepreglottic contact: (1.12, 11.60, -20.00)
Maxillary contact: (26.28, 90.31, -91.02)
Blade contact: (6.45, 31.89, -42.33)
Vallecula: (--, 25.00, -33.00)
Jaw width: 65.4mm
Resolve speed: 0.01
Cycles: 93
Cycles at speed: 1
Auto: On
Ghosts: Off
Cache hit: No
Results: 2

##### intubation view

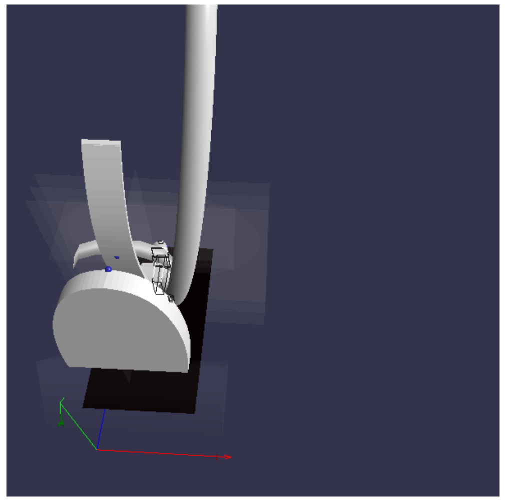

Camera position: (72.19, 330.13, -225.53)
Camera target: (60.00, 0.00, 0.00)

##### side view

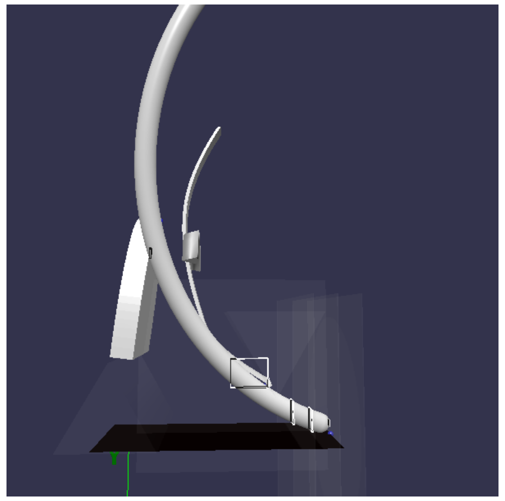

Camera position: (305.62, 61.30, -59.83)
Camera target: (8.00, 94.00, -41.00)

##### top view

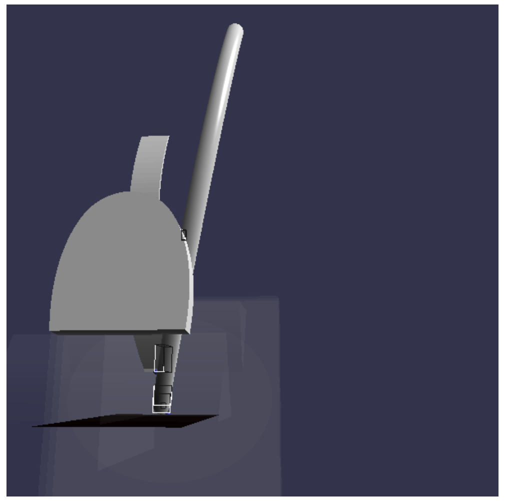

Camera position: (84.94, 16.85, -367.75)
Camera target: (50.00, 110.00, 30.00)

##### above view

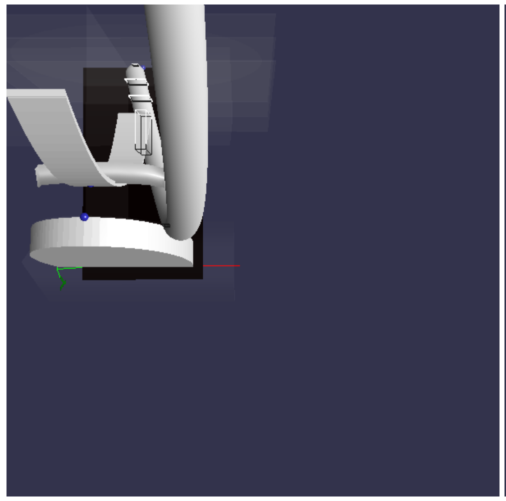

Camera position: (59.99, 316.98, -103.17)
Camera target: (60.00, 0.00, -100.00)

##### glottis view

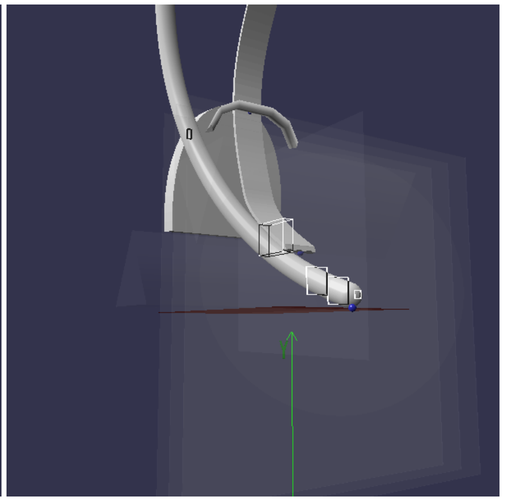

Camera position: (149.73, -6.98, 170.04)
Camera target: (0.00, 30.00, -72.00)

Camera position: (149.73, -6.98, 170.04)
Camera target: (0.00, 30.00, -72.00)
Camera orientation: 7.3, 1.7, 287.0

Camera orientation: -7.8, 0.6, 400.0

Camera orientation: 6.2, 1.7, 300.0

Camera orientation: 4.8, 1.8, 410.0

Camera orientation: 4.7, 0.0, 317.0

Camera orientation: 7.3, 1.7, 287.0

#### ELGent Yaw: 15.0° Roll: 45.0°

Scenario: #3 (resolved)
Subject: #0 ELGent
Yaw: 15.0°
Roll: 45.0°
Tube lift: 1.067 mm
Jaw contact X: 29.9 mm
Jaw contact Y: 79.513 mm
Tube position: (65.55, 88.85, -51.48)
Tube rotation: (41.07, 15.00, 45.00)
Upper incisor: (0.00, 110.00, -89.00)
Lower incisor: (0.00, 102.00, -76.00)
Glottic contact: (-2.97, 1.07, -0.12)
Preglottic contact: (-3.00, 3.58, -10.00)
Prepreglottic contact: (-2.54, 9.06, -20.00)
Maxillary contact: (29.90, 79.51, -92.37)
Blade contact: (7.27, 44.50, -57.58)
Vallecula: (--, 25.00, -33.00)
Jaw width: 65.4mm
Resolve speed: 0.01
Cycles: 83
Cycles at speed: 1
Auto: On
Ghosts: Off
Cache hit: No
Results: 3
Camera position: (149.73, -6.98, 170.04)
Camera target: (0.00, 30.00, -72.00)
Camera orientation: 7.3, 1.7, 287.0

##### intubation view

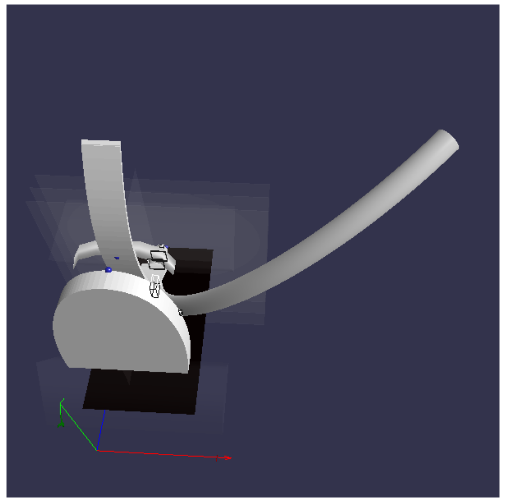

Camera position: (72.19, 330.13, -225.53)
Camera target: (60.00, 0.00, 0.00)
Camera orientation: -7.8, 0.6, 400.0

##### side view

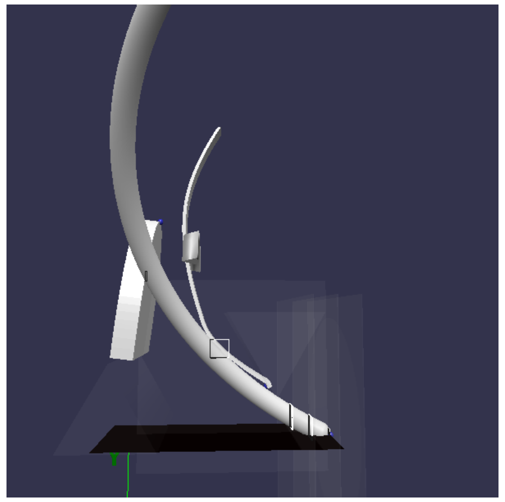

Camera position: (305.62, 61.30, -59.83)
Camera target: (8.00, 94.00, -41.00)
Camera orientation: 6.2, 1.7, 300.0

##### top view

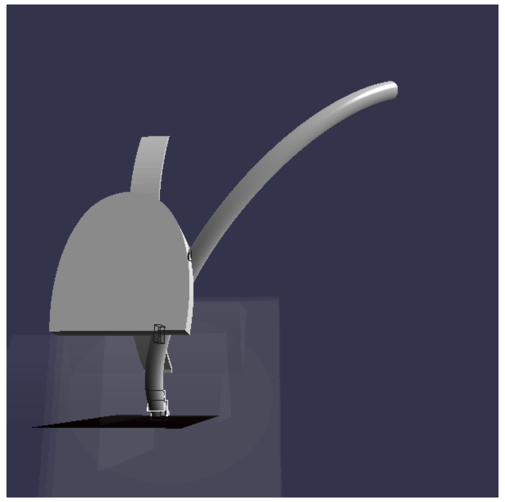

Camera position: (84.94, 16.85, -367.75)
Camera target: (50.00, 110.00, 30.00)
Camera orientation: 4.8, 1.8, 410.0

##### above view

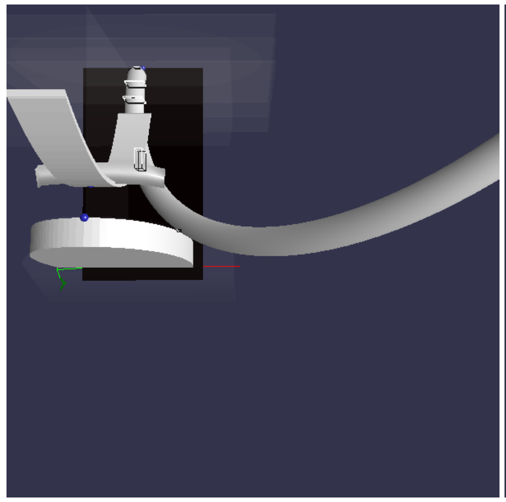

Camera position: (59.99, 316.98, -103.17)
Camera target: (60.00, 0.00, -100.00)
Camera orientation: 4.7, 0.0, 317.0

##### glottis view

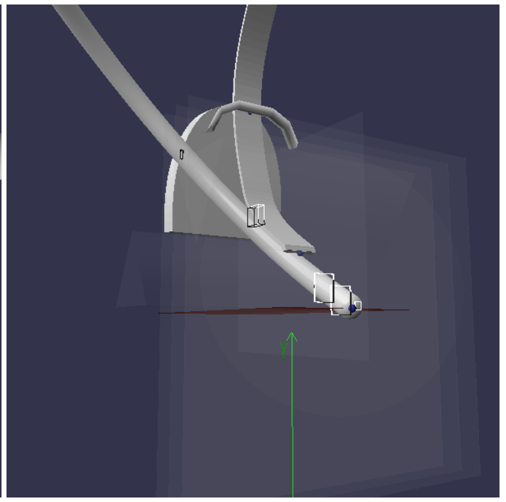

Camera position: (149.73, -6.98, 170.04)
Camera target: (0.00, 30.00, -72.00)
Camera orientation: 7.3, 1.7, 287.0

#### DLGent Yaw: 0.0° Roll: 0.0°

Scenario: #0 (resolved)
Subject: #1 DLGent
Yaw: 0.0°
Roll: 0.0°
Tube lift: -16.249 mm
Jaw contact X: -2.932 mm
Jaw contact Y: 104.263 mm
Tube position: (-3.00, 99.53, -37.08)
Tube rotation: (55.10, 0.00, 0.00)
Upper incisor: (0.00, 107.00, -74.00)
Lower incisor: (0.00, 98.00, -60.00)
Glottic contact: (-3.00, -16.25, -0.12)
Preglottic contact: (-3.00, -12.77, -10.00)
Prepreglottic contact: (-3.00, -4.96, -20.00)
Maxillary contact: (-2.93, 104.26, -73.84)
Blade contact: (-3.00, 59.13, -57.25)
Vallecula: (--, 25.00, -33.00)
Jaw width: 65.4mm
Resolve speed: 0.01
Cycles: 96
Cycles at speed: 2
Auto: On
Ghosts: Off
Cache hit: No
Results: 4
Camera position: (149.73, -6.98, 170.04)
Camera target: (0.00, 30.00, -72.00)
Camera orientation: 7.3, 1.7, 287.0

##### intubation view

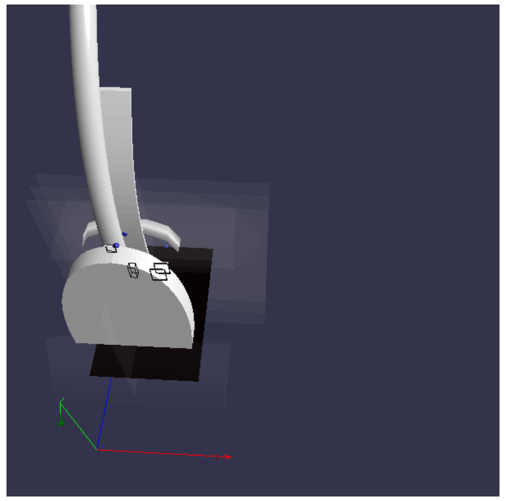

Camera position: (72.19, 330.13, -225.53)
Camera target: (60.00, 0.00, 0.00)
Camera orientation: -7.8, 0.6, 400.0

##### side view

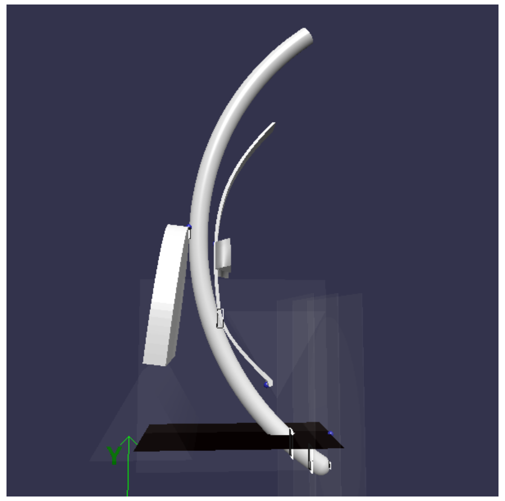

Camera position: (305.62, 61.30, -59.83)
Camera target: (8.00, 94.00, -41.00)
Camera orientation: 6.2, 1.7, 300.0

##### top view

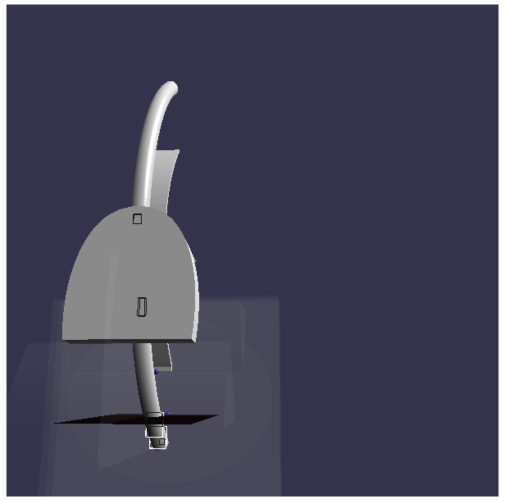

Camera position: (84.94, 16.85, -367.75)
Camera target: (50.00, 110.00, 30.00)
Camera orientation: 4.8, 1.8, 410.0

##### above view

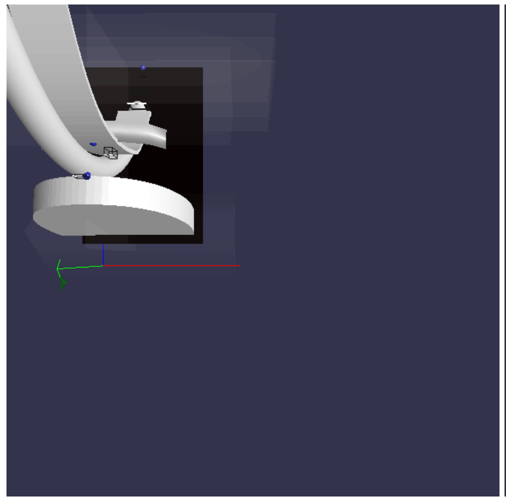

Camera position: (59.99, 316.98, -103.17)
Camera target: (60.00, 0.00, -100.00)
Camera orientation: 4.7, 0.0, 317.0

##### glottis view

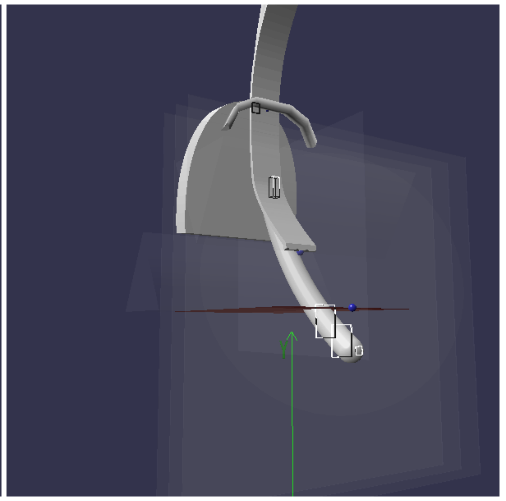

Camera position: (149.73, -6.98, 170.04)
Camera target: (0.00, 30.00, -72.00)
Camera orientation: 7.3, 1.7, 287.0

#### DLGent Yaw: 0.0° Roll: 45.0°

Scenario: #1 (resolved)
Subject: #1 DLGent
Yaw: 0.0°
Roll: 45.0°
Tube lift: -22.547 mm
Jaw contact X: 16.948 mm
Jaw contact Y: 99.983 mm
Tube position: (45.76, 83.11, -37.63)
Tube rotation: (56.81, 0.00, 45.00)
Upper incisor: (0.00, 107.00, -74.00)
Lower incisor: (0.00, 98.00, -60.00)
Glottic contact: (4.40, -22.55, -0.12)
Preglottic contact: (2.68, -17.98, -10.00)
Prepreglottic contact: (-0.22, -7.96, -20.00)
Maxillary contact: (16.95, 99.98, -74.46)
Blade contact: (2.24, 59.26, -54.57)
Vallecula: (--, 25.00, -33.00)

##### intubation view

##### side view

##### top view

##### above view

##### glottis view

Jaw width: 65.4mm
Resolve speed: 0.01
Cycles: 82
Cycles at speed: 3
Auto: On
Ghosts: Off
Cache hit: No
Results: 5
Camera position: (149.73, -6.98, 170.04)
Camera target: (0.00, 30.00, -72.00)
Camera orientation: 7.3, 1.7, 287.0

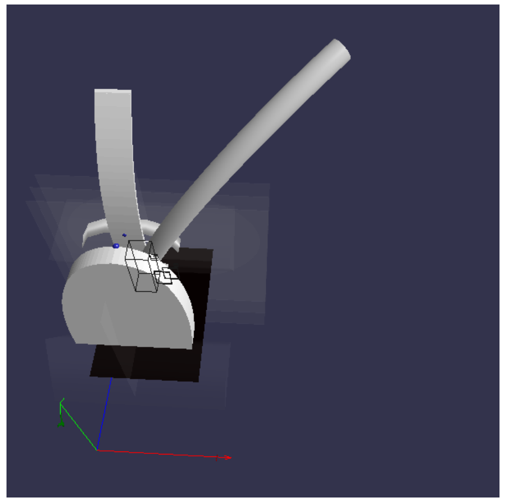

Camera position: (72.19, 330.13, -225.53)
Camera target: (60.00, 0.00, 0.00)
Camera orientation: -7.8, 0.6, 400.0

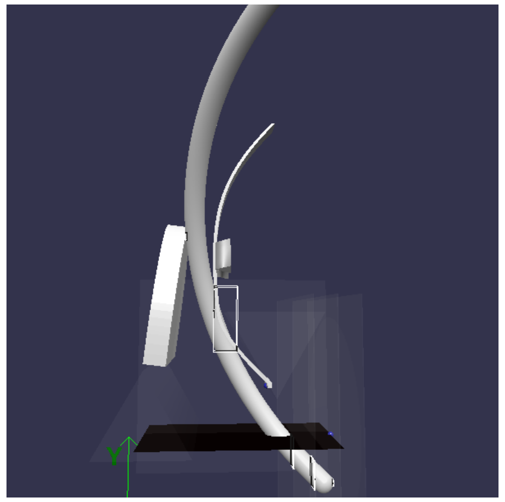

Camera position: (305.62, 61.30, -59.83)
Camera target: (8.00, 94.00, -41.00)
Camera orientation: 6.2, 1.7, 300.0

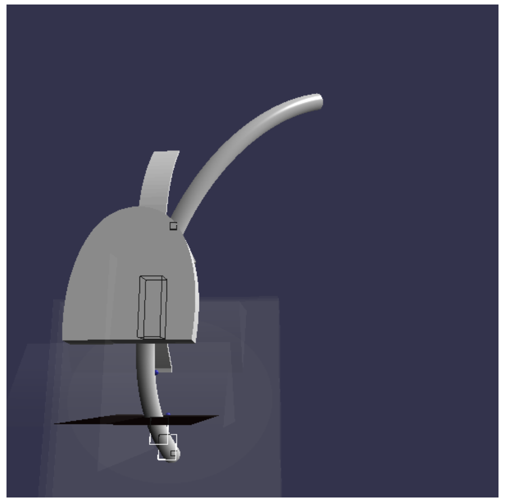

Camera position: (84.94, 16.85, -367.75)
Camera target: (50.00, 110.00, 30.00)
Camera orientation: 4.8, 1.8, 410.0

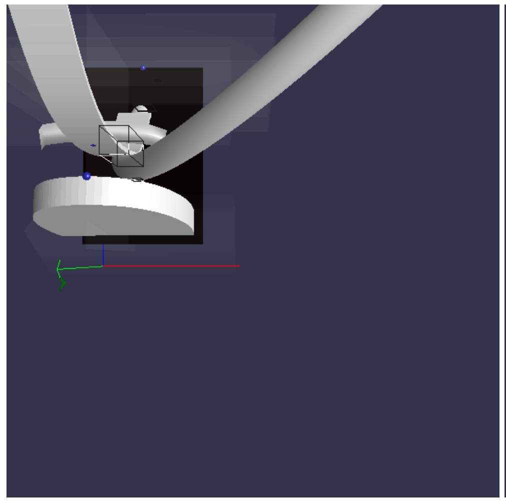

Camera position: (59.99, 316.98, -103.17)
Camera target: (60.00, 0.00, -100.00)
Camera orientation: 4.7, 0.0, 317.0

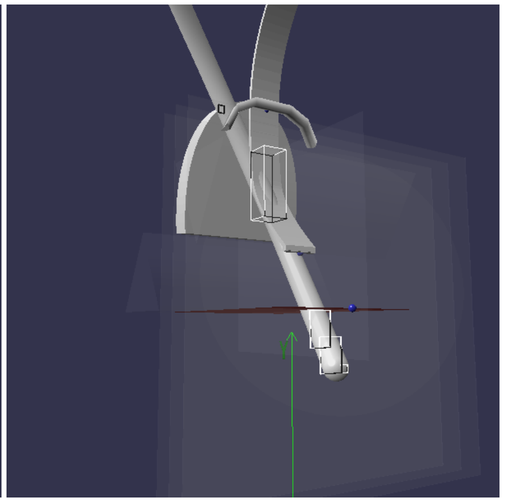

Camera position: (149.73, -6.98, 170.04)
Camera target: (0.00, 30.00, -72.00)
Camera orientation: 7.3, 1.7, 287.0

#### DLGent Yaw: 15.0° Roll: 0.0°

Scenario: #2 (resolved)
Subject: #1 DLGent
Yaw: 15.0°
Roll: 0.0°
Tube lift: -2.406 mm
Jaw contact X: 25.525 mm
Jaw contact Y: 88.895 mm
Tube position: (21.99, 110.32, -39.40)
Tube rotation: (51.46, 15.00, 0.00)
Upper incisor: (0.00, 107.00, -74.00)
Lower incisor: (0.00, 98.00, -60.00)
Glottic contact: (-3.00, -2.41, -0.12)
Preglottic contact: (-1.56, 0.52, -10.00)
Prepreglottic contact: (1.50, 7.17, -20.00)
Maxillary contact: (25.53, 88.90, -75.83)
Blade contact: (7.11, 25.04, -33.07)
Vallecula: (--, 25.00, -33.00)
Jaw width: 65.4mm
Resolve speed: 0.01
Cycles: 92
Cycles at speed: 3
Auto: On
Ghosts: Off
Cache hit: No
Results: 6
Camera position: (149.73, -6.98, 170.04)
Camera target: (0.00, 30.00, -72.00)
Camera orientation: 7.3, 1.7, 287.0

##### intubation view

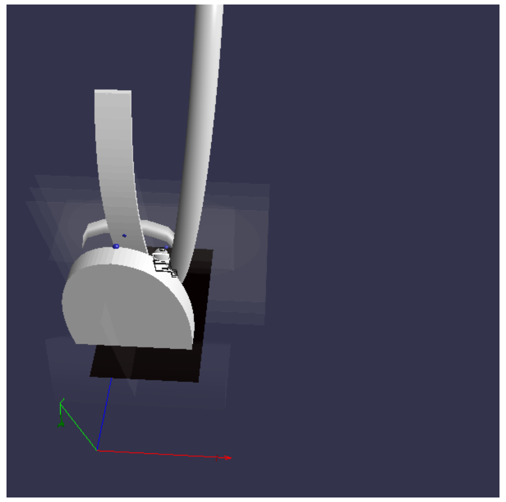

Camera position: (72.19, 330.13, -225.53)
Camera target: (60.00, 0.00, 0.00)
Camera orientation: -7.8, 0.6, 400.0

##### side view

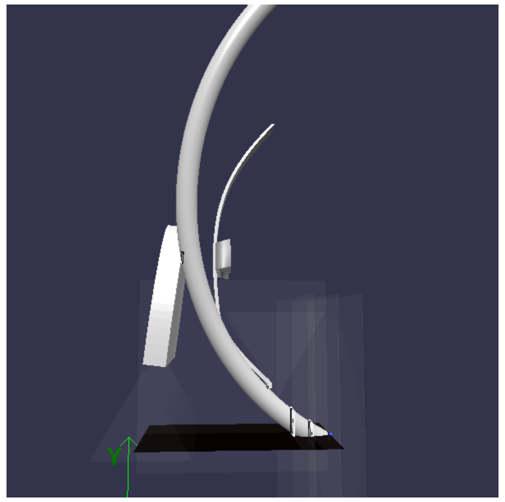

Camera position: (305.62, 61.30, -59.83)
Camera target: (8.00, 94.00, -41.00)
Camera orientation: 6.2, 1.7, 300.0

##### top view

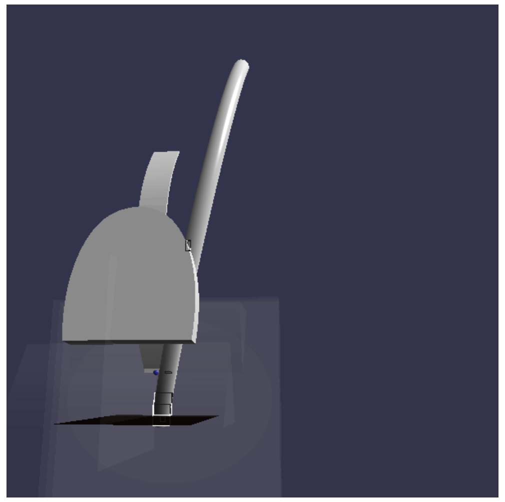

Camera position: (84.94, 16.85, -367.75)
Camera target: (50.00, 110.00, 30.00)
Camera orientation: 4.8, 1.8, 410.0

##### above view

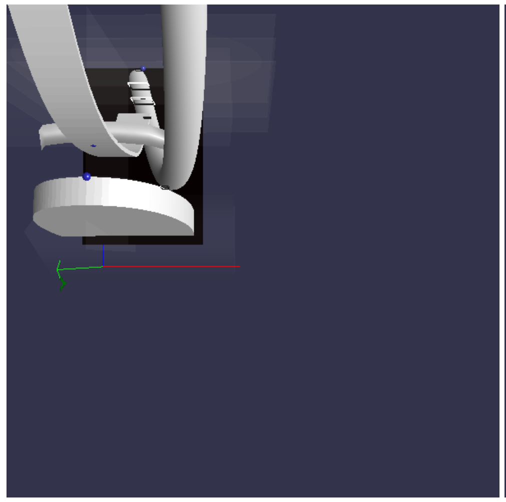

Camera position: (59.99, 316.98, -103.17)
Camera target: (60.00, 0.00, -100.00)
Camera orientation: 4.7, 0.0, 317.0

##### glottis view

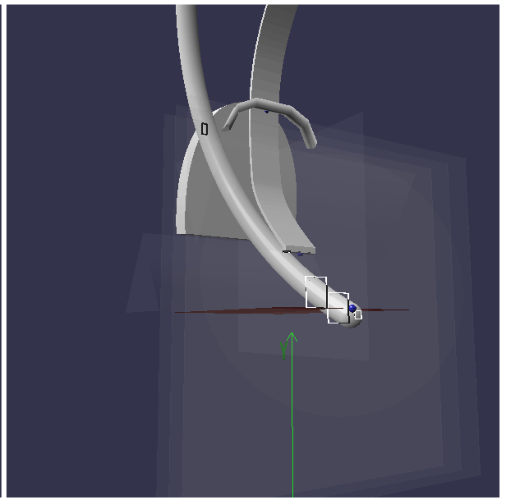

Camera position: (149.73, -6.98, 170.04)
Camera target: (0.00, 30.00, -72.00)
Camera orientation: 7.3, 1.7, 287.0

#### DLGent Yaw: 15.0° Roll: 45.0°

Scenario: #3 (resolved)
Subject: #1 DLGent
Yaw: 15.0°
Roll: 45.0°
Tube lift: -8.217 mm
Jaw contact X: 28.613 mm
Jaw contact Y: 80.913 mm
Tube position: (65.55, 83.79, -41.01)
Tube rotation: (51.13, 15.00, 45.00)
Upper incisor: (0.00, 107.00, -74.00)
Lower incisor: (0.00, 98.00, -60.00)
Glottic contact: (-2.97, -8.22, -0.12)
Preglottic contact: (-2.99, -4.45, -10.00)
Prepreglottic contact: (-2.38, 3.70, -20.00)
Maxillary contact: (28.61, 80.91, -76.82)
Blade contact: (7.97, 46.21, -51.06)
Vallecula: (--, 25.00, -33.00)
Jaw width: 65.4mm
Resolve speed: 0.01
Cycles: 87
Cycles at speed: 1
Auto: On
Ghosts: Off
Cache hit: No
Results: 7

##### intubation view

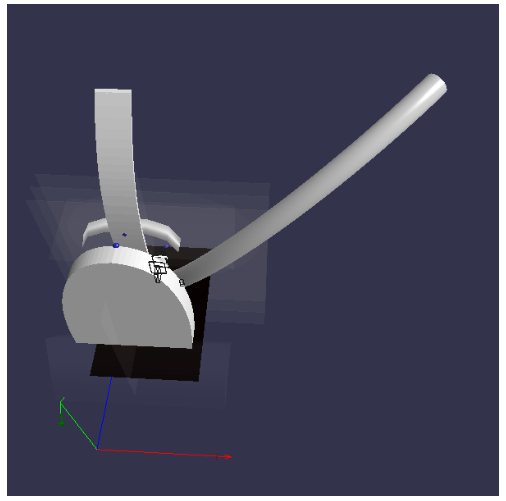

Camera position: (72.19, 330.13, -225.53)
Camera target: (60.00, 0.00, 0.00)

##### side view

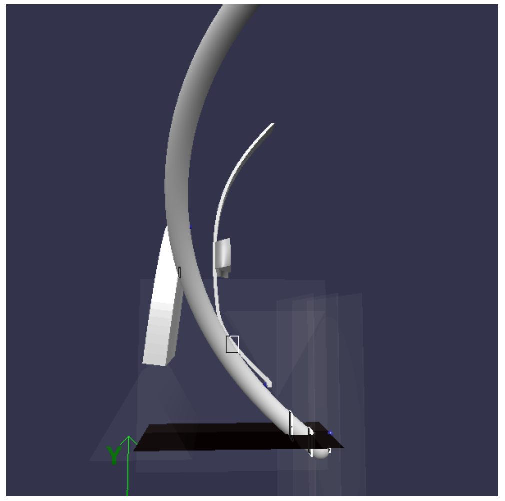

Camera position: (305.62, 61.30, -59.83)
Camera target: (8.00, 94.00, -41.00)

##### top view

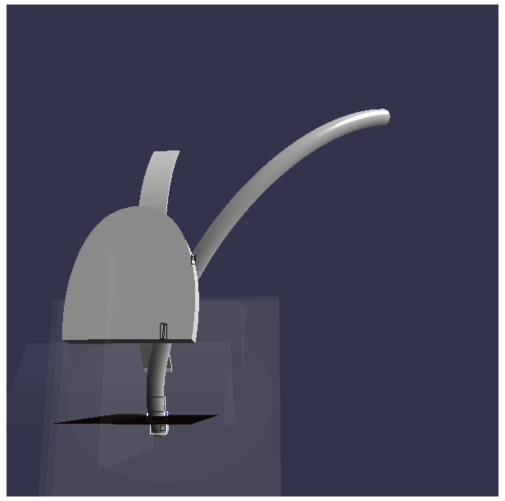

Camera position: (84.94, 16.85, -367.75)
Camera target: (50.00, 110.00, 30.00)

##### above view

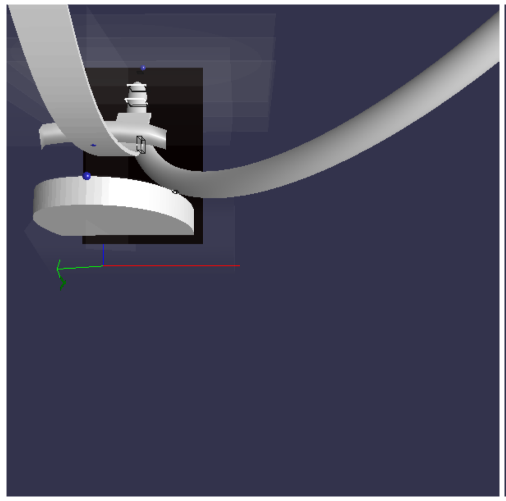

Camera position: (59.99, 316.98, -103.17)
Camera target: (60.00, 0.00, -100.00)

##### glottis view

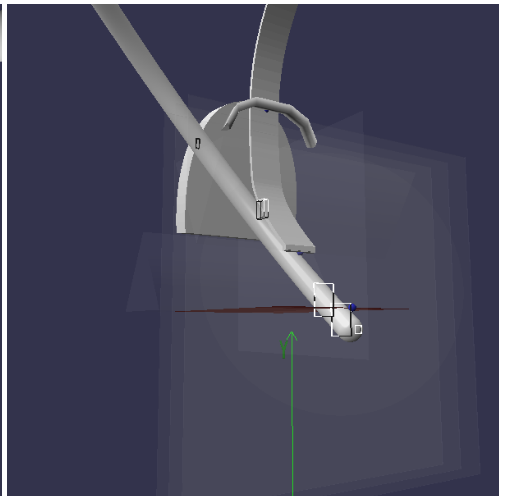

Camera position: (149.73, -6.98, 170.04)
Camera target: (0.00, 30.00, -72.00)

Camera position: (149.73, -6.98, 170.04)
Camera target: (0.00, 30.00, -72.00)
Camera orientation: 7.3, 1.7, 287.0

Camera orientation: -7.8, 0.6, 400.0

Camera orientation: 6.2, 1.7, 300.0

Camera orientation: 4.8, 1.8, 410.0

Camera orientation: 4.7, 0.0, 317.0

Camera orientation: 7.3, 1.7, 287.0

#### EL50N Yaw: 0.0° Roll: 0.0°

Scenario: #0 (resolved)
Subject: #2 EL50N
Yaw: 0.0°
Roll: 0.0°
Tube lift: -0.449 mm
Jaw contact X: -2.788 mm
Jaw contact Y: 101.038 mm
Tube position: (-3.00, 111.34, -51.59)
Tube rotation: (41.84, 0.00, 0.00)
Upper incisor: (0.00, 102.00, -100.00)
Lower incisor: (0.00, 100.00, -82.00)
Glottic contact: (-3.00, -0.45, -0.12)
Preglottic contact: (-3.00, 1.48, -10.00)
Prepreglottic contact: (-3.00, 5.94, -20.00)
Maxillary contact: (-2.79, 101.04, -99.79)
Blade contact: (-3.00, 49.70, -65.40)
Vallecula: (--, 25.00, -33.00)
Jaw width: 65.4mm
Resolve speed: 0.01
Cycles: 99
Cycles at speed: 3
Auto: On
Ghosts: Off
Cache hit: No
Results: 8
Camera position: (149.73, -6.98, 170.04)
Camera target: (0.00, 30.00, -72.00)
Camera orientation: 7.3, 1.7, 287.0

##### intubation view

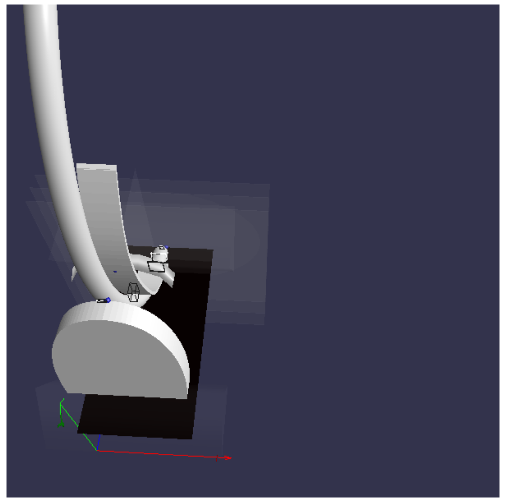

Camera position: (72.19, 330.13, -225.53)
Camera target: (60.00, 0.00, 0.00)
Camera orientation: -7.8, 0.6, 400.0

##### side view

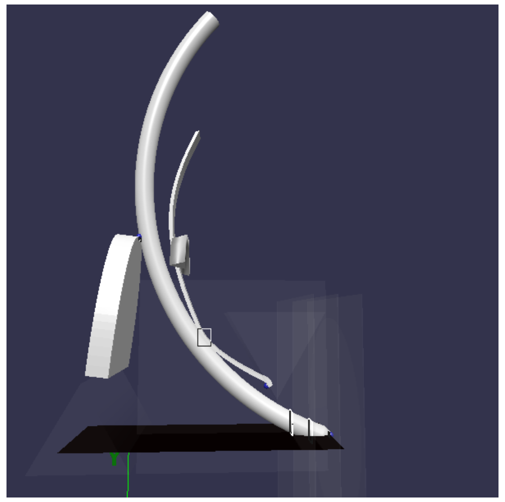

Camera position: (305.62, 61.30, -59.83)
Camera target: (8.00, 94.00, -41.00)
Camera orientation: 6.2, 1.7, 300.0

##### top view

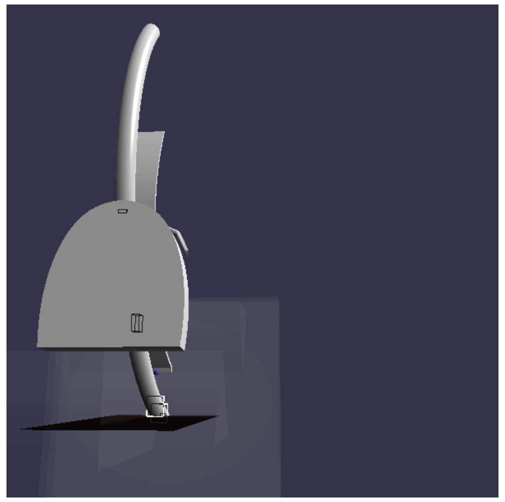

Camera position: (84.94, 16.85, -367.75)
Camera target: (50.00, 110.00, 30.00)
Camera orientation: 4.8, 1.8, 410.0

##### above view

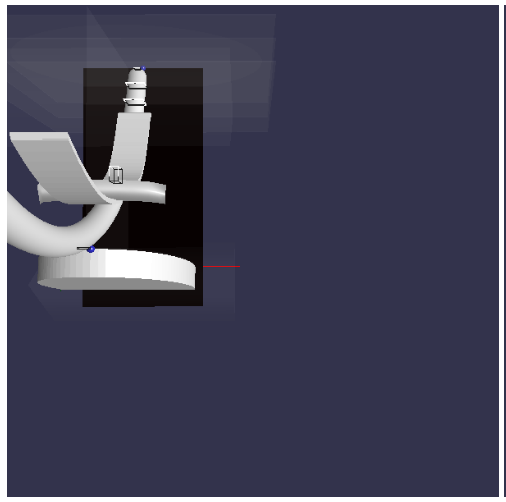

Camera position: (59.99, 316.98, -103.17)
Camera target: (60.00, 0.00, -100.00)
Camera orientation: 4.7, 0.0, 317.0

##### glottis view

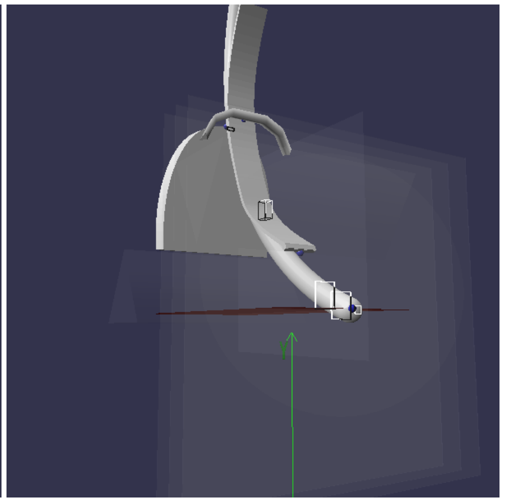

Camera position: (149.73, -6.98, 170.04)
Camera target: (0.00, 30.00, -72.00)
Camera orientation: 7.3, 1.7, 287.0

#### EL50N Yaw: 0.0° Roll: 45.0°

Scenario: #1 (resolved)
Subject: #2 EL50N
Yaw: 0.0°
Roll: 45.0°
Tube lift: -9.945 mm
Jaw contact X: 16.88 mm
Jaw contact Y: 95.557 mm
Tube position: (45.76, 89.63, -53.39)
Tube rotation: (43.90, 0.00, 45.00)
Upper incisor: (0.00, 102.00, -100.00)
Lower incisor: (0.00, 100.00, -82.00)
Glottic contact: (4.40, -9.94, -0.12)
Preglottic contact: (2.92, -7.16, -10.00)
Prepreglottic contact: (0.37, -1.05, -20.00)
Maxillary contact: (16.88, 95.56, -100.45)
Blade contact: (-0.12, 49.53, -66.19)
Vallecula: (--, 25.00, -33.00)
Jaw width: 65.4mm
Resolve speed: 0.01
Cycles: 103
Cycles at speed: 3
Auto: On
Ghosts: Off
Cache hit: No
Results: 9
Camera position: (149.73, -6.98, 170.04)
Camera target: (0.00, 30.00, -72.00)
Camera orientation: 7.3, 1.7, 287.0

##### intubation view

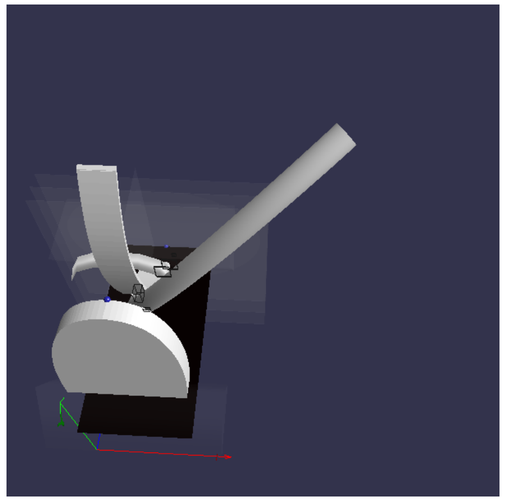

Camera position: (72.19, 330.13, -225.53)
Camera target: (60.00, 0.00, 0.00)
Camera orientation: -7.8, 0.6, 400.0

##### side view

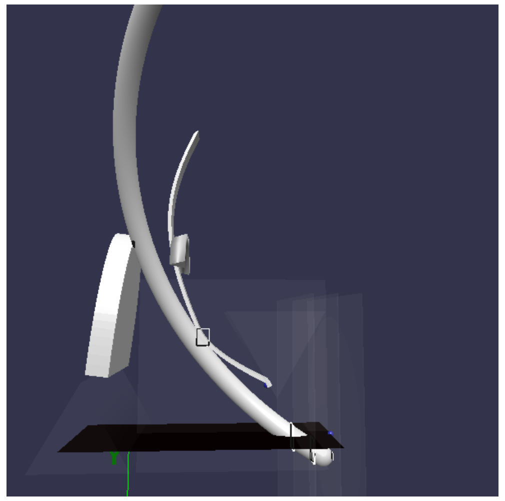

Camera position: (305.62, 61.30, -59.83)
Camera target: (8.00, 94.00, -41.00)
Camera orientation: 6.2, 1.7, 300.0

##### top view

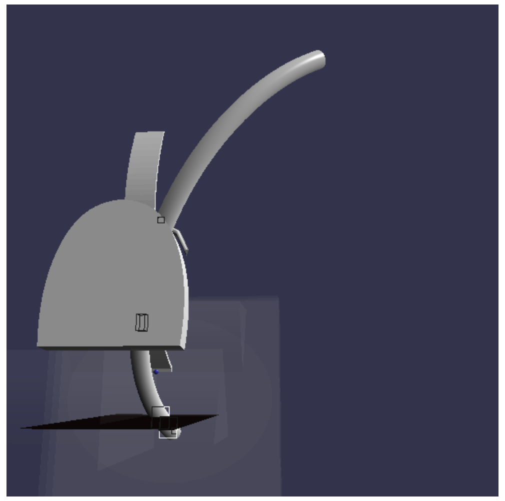

Camera position: (84.94, 16.85, -367.75)
Camera target: (50.00, 110.00, 30.00)
Camera orientation: 4.8, 1.8, 410.0

##### above view

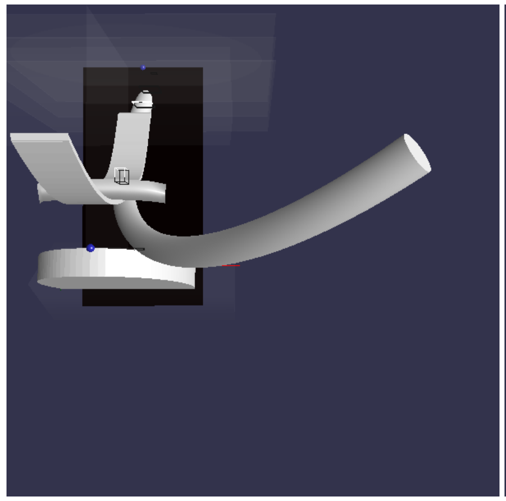

Camera position: (59.99, 316.98, -103.17)
Camera target: (60.00, 0.00, -100.00)
Camera orientation: 4.7, 0.0, 317.0

##### glottis view

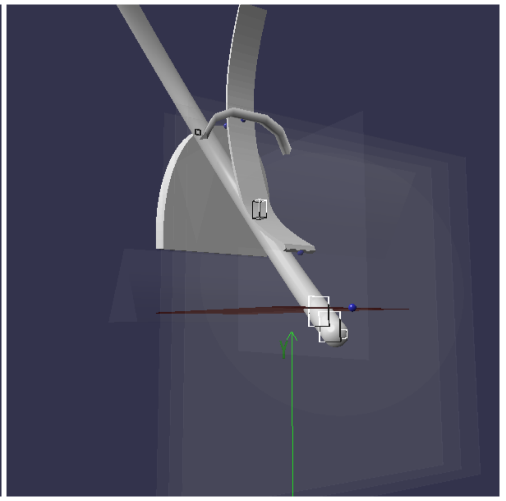

Camera position: (149.73, -6.98, 170.04)
Camera target: (0.00, 30.00, -72.00)
Camera orientation: 7.3, 1.7, 287.0

#### EL50N Yaw: 15.0° Roll: 0.0°

Scenario: #2 (resolved)
Subject: #2 EL50N
Yaw: 15.0°
Roll: 0.0°
Tube lift: 10.506 mm
Jaw contact X: 25.876 mm
Jaw contact Y: 83.23 mm
Tube position: (21.99, 115.50, -57.98)
Tube rotation: (34.38, 15.00, 0.00)
Upper incisor: (0.00, 102.00, -100.00)
Lower incisor: (0.00, 100.00, -82.00)
Glottic contact: (-3.00, 10.51, -0.12)
Preglottic contact: (-1.72, 11.64, -10.00)
Prepreglottic contact: (0.92, 14.49, -20.00)
Maxillary contact: (25.88, 83.23, -101.93)
Blade contact: (5.19, 24.96, -33.17)
Vallecula: (--, 25.00, -33.00)

##### intubation view

##### side view

##### top view

##### above view

##### glottis view

Jaw width: 65.4mm
Resolve speed: 0.01
Cycles: 98
Cycles at speed: 3
Auto: On
Ghosts: Off
Cache hit: No
Results: 10
Camera position: (149.73, -6.98, 170.04)
Camera target: (0.00, 30.00, -72.00)
Camera orientation: 7.3, 1.7, 287.0

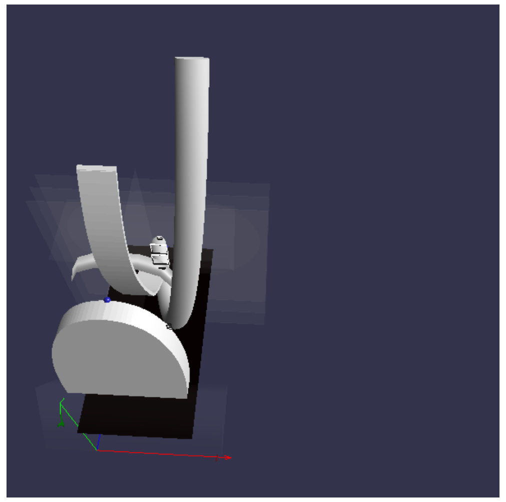

Camera position: (72.19, 330.13, -225.53)
Camera target: (60.00, 0.00, 0.00)
Camera orientation: -7.8, 0.6, 400.0

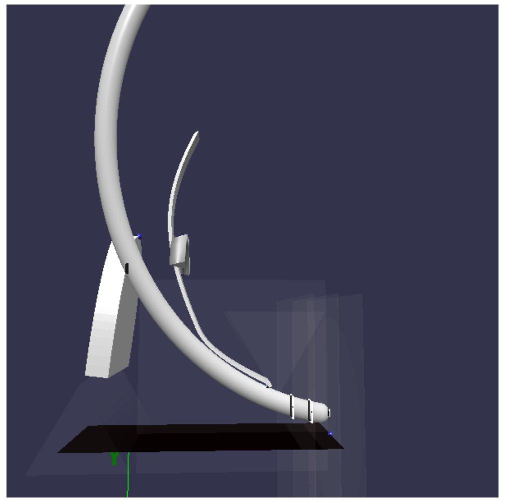

Camera position: (305.62, 61.30, -59.83)
Camera target: (8.00, 94.00, -41.00)
Camera orientation: 6.2, 1.7, 300.0

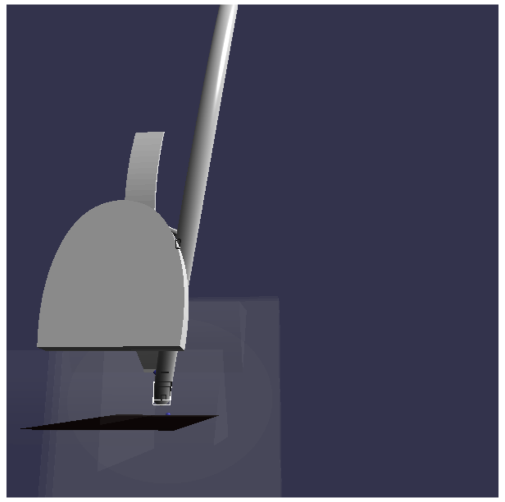

Camera position: (84.94, 16.85, -367.75)
Camera target: (50.00, 110.00, 30.00)
Camera orientation: 4.8, 1.8, 410.0

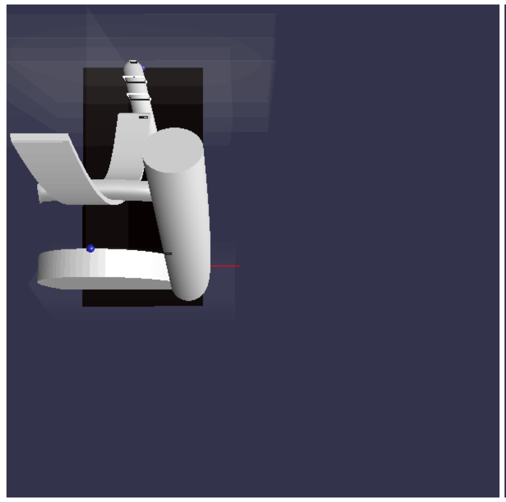

Camera position: (59.99, 316.98, -103.17)
Camera target: (60.00, 0.00, -100.00)
Camera orientation: 4.7, 0.0, 317.0

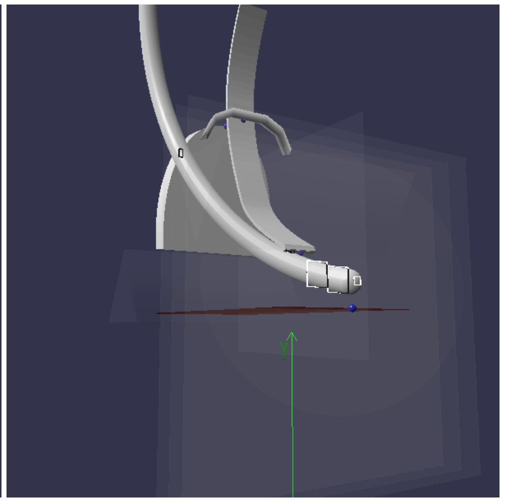

Camera position: (149.73, -6.98, 170.04)
Camera target: (0.00, 30.00, -72.00)
Camera orientation: 7.3, 1.7, 287.0

#### EL50N Yaw: 15.0° Roll: 45.0°

Scenario: #3 (resolved)
Subject: #2 EL50N
Yaw: 15.0°
Roll: 45.0°
Tube lift: 8.768 mm
Jaw contact X: 29.493 mm
Jaw contact Y: 73.215 mm
Tube position: (65.55, 90.58, -60.48)
Tube rotation: (32.24, 15.00, 45.00)
Upper incisor: (0.00, 102.00, -100.00)
Lower incisor: (0.00, 100.00, -82.00)
Glottic contact: (-2.97, 8.77, -0.12)
Preglottic contact: (-3.00, 10.36, -10.00)
Prepreglottic contact: (-2.62, 13.93, -20.00)
Maxillary contact: (29.49, 73.22, -103.17)
Blade contact: (-0.83, 24.98, -33.17)
Vallecula: (--, 25.00, -33.00)
Jaw width: 65.4mm
Resolve speed: 0.01
Cycles: 84
Cycles at speed: 1
Auto: On
Ghosts: Off
Cache hit: No
Results: 11
Camera position: (149.73, -6.98, 170.04)
Camera target: (0.00, 30.00, -72.00)
Camera orientation: 7.3, 1.7, 287.0

##### intubation view

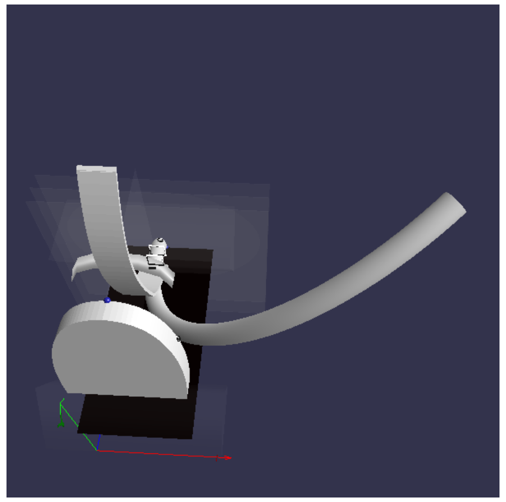

Camera position: (72.19, 330.13, -225.53)
Camera target: (60.00, 0.00, 0.00)
Camera orientation: -7.8, 0.6, 400.0

##### side view

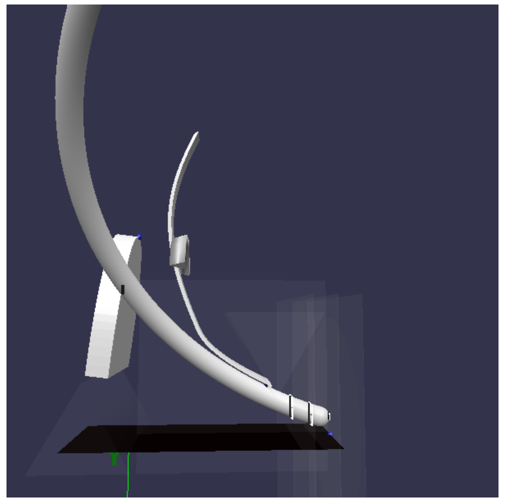

Camera position: (305.62, 61.30, -59.83)
Camera target: (8.00, 94.00, -41.00)
Camera orientation: 6.2, 1.7, 300.0

##### top view

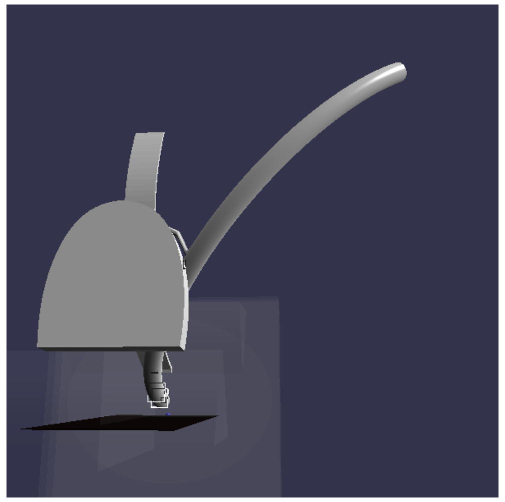

Camera position: (84.94, 16.85, -367.75)
Camera target: (50.00, 110.00, 30.00)
Camera orientation: 4.8, 1.8, 410.0

##### above view

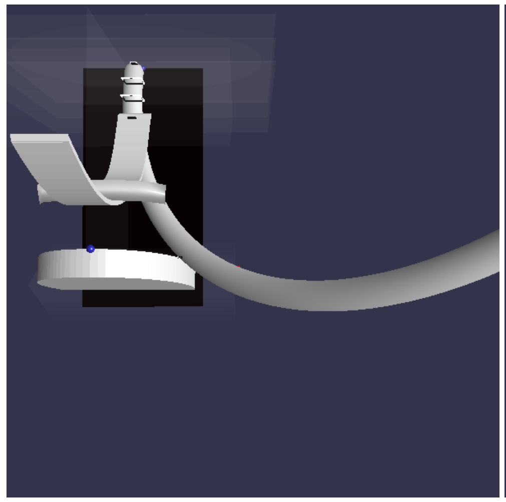

Camera position: (59.99, 316.98, -103.17)
Camera target: (60.00, 0.00, -100.00)
Camera orientation: 4.7, 0.0, 317.0

##### glottis view

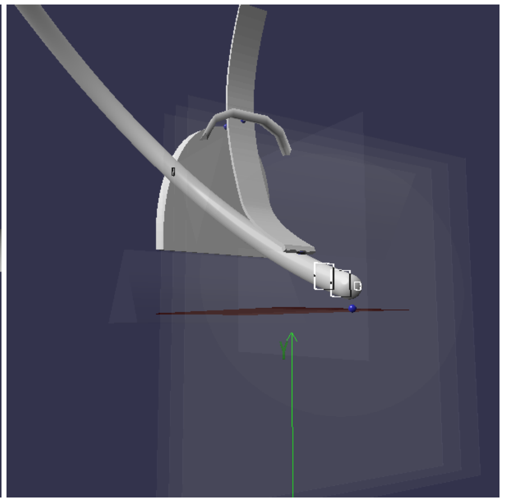

Camera position: (149.73, -6.98, 170.04)
Camera target: (0.00, 30.00, -72.00)
Camera orientation: 7.3, 1.7, 287.0

#### DL50N Yaw: 0.0° Roll: 0.0°

Scenario: #0 (resolved)
Subject: #3 DL50N
Yaw: 0.0°
Roll: 0.0°
Tube lift: -10.026 mm
Jaw contact X: -2.886 mm
Jaw contact Y: 106.199 mm
Tube position: (-3.00, 105.24, -41.08)
Tube rotation: (51.28, 0.00, 0.00)
Upper incisor: (0.00, 108.00, -82.00)
Lower incisor: (0.00, 97.00, -66.00)
Glottic contact: (-3.00, -10.03, -0.12)
Preglottic contact: (-3.00, -7.04, -10.00)
Prepreglottic contact: (-3.00, -0.33, -20.00)
Maxillary contact: (-2.89, 106.20, -81.79)
Blade contact: (-3.00, 56.27, -59.80)
Vallecula: (--, 25.00, -33.00)
Jaw width: 65.4mm
Resolve speed: 0.01
Cycles: 104
Cycles at speed: 2
Auto: On
Ghosts: Off
Cache hit: No
Results: 12

##### intubation view

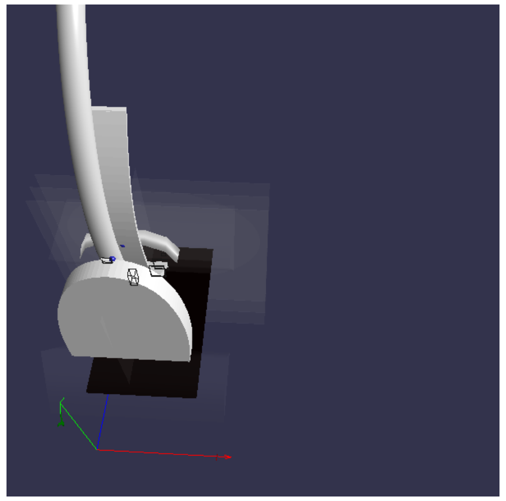

Camera position: (72.19, 330.13, -225.53)
Camera target: (60.00, 0.00, 0.00)

##### side view

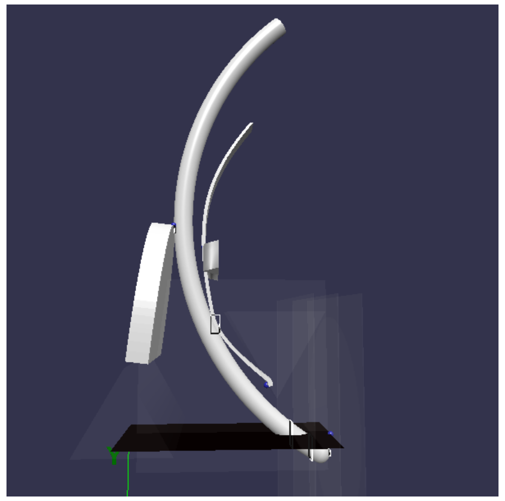

Camera position: (305.62, 61.30, -59.83)
Camera target: (8.00, 94.00, -41.00)

##### top view

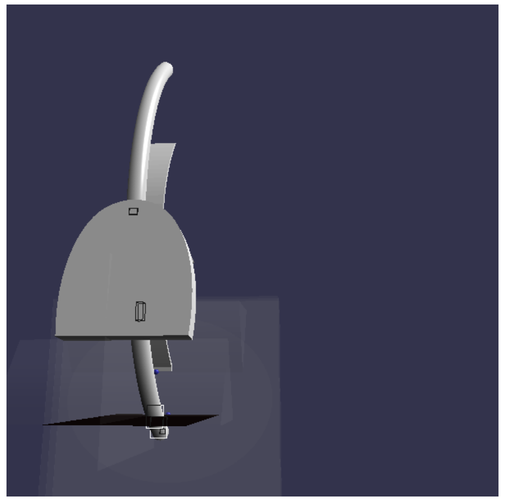

Camera position: (84.94, 16.85, -367.75)
Camera target: (50.00, 110.00, 30.00)

##### above view

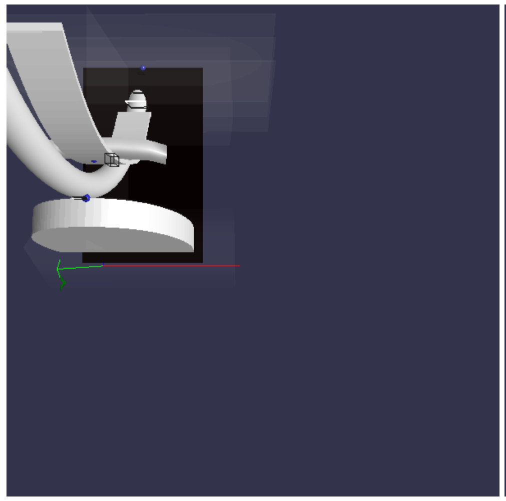

Camera position: (59.99, 316.98, -103.17)
Camera target: (60.00, 0.00, -100.00)

##### glottis view

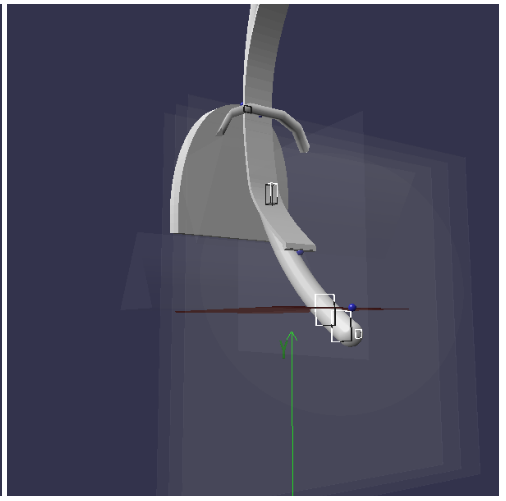

Camera position: (149.73, -6.98, 170.04)
Camera target: (0.00, 30.00, -72.00)

Camera position: (149.73, -6.98, 170.04)
Camera target: (0.00, 30.00, -72.00)
Camera orientation: 7.3, 1.7, 287.0

Camera orientation: -7.8, 0.6, 400.0

Camera orientation: 6.2, 1.7, 300.0

Camera orientation: 4.8, 1.8, 410.0

Camera orientation: 4.7, 0.0, 317.0

Camera orientation: 7.3, 1.7, 287.0

#### DL50N Yaw: 0.0° Roll: 45.0°

Scenario: #1 (resolved)
Subject: #3 DL50N
Yaw: 0.0°
Roll: 45.0°
Tube lift: -22.186 mm
Jaw contact X: 18.585 mm
Jaw contact Y: 99.72 mm
Tube position: (45.76, 82.29, -41.99)
Tube rotation: (53.15, 0.00, 45.00)
Upper incisor: (0.00, 108.00, -82.00)
Lower incisor: (0.00, 97.00, -66.00)
Glottic contact: (4.40, -22.19, -0.12)
Preglottic contact: (2.76, -18.20, -10.00)
Prepreglottic contact: (-0.02, -9.45, -20.00)
Maxillary contact: (18.58, 99.72, -82.64)
Blade contact: (2.07, 56.11, -60.47)
Vallecula: (--, 25.00, -33.00)
Jaw width: 65.4mm
Resolve speed: 0.01
Cycles: 85
Cycles at speed: 1
Auto: On
Ghosts: Off
Cache hit: No
Results: 13
Camera position: (149.73, -6.98, 170.04)
Camera target: (0.00, 30.00, -72.00)
Camera orientation: 7.3, 1.7, 287.0

##### intubation view

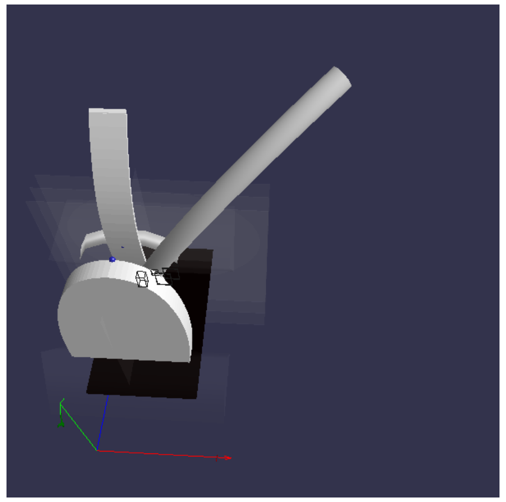

Camera position: (72.19, 330.13, -225.53)
Camera target: (60.00, 0.00, 0.00)
Camera orientation: -7.8, 0.6, 400.0

##### side view

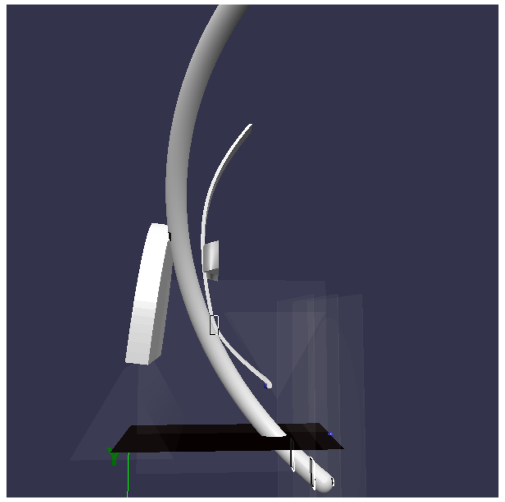

Camera position: (305.62, 61.30, -59.83)
Camera target: (8.00, 94.00, -41.00)
Camera orientation: 6.2, 1.7, 300.0

##### top view

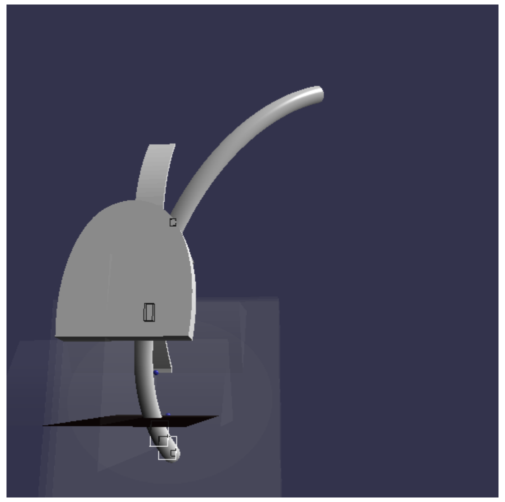

Camera position: (84.94, 16.85, -367.75)
Camera target: (50.00, 110.00, 30.00)
Camera orientation: 4.8, 1.8, 410.0

##### above view

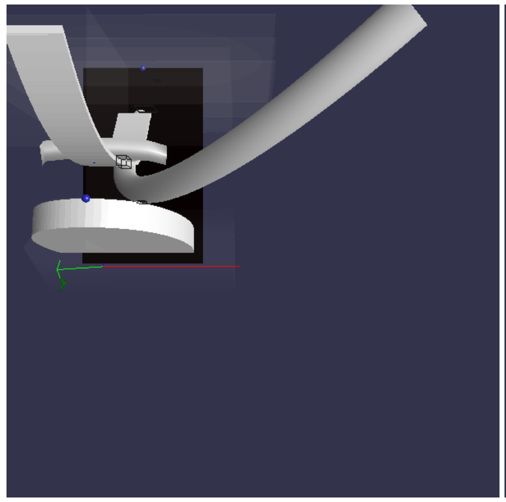

Camera position: (59.99, 316.98, -103.17)
Camera target: (60.00, 0.00, -100.00)
Camera orientation: 4.7, 0.0, 317.0

##### glottis view

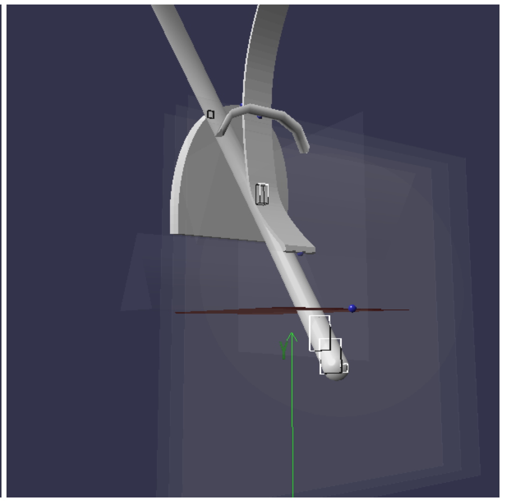

Camera position: (149.73, -6.98, 170.04)
Camera target: (0.00, 30.00, -72.00)
Camera orientation: 7.3, 1.7, 287.0

#### DL50N Yaw: 15.0° Roll: 0.0°

Scenario: #2 (resolved)
Subject: #3 DL50N
Yaw: 15.0°
Roll: 0.0°
Tube lift: 1.982 mm
Jaw contact X: 25.75 mm
Jaw contact Y: 89.449 mm
Tube position: (21.99, 113.52, -44.48)
Tube rotation: (46.57, 15.00, 0.00)
Upper incisor: (0.00, 108.00, -82.00)
Lower incisor: (0.00, 97.00, -66.00)
Glottic contact: (-3.00, 1.98, -0.12)
Preglottic contact: (-1.62, 4.35, -10.00)
Prepreglottic contact: (1.27, 9.77, -20.00)
Maxillary contact: (25.75, 89.45, -83.89)
Blade contact: (6.48, 25.02, -33.09)
Vallecula: (--, 25.00, -33.00)
Jaw width: 65.4mm
Resolve speed: 0.01
Cycles: 90
Cycles at speed: 1
Auto: On
Ghosts: Off
Cache hit: No
Results: 14
Camera position: (149.73, -6.98, 170.04)
Camera target: (0.00, 30.00, -72.00)
Camera orientation: 7.3, 1.7, 287.0

##### intubation view

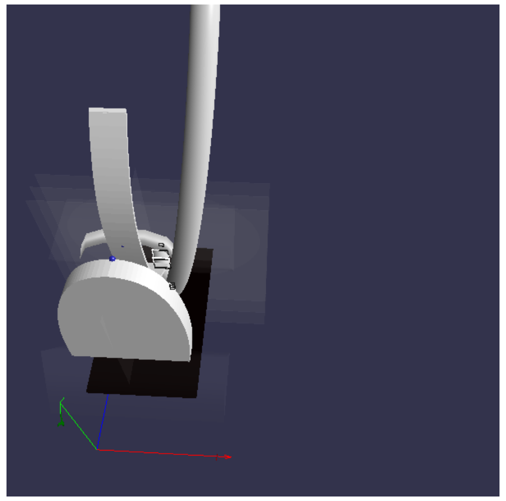

Camera position: (72.19, 330.13, -225.53)
Camera target: (60.00, 0.00, 0.00)
Camera orientation: -7.8, 0.6, 400.0

##### side view

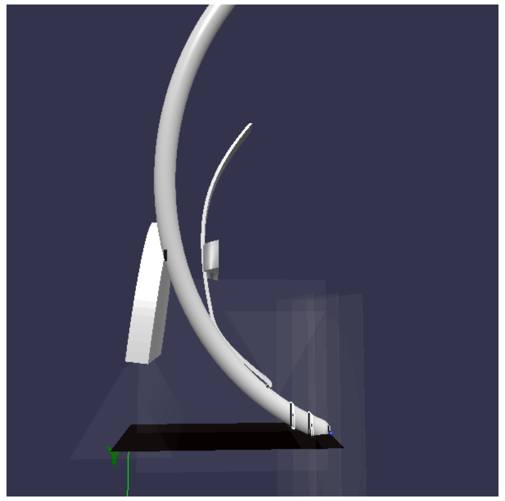

Camera position: (305.62, 61.30, -59.83)
Camera target: (8.00, 94.00, -41.00)
Camera orientation: 6.2, 1.7, 300.0

##### top view

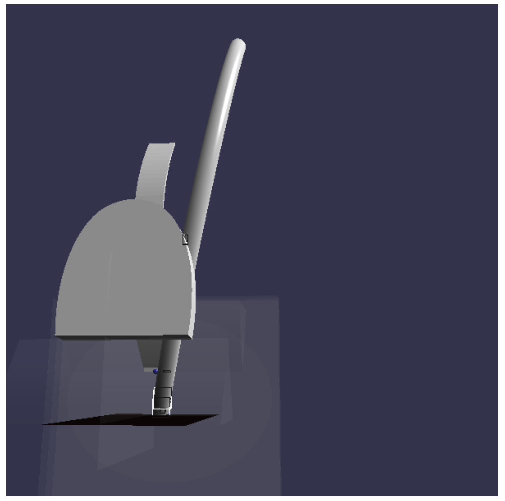

Camera position: (84.94, 16.85, -367.75)
Camera target: (50.00, 110.00, 30.00)
Camera orientation: 4.8, 1.8, 410.0

##### above view

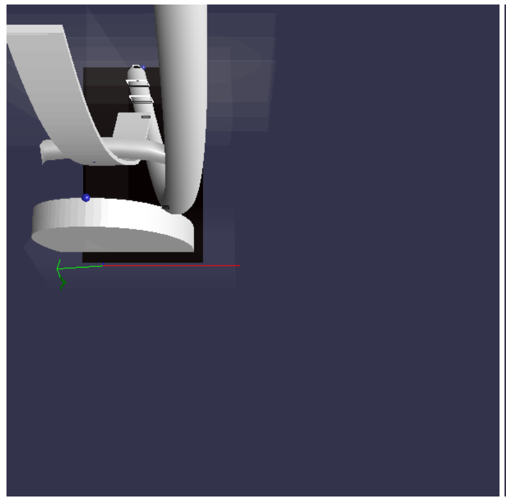

Camera position: (59.99, 316.98, -103.17)
Camera target: (60.00, 0.00, -100.00)
Camera orientation: 4.7, 0.0, 317.0

##### glottis view

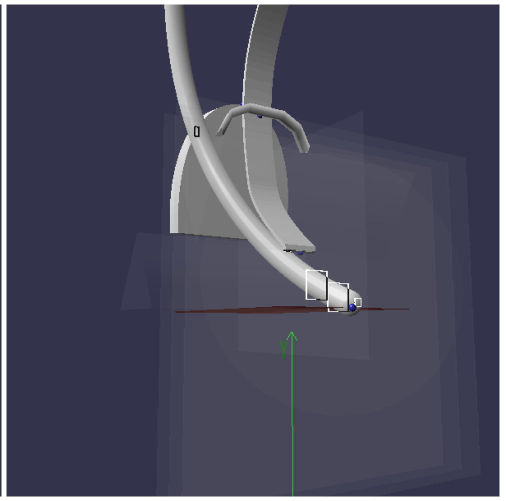

Camera position: (149.73, -6.98, 170.04)
Camera target: (0.00, 30.00, -72.00)
Camera orientation: 7.3, 1.7, 287.0

#### DL50N Yaw: 15.0° Roll: 45.0°

Scenario: #3 (resolved)
Subject: #3 DL50N
Yaw: 15.0°
Roll: 45.0°
Tube lift: -1.804 mm
Jaw contact X: 28.824 mm
Jaw contact Y: 81.367 mm
Tube position: (65.55, 88.33, -46.50)
Tube rotation: (45.85, 15.00, 45.00)
Upper incisor: (0.00, 108.00, -82.00)
Lower incisor: (0.00, 97.00, -66.00)
Glottic contact: (-2.97, -1.80, -0.12)
Preglottic contact: (-3.00, 1.27, -10.00)
Prepreglottic contact: (-2.48, 7.92, -20.00)
Maxillary contact: (28.82, 81.37, -84.90)
Blade contact: (4.39, 37.73, -45.62)
Vallecula: (--, 25.00, -33.00)

##### intubation view

##### side view

##### top view

##### above view

##### glottis view

Jaw width: 65.4mm
Resolve speed: 0.01
Cycles: 91
Cycles at speed: 2
Auto: On
Ghosts: Off
Cache hit: No
Results: 15
Camera position: (149.73, -6.98, 170.04)
Camera target: (0.00, 30.00, -72.00)
Camera orientation: 7.3, 1.7, 287.0

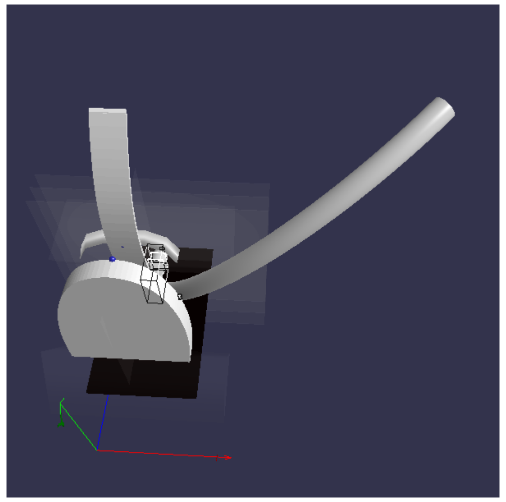

Camera position: (72.19, 330.13, -225.53)
Camera target: (60.00, 0.00, 0.00)
Camera orientation: -7.8, 0.6, 400.0

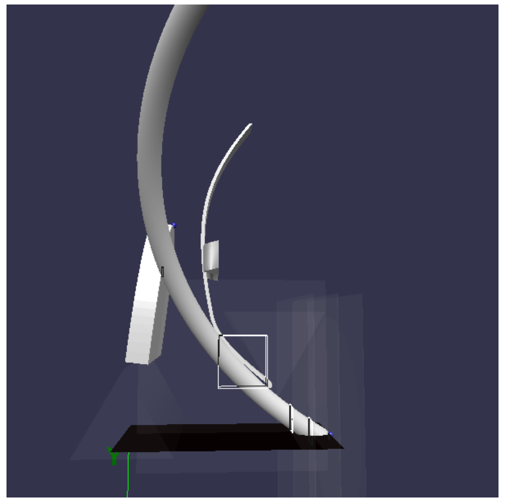

Camera position: (305.62, 61.30, -59.83)
Camera target: (8.00, 94.00, -41.00)
Camera orientation: 6.2, 1.7, 300.0

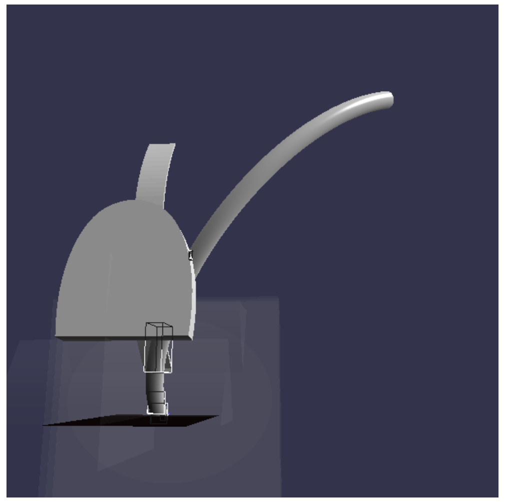

Camera position: (84.94, 16.85, -367.75)
Camera target: (50.00, 110.00, 30.00)
Camera orientation: 4.8, 1.8, 410.0

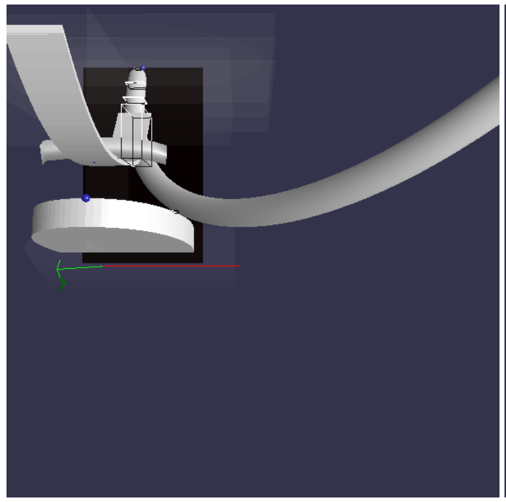

Camera position: (59.99, 316.98, -103.17)
Camera target: (60.00, 0.00, -100.00)
Camera orientation: 4.7, 0.0, 317.0

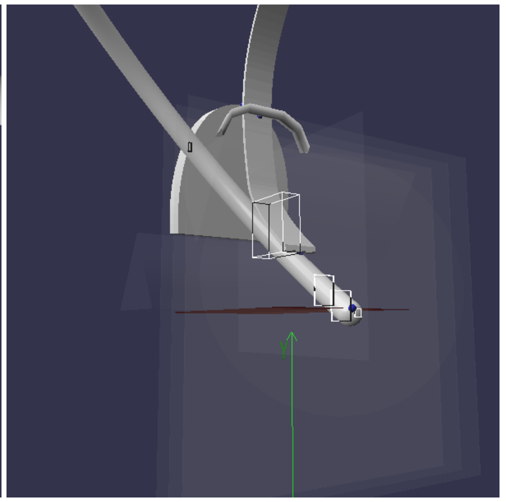

Camera position: (149.73, -6.98, 170.04)
Camera target: (0.00, 30.00, -72.00)
Camera orientation: 7.3, 1.7, 287.0
