## Supplementary material for "Optimal tracheal tube rotation patterns for navigating through the glottis: an in-silico quantification": Full Renders: AirwaySimulatorFull.html

Airway simulator


### Model

### Gallery

|  |  |  |  |  |  |
| --- | --- | --- | --- | --- | --- |
| ELGent Yaw: 0.0° Roll: 0.0°   ``` Scenario: #0 (resolved) Subject: #0 ELGent Yaw: 0.0° Roll: 0.0° Tube lift: -10.465 mm Jaw contact X: -2.89 mm Jaw contact Y: 108.426 mm Tube position: (-3.00, 103.97, -44.58) Tube rotation: (48.05, 0.00, 0.00) Upper incisor: (0.00, 110.00, -89.00) Lower incisor: (0.00, 102.00, -76.00) Glottic contact: (-3.00, -10.47, -0.12) Preglottic contact: (-3.00, -7.86, -10.00) Prepreglottic contact: (-3.00, -1.97, -20.00) Maxillary contact: (-2.89, 108.43, -88.78) Blade contact: (-3.00, 55.03, -64.65) Vallecula: (--, 25.00, -33.00) Jaw width: 65.4mm Resolve speed: 0.01 Cycles: 91 Cycles at speed: 1 Auto: On Ghosts: Off Cache hit: No Results: 0 Camera position: (305.62, 61.30, -59.83) Camera target: (8.00, 94.00, -41.00) Camera orientation: 6.2, 1.7, 300.0 ``` | intubation view   ```     Camera position: (20.99, 345.12, -201.12)     Camera target: (0.00, 0.00, 0.00)     Camera orientation: -7.8, 0.5, 400.0 ``` | side view   ```     Camera position: (305.62, 61.30, -59.83)     Camera target: (8.00, 94.00, -41.00)     Camera orientation: 6.2, 1.7, 300.0 ``` | top view   ```     Camera position: (11.26, 6.17, -377.84)     Camera target: (0.00, 110.00, 30.00)     Camera orientation: 4.7, 1.8, 421.0 ``` | above view   ```     Camera position: (-3.01, 315.98, -75.17)     Camera target: (-3.00, -1.00, -72.00)     Camera orientation: 4.7, 0.0, 317.0 ``` | glottis view   ```     Camera position: (122.84, -6.51, 185.88)     Camera target: (-3.00, -1.00, -72.00)     Camera orientation: 7.4, 1.6, 287.0 ``` |
| ELGent Yaw: 0.0° Roll: 15.0°   ``` Scenario: #1 (resolved) Subject: #0 ELGent Yaw: 0.0° Roll: 15.0° Tube lift: -13.223 mm Jaw contact X: 5.544 mm Jaw contact Y: 107.859 mm Tube position: (15.03, 99.88, -44.60) Tube rotation: (48.33, 0.00, 15.00) Upper incisor: (0.00, 110.00, -89.00) Lower incisor: (0.00, 102.00, -76.00) Glottic contact: (-0.30, -13.22, -0.12) Preglottic contact: (-0.82, -10.52, -10.00) Prepreglottic contact: (-1.74, -4.42, -20.00) Maxillary contact: (5.54, 107.86, -88.83) Blade contact: (-1.59, 54.84, -65.29) Vallecula: (--, 25.00, -33.00) Jaw width: 65.4mm Resolve speed: 0.01 Cycles: 97 Cycles at speed: 2 Auto: On Ghosts: Off Cache hit: No Results: 1 Camera position: (122.84, -6.51, 185.88) Camera target: (-3.00, -1.00, -72.00) Camera orientation: 7.4, 1.6, 287.0 ``` | intubation view   ```     Camera position: (20.99, 345.12, -201.12)     Camera target: (0.00, 0.00, 0.00)     Camera orientation: -7.8, 0.5, 400.0 ``` | side view   ```     Camera position: (305.62, 61.30, -59.83)     Camera target: (8.00, 94.00, -41.00)     Camera orientation: 6.2, 1.7, 300.0 ``` | top view   ```     Camera position: (11.26, 6.17, -377.84)     Camera target: (0.00, 110.00, 30.00)     Camera orientation: 4.7, 1.8, 421.0 ``` | above view   ```     Camera position: (-3.01, 315.98, -75.17)     Camera target: (-3.00, -1.00, -72.00)     Camera orientation: 4.7, 0.0, 317.0 ``` | glottis view   ```     Camera position: (122.84, -6.51, 185.88)     Camera target: (-3.00, -1.00, -72.00)     Camera orientation: 7.4, 1.6, 287.0 ``` |
| ELGent Yaw: 0.0° Roll: 30.0°   ``` Scenario: #2 (resolved) Subject: #0 ELGent Yaw: 0.0° Roll: 30.0° Tube lift: -16.548 mm Jaw contact X: 13.637 mm Jaw contact Y: 105.23 mm Tube position: (31.66, 92.65, -44.93) Tube rotation: (48.98, 0.00, 30.00) Upper incisor: (0.00, 110.00, -89.00) Lower incisor: (0.00, 102.00, -76.00) Glottic contact: (2.23, -16.55, -0.12) Preglottic contact: (1.18, -13.53, -10.00) Prepreglottic contact: (-0.64, -6.82, -20.00) Maxillary contact: (13.64, 105.23, -89.18) Blade contact: (0.15, 54.87, -65.31) Vallecula: (--, 25.00, -33.00) Jaw width: 65.4mm Resolve speed: 0.01 Cycles: 100 Cycles at speed: 3 Auto: On Ghosts: Off Cache hit: No Results: 2 Camera position: (122.84, -6.51, 185.88) Camera target: (-3.00, -1.00, -72.00) Camera orientation: 7.4, 1.6, 287.0 ``` | intubation view   ```     Camera position: (20.99, 345.12, -201.12)     Camera target: (0.00, 0.00, 0.00)     Camera orientation: -7.8, 0.5, 400.0 ``` | side view   ```     Camera position: (305.62, 61.30, -59.83)     Camera target: (8.00, 94.00, -41.00)     Camera orientation: 6.2, 1.7, 300.0 ``` | top view   ```     Camera position: (11.26, 6.17, -377.84)     Camera target: (0.00, 110.00, 30.00)     Camera orientation: 4.7, 1.8, 421.0 ``` | above view   ```     Camera position: (-3.01, 315.98, -75.17)     Camera target: (-3.00, -1.00, -72.00)     Camera orientation: 4.7, 0.0, 317.0 ``` | glottis view   ```     Camera position: (122.84, -6.51, 185.88)     Camera target: (-3.00, -1.00, -72.00)     Camera orientation: 7.4, 1.6, 287.0 ``` |
| ELGent Yaw: 0.0° Roll: 45.0°   ``` Scenario: #3 (resolved) Subject: #0 ELGent Yaw: 0.0° Roll: 45.0° Tube lift: -21.416 mm Jaw contact X: 20.297 mm Jaw contact Y: 99.923 mm Tube position: (45.76, 81.68, -45.86) Tube rotation: (49.97, 0.00, 45.00) Upper incisor: (0.00, 110.00, -89.00) Lower incisor: (0.00, 102.00, -76.00) Glottic contact: (4.41, -21.42, -0.12) Preglottic contact: (2.83, -17.87, -10.00) Prepreglottic contact: (0.14, -10.12, -20.00) Maxillary contact: (20.30, 99.92, -89.86) Blade contact: (2.24, 54.49, -65.15) Vallecula: (--, 25.00, -33.00) Jaw width: 65.4mm Resolve speed: 0.01 Cycles: 81 Cycles at speed: 1 Auto: On Ghosts: Off Cache hit: No Results: 3 Camera position: (122.84, -6.51, 185.88) Camera target: (-3.00, -1.00, -72.00) Camera orientation: 7.4, 1.6, 287.0 ``` | intubation view   ```     Camera position: (20.99, 345.12, -201.12)     Camera target: (0.00, 0.00, 0.00)     Camera orientation: -7.8, 0.5, 400.0 ``` | side view   ```     Camera position: (305.62, 61.30, -59.83)     Camera target: (8.00, 94.00, -41.00)     Camera orientation: 6.2, 1.7, 300.0 ``` | top view   ```     Camera position: (11.26, 6.17, -377.84)     Camera target: (0.00, 110.00, 30.00)     Camera orientation: 4.7, 1.8, 421.0 ``` | above view   ```     Camera position: (-3.01, 315.98, -75.17)     Camera target: (-3.00, -1.00, -72.00)     Camera orientation: 4.7, 0.0, 317.0 ``` | glottis view   ```     Camera position: (122.84, -6.51, 185.88)     Camera target: (-3.00, -1.00, -72.00)     Camera orientation: 7.4, 1.6, 287.0 ``` |
| ELGent Yaw: 5.0° Roll: 0.0°   ``` Scenario: #4 (resolved) Subject: #0 ELGent Yaw: 5.0° Roll: 0.0° Tube lift: -11.26 mm Jaw contact X: 9.135 mm Jaw contact Y: 107.126 mm Tube position: (5.42, 102.82, -44.68) Tube rotation: (47.79, 5.00, 0.00) Upper incisor: (0.00, 110.00, -89.00) Lower incisor: (0.00, 102.00, -76.00) Glottic contact: (-3.00, -11.26, -0.12) Preglottic contact: (-2.54, -8.69, -10.00) Prepreglottic contact: (-1.58, -2.88, -20.00) Maxillary contact: (9.14, 107.13, -88.94) Blade contact: (4.89, 54.56, -65.15) Vallecula: (--, 25.00, -33.00) Jaw width: 65.4mm Resolve speed: 0.01 Cycles: 91 Cycles at speed: 2 Auto: On Ghosts: Off Cache hit: No Results: 4 Camera position: (122.84, -6.51, 185.88) Camera target: (-3.00, -1.00, -72.00) Camera orientation: 7.4, 1.6, 287.0 ``` | intubation view   ```     Camera position: (20.99, 345.12, -201.12)     Camera target: (0.00, 0.00, 0.00)     Camera orientation: -7.8, 0.5, 400.0 ``` | side view   ```     Camera position: (305.62, 61.30, -59.83)     Camera target: (8.00, 94.00, -41.00)     Camera orientation: 6.2, 1.7, 300.0 ``` | top view   ```     Camera position: (11.26, 6.17, -377.84)     Camera target: (0.00, 110.00, 30.00)     Camera orientation: 4.7, 1.8, 421.0 ``` | above view   ```     Camera position: (-3.01, 315.98, -75.17)     Camera target: (-3.00, -1.00, -72.00)     Camera orientation: 4.7, 0.0, 317.0 ``` | glottis view   ```     Camera position: (122.84, -6.51, 185.88)     Camera target: (-3.00, -1.00, -72.00)     Camera orientation: 7.4, 1.6, 287.0 ``` |
| ELGent Yaw: 5.0° Roll: 15.0°   ``` Scenario: #5 (resolved) Subject: #0 ELGent Yaw: 5.0° Roll: 15.0° Tube lift: -11.224 mm Jaw contact X: 14.2 mm Jaw contact Y: 104.9 mm Tube position: (22.06, 100.34, -44.92) Tube rotation: (47.69, 5.00, 15.00) Upper incisor: (0.00, 110.00, -89.00) Lower incisor: (0.00, 102.00, -76.00) Glottic contact: (-2.93, -11.22, -0.12) Preglottic contact: (-2.99, -8.59, -10.00) Prepreglottic contact: (-2.93, -2.64, -20.00) Maxillary contact: (14.20, 104.90, -89.22) Blade contact: (3.51, 54.20, -64.97) Vallecula: (--, 25.00, -33.00) Jaw width: 65.4mm Resolve speed: 0.01 Cycles: 95 Cycles at speed: 2 Auto: On Ghosts: Off Cache hit: No Results: 5 Camera position: (122.84, -6.51, 185.88) Camera target: (-3.00, -1.00, -72.00) Camera orientation: 7.4, 1.6, 287.0 ``` | intubation view   ```     Camera position: (21.59, 354.96, -206.85)     Camera target: (0.00, 0.00, 0.00)     Camera orientation: -7.8, 0.5, 411.4 ``` | side view   ```     Camera position: (313.86, 60.40, -60.35)     Camera target: (8.00, 94.00, -41.00)     Camera orientation: 6.2, 1.7, 308.3 ``` | top view   ```     Camera position: (11.49, 4.04, -386.21)     Camera target: (0.00, 110.00, 30.00)     Camera orientation: 4.7, 1.8, 429.6 ``` | above view   ```     Camera position: (-3.01, 324.55, -75.26)     Camera target: (-3.00, -1.00, -72.00)     Camera orientation: 4.7, 0.0, 325.6 ``` | glottis view   ```     Camera position: (125.12, -6.61, 190.56)     Camera target: (-3.00, -1.00, -72.00)     Camera orientation: 7.4, 1.6, 292.2 ``` |
| ELGent Yaw: 5.0° Roll: 30.0°   ``` Scenario: #6 (resolved) Subject: #0 ELGent Yaw: 5.0° Roll: 30.0° Tube lift: -12.933 mm Jaw contact X: 19.705 mm Jaw contact Y: 100.529 mm Tube position: (38.16, 93.51, -45.54) Tube rotation: (47.90, 5.00, 30.00) Upper incisor: (0.00, 110.00, -89.00) Lower incisor: (0.00, 102.00, -76.00) Glottic contact: (-1.36, -12.93, -0.12) Preglottic contact: (-1.93, -10.03, -10.00) Prepreglottic contact: (-2.72, -3.58, -20.00) Maxillary contact: (19.70, 100.53, -89.77) Blade contact: (3.94, 53.74, -64.73) Vallecula: (--, 25.00, -33.00) Jaw width: 65.4mm Resolve speed: 0.01 Cycles: 91 Cycles at speed: 1 Auto: On Ghosts: Off Cache hit: No Results: 6 Camera position: (127.71, -6.72, 195.85) Camera target: (-3.00, -1.00, -72.00) Camera orientation: 7.4, 1.6, 298.1 ``` | intubation view   ```     Camera position: (20.99, 345.12, -201.12)     Camera target: (0.00, 0.00, 0.00)     Camera orientation: -7.8, 0.5, 400.0 ``` | side view   ```     Camera position: (305.62, 61.30, -59.83)     Camera target: (8.00, 94.00, -41.00)     Camera orientation: 6.2, 1.7, 300.0 ``` | top view   ```     Camera position: (11.26, 6.17, -377.84)     Camera target: (0.00, 110.00, 30.00)     Camera orientation: 4.7, 1.8, 421.0 ``` | above view   ```     Camera position: (-3.01, 315.98, -75.17)     Camera target: (-3.00, -1.00, -72.00)     Camera orientation: 4.7, 0.0, 317.0 ``` | glottis view   ```     Camera position: (122.84, -6.51, 185.88)     Camera target: (-3.00, -1.00, -72.00)     Camera orientation: 7.4, 1.6, 287.0 ``` |
| ELGent Yaw: 5.0° Roll: 45.0°   ``` Scenario: #7 (resolved) Subject: #0 ELGent Yaw: 5.0° Roll: 45.0° Tube lift: -15.933 mm Jaw contact X: 24.113 mm Jaw contact Y: 94.472 mm Tube position: (52.05, 83.13, -46.84) Tube rotation: (48.34, 5.00, 45.00) Upper incisor: (0.00, 110.00, -89.00) Lower incisor: (0.00, 102.00, -76.00) Glottic contact: (0.49, -15.93, -0.12) Preglottic contact: (-0.54, -12.58, -10.00) Prepreglottic contact: (-2.11, -5.27, -20.00) Maxillary contact: (24.11, 94.47, -90.54) Blade contact: (5.09, 52.94, -64.31) Vallecula: (--, 25.00, -33.00) Jaw width: 65.4mm Resolve speed: 0.01 Cycles: 95 Cycles at speed: 2 Auto: On Ghosts: Off Cache hit: No Results: 7 Camera position: (122.84, -6.51, 185.88) Camera target: (-3.00, -1.00, -72.00) Camera orientation: 7.4, 1.6, 287.0 ``` | intubation view   ```     Camera position: (20.99, 345.12, -201.12)     Camera target: (0.00, 0.00, 0.00)     Camera orientation: -7.8, 0.5, 400.0 ``` | side view   ```     Camera position: (305.62, 61.30, -59.83)     Camera target: (8.00, 94.00, -41.00)     Camera orientation: 6.2, 1.7, 300.0 ``` | top view   ```     Camera position: (11.26, 6.17, -377.84)     Camera target: (0.00, 110.00, 30.00)     Camera orientation: 4.7, 1.8, 421.0 ``` | above view   ```     Camera position: (-3.01, 315.98, -75.17)     Camera target: (-3.00, -1.00, -72.00)     Camera orientation: 4.7, 0.0, 317.0 ``` | glottis view   ```     Camera position: (122.84, -6.51, 185.88)     Camera target: (-3.00, -1.00, -72.00)     Camera orientation: 7.4, 1.6, 287.0 ``` |
| ELGent Yaw: 10.0° Roll: 0.0°   ``` Scenario: #8 (resolved) Subject: #0 ELGent Yaw: 10.0° Roll: 0.0° Tube lift: -5.816 mm Jaw contact X: 19.426 mm Jaw contact Y: 100.917 mm Tube position: (13.77, 106.97, -45.66) Tube rotation: (46.36, 10.00, 0.00) Upper incisor: (0.00, 110.00, -89.00) Lower incisor: (0.00, 102.00, -76.00) Glottic contact: (-3.00, -5.82, -0.12) Preglottic contact: (-2.09, -3.44, -10.00) Prepreglottic contact: (-0.18, 2.00, -20.00) Maxillary contact: (19.43, 100.92, -89.73) Blade contact: (8.63, 49.90, -62.37) Vallecula: (--, 25.00, -33.00) Jaw width: 65.4mm Resolve speed: 0.01 Cycles: 91 Cycles at speed: 3 Auto: On Ghosts: Off Cache hit: No Results: 8 Camera position: (122.84, -6.51, 185.88) Camera target: (-3.00, -1.00, -72.00) Camera orientation: 7.4, 1.6, 287.0 ``` | intubation view   ```     Camera position: (21.70, 356.81, -207.93)     Camera target: (0.00, 0.00, 0.00)     Camera orientation: -7.8, 0.5, 413.6 ``` | side view   ```     Camera position: (316.81, 60.07, -60.54)     Camera target: (8.00, 94.00, -41.00)     Camera orientation: 6.2, 1.7, 311.3 ``` | top view   ```     Camera position: (11.48, 4.21, -385.52)     Camera target: (0.00, 110.00, 30.00)     Camera orientation: 4.7, 1.8, 428.9 ``` | above view   ```     Camera position: (-3.01, 320.20, -75.21)     Camera target: (-3.00, -1.00, -72.00)     Camera orientation: 4.7, 0.0, 321.2 ``` | glottis view   ```     Camera position: (123.96, -6.56, 188.19)     Camera target: (-3.00, -1.00, -72.00)     Camera orientation: 7.4, 1.6, 289.6 ``` |
| ELGent Yaw: 10.0° Roll: 15.0°   ``` Scenario: #9 (resolved) Subject: #0 ELGent Yaw: 10.0° Roll: 15.0° Tube lift: -5.934 mm Jaw contact X: 22.554 mm Jaw contact Y: 97.074 mm Tube position: (30.19, 103.16, -46.15) Tube rotation: (45.84, 10.00, 15.00) Upper incisor: (0.00, 110.00, -89.00) Lower incisor: (0.00, 102.00, -76.00) Glottic contact: (-3.01, -5.93, -0.12) Preglottic contact: (-2.61, -3.51, -10.00) Prepreglottic contact: (-1.60, 1.98, -20.00) Maxillary contact: (22.55, 97.07, -90.21) Blade contact: (8.23, 51.12, -63.30) Vallecula: (--, 25.00, -33.00) Jaw width: 65.4mm Resolve speed: 0.01 Cycles: 85 Cycles at speed: 2 Auto: On Ghosts: Off Cache hit: No Results: 9 Camera position: (125.23, -6.62, 190.78) Camera target: (-3.00, -1.00, -72.00) Camera orientation: 7.4, 1.6, 292.5 ``` | intubation view   ```     Camera position: (20.99, 345.12, -201.12)     Camera target: (0.00, 0.00, 0.00)     Camera orientation: -7.8, 0.5, 400.0 ``` | side view   ```     Camera position: (305.62, 61.30, -59.83)     Camera target: (8.00, 94.00, -41.00)     Camera orientation: 6.2, 1.7, 300.0 ``` | top view   ```     Camera position: (11.26, 6.17, -377.84)     Camera target: (0.00, 110.00, 30.00)     Camera orientation: 4.7, 1.8, 421.0 ``` | above view   ```     Camera position: (-3.01, 315.98, -75.17)     Camera target: (-3.00, -1.00, -72.00)     Camera orientation: 4.7, 0.0, 317.0 ``` | glottis view   ```     Camera position: (122.84, -6.51, 185.88)     Camera target: (-3.00, -1.00, -72.00)     Camera orientation: 7.4, 1.6, 287.0 ``` |
| ELGent Yaw: 10.0° Roll: 30.0°   ``` Scenario: #10 (resolved) Subject: #0 ELGent Yaw: 10.0° Roll: 30.0° Tube lift: -6.705 mm Jaw contact X: 25.035 mm Jaw contact Y: 92.869 mm Tube position: (45.37, 96.05, -46.98) Tube rotation: (45.64, 10.00, 30.00) Upper incisor: (0.00, 110.00, -89.00) Lower incisor: (0.00, 102.00, -76.00) Glottic contact: (-2.91, -6.71, -0.12) Preglottic contact: (-3.00, -4.06, -10.00) Prepreglottic contact: (-2.81, 1.83, -20.00) Maxillary contact: (25.03, 92.87, -90.73) Blade contact: (7.38, 51.40, -63.37) Vallecula: (--, 25.00, -33.00) Jaw width: 65.4mm Resolve speed: 0.01 Cycles: 96 Cycles at speed: 3 Auto: On Ghosts: Off Cache hit: No Results: 10 Camera position: (122.84, -6.51, 185.88) Camera target: (-3.00, -1.00, -72.00) Camera orientation: 7.4, 1.6, 287.0 ``` | intubation view   ```     Camera position: (20.99, 345.12, -201.12)     Camera target: (0.00, 0.00, 0.00)     Camera orientation: -7.8, 0.5, 400.0 ``` | side view   ```     Camera position: (305.62, 61.30, -59.83)     Camera target: (8.00, 94.00, -41.00)     Camera orientation: 6.2, 1.7, 300.0 ``` | top view   ```     Camera position: (11.26, 6.17, -377.84)     Camera target: (0.00, 110.00, 30.00)     Camera orientation: 4.7, 1.8, 421.0 ``` | above view   ```     Camera position: (-3.01, 315.98, -75.17)     Camera target: (-3.00, -1.00, -72.00)     Camera orientation: 4.7, 0.0, 317.0 ``` | glottis view   ```     Camera position: (122.84, -6.51, 185.88)     Camera target: (-3.00, -1.00, -72.00)     Camera orientation: 7.4, 1.6, 287.0 ``` |
| ELGent Yaw: 10.0° Roll: 45.0°   ``` Scenario: #11 (resolved) Subject: #0 ELGent Yaw: 10.0° Roll: 45.0° Tube lift: -8.465 mm Jaw contact X: 27.326 mm Jaw contact Y: 87.626 mm Tube position: (58.67, 85.61, -48.55) Tube rotation: (45.61, 10.00, 45.00) Upper incisor: (0.00, 110.00, -89.00) Lower incisor: (0.00, 102.00, -76.00) Glottic contact: (-1.96, -8.47, -0.12) Preglottic contact: (-2.47, -5.43, -10.00) Prepreglottic contact: (-2.98, 1.15, -20.00) Maxillary contact: (27.33, 87.63, -91.37) Blade contact: (6.98, 50.49, -62.71) Vallecula: (--, 25.00, -33.00) Jaw width: 65.4mm Resolve speed: 0.01 Cycles: 85 Cycles at speed: 1 Auto: On Ghosts: Off Cache hit: No Results: 11 Camera position: (122.84, -6.51, 185.88) Camera target: (-3.00, -1.00, -72.00) Camera orientation: 7.4, 1.6, 287.0 ``` | intubation view   ```     Camera position: (20.99, 345.12, -201.12)     Camera target: (0.00, 0.00, 0.00)     Camera orientation: -7.8, 0.5, 400.0 ``` | side view   ```     Camera position: (305.62, 61.30, -59.83)     Camera target: (8.00, 94.00, -41.00)     Camera orientation: 6.2, 1.7, 300.0 ``` | top view   ```     Camera position: (11.26, 6.17, -377.84)     Camera target: (0.00, 110.00, 30.00)     Camera orientation: 4.7, 1.8, 421.0 ``` | above view   ```     Camera position: (-3.01, 315.98, -75.17)     Camera target: (-3.00, -1.00, -72.00)     Camera orientation: 4.7, 0.0, 317.0 ``` | glottis view   ```     Camera position: (122.84, -6.51, 185.88)     Camera target: (-3.00, -1.00, -72.00)     Camera orientation: 7.4, 1.6, 287.0 ``` |
| ELGent Yaw: 15.0° Roll: 0.0°   ``` Scenario: #12 (resolved) Subject: #0 ELGent Yaw: 15.0° Roll: 0.0° Tube lift: 5.198 mm Jaw contact X: 26.278 mm Jaw contact Y: 90.313 mm Tube position: (21.99, 115.06, -48.99) Tube rotation: (42.39, 15.00, 0.00) Upper incisor: (0.00, 110.00, -89.00) Lower incisor: (0.00, 102.00, -76.00) Glottic contact: (-3.00, 5.20, -0.12) Preglottic contact: (-1.66, 7.12, -10.00) Prepreglottic contact: (1.12, 11.60, -20.00) Maxillary contact: (26.28, 90.31, -91.02) Blade contact: (6.45, 31.89, -42.33) Vallecula: (--, 25.00, -33.00) Jaw width: 65.4mm Resolve speed: 0.01 Cycles: 93 Cycles at speed: 1 Auto: On Ghosts: Off Cache hit: No Results: 12 Camera position: (109.70, -5.94, 158.95) Camera target: (-3.00, -1.00, -72.00) Camera orientation: 7.4, 1.6, 257.0 ``` | intubation view   ```     Camera position: (20.99, 345.12, -201.12)     Camera target: (0.00, 0.00, 0.00)     Camera orientation: -7.8, 0.5, 400.0 ``` | side view   ```     Camera position: (305.62, 61.30, -59.83)     Camera target: (8.00, 94.00, -41.00)     Camera orientation: 6.2, 1.7, 300.0 ``` | top view   ```     Camera position: (11.26, 6.17, -377.84)     Camera target: (0.00, 110.00, 30.00)     Camera orientation: 4.7, 1.8, 421.0 ``` | above view   ```     Camera position: (-3.01, 315.98, -75.17)     Camera target: (-3.00, -1.00, -72.00)     Camera orientation: 4.7, 0.0, 317.0 ``` | glottis view   ```     Camera position: (122.84, -6.51, 185.88)     Camera target: (-3.00, -1.00, -72.00)     Camera orientation: 7.4, 1.6, 287.0 ``` |
| ELGent Yaw: 15.0° Roll: 15.0°   ``` Scenario: #13 (resolved) Subject: #0 ELGent Yaw: 15.0° Roll: 15.0° Tube lift: 4.841 mm Jaw contact X: 27.722 mm Jaw contact Y: 86.814 mm Tube position: (38.10, 109.99, -49.37) Tube rotation: (41.63, 15.00, 15.00) Upper incisor: (0.00, 110.00, -89.00) Lower incisor: (0.00, 102.00, -76.00) Glottic contact: (-3.00, 4.84, -0.12) Preglottic contact: (-2.17, 6.81, -10.00) Prepreglottic contact: (-0.28, 11.31, -20.00) Maxillary contact: (27.72, 86.81, -91.45) Blade contact: (5.91, 34.99, -46.04) Vallecula: (--, 25.00, -33.00) Jaw width: 65.4mm Resolve speed: 0.01 Cycles: 86 Cycles at speed: 1 Auto: On Ghosts: Off Cache hit: No Results: 13 Camera position: (122.84, -6.51, 185.88) Camera target: (-3.00, -1.00, -72.00) Camera orientation: 7.4, 1.6, 287.0 ``` | intubation view   ```     Camera position: (20.99, 345.12, -201.12)     Camera target: (0.00, 0.00, 0.00)     Camera orientation: -7.8, 0.5, 400.0 ``` | side view   ```     Camera position: (305.62, 61.30, -59.83)     Camera target: (8.00, 94.00, -41.00)     Camera orientation: 6.2, 1.7, 300.0 ``` | top view   ```     Camera position: (11.26, 6.17, -377.84)     Camera target: (0.00, 110.00, 30.00)     Camera orientation: 4.7, 1.8, 421.0 ``` | above view   ```     Camera position: (-3.01, 315.98, -75.17)     Camera target: (-3.00, -1.00, -72.00)     Camera orientation: 4.7, 0.0, 317.0 ``` | glottis view   ```     Camera position: (122.84, -6.51, 185.88)     Camera target: (-3.00, -1.00, -72.00)     Camera orientation: 7.4, 1.6, 287.0 ``` |
| ELGent Yaw: 15.0° Roll: 30.0°   ``` Scenario: #14 (resolved) Subject: #0 ELGent Yaw: 15.0° Roll: 30.0° Tube lift: 3.107 mm Jaw contact X: 28.942 mm Jaw contact Y: 83.098 mm Tube position: (52.95, 100.78, -50.11) Tube rotation: (41.22, 15.00, 30.00) Upper incisor: (0.00, 110.00, -89.00) Lower incisor: (0.00, 102.00, -76.00) Glottic contact: (-3.00, 3.11, -0.12) Preglottic contact: (-2.63, 5.27, -10.00) Prepreglottic contact: (-1.53, 10.12, -20.00) Maxillary contact: (28.94, 83.10, -91.92) Blade contact: (8.02, 43.56, -56.74) Vallecula: (--, 25.00, -33.00) Jaw width: 65.4mm Resolve speed: 0.01 Cycles: 80 Cycles at speed: 1 Auto: On Ghosts: Off Cache hit: No Results: 14 Camera position: (122.84, -6.51, 185.88) Camera target: (-3.00, -1.00, -72.00) Camera orientation: 7.4, 1.6, 287.0 ``` | intubation view   ```     Camera position: (20.99, 345.12, -201.12)     Camera target: (0.00, 0.00, 0.00)     Camera orientation: -7.8, 0.5, 400.0 ``` | side view   ```     Camera position: (305.62, 61.30, -59.83)     Camera target: (8.00, 94.00, -41.00)     Camera orientation: 6.2, 1.7, 300.0 ``` | top view   ```     Camera position: (11.26, 6.17, -377.84)     Camera target: (0.00, 110.00, 30.00)     Camera orientation: 4.7, 1.8, 421.0 ``` | above view   ```     Camera position: (-3.01, 315.98, -75.17)     Camera target: (-3.00, -1.00, -72.00)     Camera orientation: 4.7, 0.0, 317.0 ``` | glottis view   ```     Camera position: (122.84, -6.51, 185.88)     Camera target: (-3.00, -1.00, -72.00)     Camera orientation: 7.4, 1.6, 287.0 ``` |
| ELGent Yaw: 15.0° Roll: 45.0°   ``` Scenario: #15 (resolved) Subject: #0 ELGent Yaw: 15.0° Roll: 45.0° Tube lift: 1.067 mm Jaw contact X: 29.9 mm Jaw contact Y: 79.513 mm Tube position: (65.55, 88.85, -51.48) Tube rotation: (41.07, 15.00, 45.00) Upper incisor: (0.00, 110.00, -89.00) Lower incisor: (0.00, 102.00, -76.00) Glottic contact: (-2.97, 1.07, -0.12) Preglottic contact: (-3.00, 3.58, -10.00) Prepreglottic contact: (-2.54, 9.06, -20.00) Maxillary contact: (29.90, 79.51, -92.37) Blade contact: (7.27, 44.50, -57.58) Vallecula: (--, 25.00, -33.00) Jaw width: 65.4mm Resolve speed: 0.01 Cycles: 83 Cycles at speed: 1 Auto: On Ghosts: Off Cache hit: No Results: 15 Camera position: (122.84, -6.51, 185.88) Camera target: (-3.00, -1.00, -72.00) Camera orientation: 7.4, 1.6, 287.0 ``` | intubation view   ```     Camera position: (20.99, 345.12, -201.12)     Camera target: (0.00, 0.00, 0.00)     Camera orientation: -7.8, 0.5, 400.0 ``` | side view   ```     Camera position: (305.62, 61.30, -59.83)     Camera target: (8.00, 94.00, -41.00)     Camera orientation: 6.2, 1.7, 300.0 ``` | top view   ```     Camera position: (11.26, 6.17, -377.84)     Camera target: (0.00, 110.00, 30.00)     Camera orientation: 4.7, 1.8, 421.0 ``` | above view   ```     Camera position: (-3.01, 315.98, -75.17)     Camera target: (-3.00, -1.00, -72.00)     Camera orientation: 4.7, 0.0, 317.0 ``` | glottis view   ```     Camera position: (122.84, -6.51, 185.88)     Camera target: (-3.00, -1.00, -72.00)     Camera orientation: 7.4, 1.6, 287.0 ``` |
| DLGent Yaw: 0.0° Roll: 0.0°   ``` Scenario: #0 (resolved) Subject: #1 DLGent Yaw: 0.0° Roll: 0.0° Tube lift: -16.249 mm Jaw contact X: -2.932 mm Jaw contact Y: 104.263 mm Tube position: (-3.00, 99.53, -37.08) Tube rotation: (55.10, 0.00, 0.00) Upper incisor: (0.00, 107.00, -74.00) Lower incisor: (0.00, 98.00, -60.00) Glottic contact: (-3.00, -16.25, -0.12) Preglottic contact: (-3.00, -12.77, -10.00) Prepreglottic contact: (-3.00, -4.96, -20.00) Maxillary contact: (-2.93, 104.26, -73.84) Blade contact: (-3.00, 59.13, -57.25) Vallecula: (--, 25.00, -33.00) Jaw width: 65.4mm Resolve speed: 0.01 Cycles: 96 Cycles at speed: 2 Auto: On Ghosts: Off Cache hit: No Results: 16 Camera position: (122.84, -6.51, 185.88) Camera target: (-3.00, -1.00, -72.00) Camera orientation: 7.4, 1.6, 287.0 ``` | intubation view   ```     Camera position: (20.99, 345.12, -201.12)     Camera target: (0.00, 0.00, 0.00)     Camera orientation: -7.8, 0.5, 400.0 ``` | side view   ```     Camera position: (305.62, 61.30, -59.83)     Camera target: (8.00, 94.00, -41.00)     Camera orientation: 6.2, 1.7, 300.0 ``` | top view   ```     Camera position: (11.26, 6.17, -377.84)     Camera target: (0.00, 110.00, 30.00)     Camera orientation: 4.7, 1.8, 421.0 ``` | above view   ```     Camera position: (-3.01, 315.98, -75.17)     Camera target: (-3.00, -1.00, -72.00)     Camera orientation: 4.7, 0.0, 317.0 ``` | glottis view   ```     Camera position: (122.84, -6.51, 185.88)     Camera target: (-3.00, -1.00, -72.00)     Camera orientation: 7.4, 1.6, 287.0 ``` |
| DLGent Yaw: 0.0° Roll: 15.0°   ``` Scenario: #1 (resolved) Subject: #1 DLGent Yaw: 0.0° Roll: 15.0° Tube lift: -19.566 mm Jaw contact X: 4.706 mm Jaw contact Y: 103.958 mm Tube position: (15.03, 94.99, -37.12) Tube rotation: (55.26, 0.00, 15.00) Upper incisor: (0.00, 107.00, -74.00) Lower incisor: (0.00, 98.00, -60.00) Glottic contact: (-0.30, -19.57, -0.12) Preglottic contact: (-0.86, -15.99, -10.00) Prepreglottic contact: (-1.84, -7.95, -20.00) Maxillary contact: (4.71, 103.96, -73.87) Blade contact: (-1.23, 59.03, -57.87) Vallecula: (--, 25.00, -33.00) Jaw width: 65.4mm Resolve speed: 0.01 Cycles: 95 Cycles at speed: 0 Auto: On Ghosts: Off Cache hit: No Results: 17 Camera position: (122.84, -6.51, 185.88) Camera target: (-3.00, -1.00, -72.00) Camera orientation: 7.4, 1.6, 287.0 ``` | intubation view   ```     Camera position: (20.99, 345.12, -201.12)     Camera target: (0.00, 0.00, 0.00)     Camera orientation: -7.8, 0.5, 400.0 ``` | side view   ```     Camera position: (305.62, 61.30, -59.83)     Camera target: (8.00, 94.00, -41.00)     Camera orientation: 6.2, 1.7, 300.0 ``` | top view   ```     Camera position: (11.26, 6.17, -377.84)     Camera target: (0.00, 110.00, 30.00)     Camera orientation: 4.7, 1.8, 421.0 ``` | above view   ```     Camera position: (-3.01, 315.98, -75.17)     Camera target: (-3.00, -1.00, -72.00)     Camera orientation: 4.7, 0.0, 317.0 ``` | glottis view   ```     Camera position: (122.84, -6.51, 185.88)     Camera target: (-3.00, -1.00, -72.00)     Camera orientation: 7.4, 1.6, 287.0 ``` |
| DLGent Yaw: 0.0° Roll: 30.0°   ``` Scenario: #2 (resolved) Subject: #1 DLGent Yaw: 0.0° Roll: 30.0° Tube lift: -22.637 mm Jaw contact X: 12.078 mm Jaw contact Y: 102.241 mm Tube position: (31.66, 88.43, -37.21) Tube rotation: (55.85, 0.00, 30.00) Upper incisor: (0.00, 107.00, -74.00) Lower incisor: (0.00, 98.00, -60.00) Glottic contact: (2.23, -22.64, -0.12) Preglottic contact: (1.09, -18.70, -10.00) Prepreglottic contact: (-0.86, -9.93, -20.00) Maxillary contact: (12.08, 102.24, -74.09) Blade contact: (0.80, 59.07, -57.42) Vallecula: (--, 25.00, -33.00) Jaw width: 65.4mm Resolve speed: 0.01 Cycles: 72 Cycles at speed: 3 Auto: On Ghosts: Off Cache hit: No Results: 18 Camera position: (122.84, -6.51, 185.88) Camera target: (-3.00, -1.00, -72.00) Camera orientation: 7.4, 1.6, 287.0 ``` | intubation view   ```     Camera position: (20.99, 345.12, -201.12)     Camera target: (0.00, 0.00, 0.00)     Camera orientation: -7.8, 0.5, 400.0 ``` | side view   ```     Camera position: (305.62, 61.30, -59.83)     Camera target: (8.00, 94.00, -41.00)     Camera orientation: 6.2, 1.7, 300.0 ``` | top view   ```     Camera position: (11.26, 6.17, -377.84)     Camera target: (0.00, 110.00, 30.00)     Camera orientation: 4.7, 1.8, 421.0 ``` | above view   ```     Camera position: (-3.01, 315.98, -75.17)     Camera target: (-3.00, -1.00, -72.00)     Camera orientation: 4.7, 0.0, 317.0 ``` | glottis view   ```     Camera position: (122.84, -6.51, 185.88)     Camera target: (-3.00, -1.00, -72.00)     Camera orientation: 7.4, 1.6, 287.0 ``` |
| DLGent Yaw: 0.0° Roll: 45.0°   ``` Scenario: #3 (resolved) Subject: #1 DLGent Yaw: 0.0° Roll: 45.0° Tube lift: -22.547 mm Jaw contact X: 16.948 mm Jaw contact Y: 99.983 mm Tube position: (45.76, 83.11, -37.63) Tube rotation: (56.81, 0.00, 45.00) Upper incisor: (0.00, 107.00, -74.00) Lower incisor: (0.00, 98.00, -60.00) Glottic contact: (4.40, -22.55, -0.12) Preglottic contact: (2.68, -17.98, -10.00) Prepreglottic contact: (-0.22, -7.96, -20.00) Maxillary contact: (16.95, 99.98, -74.46) Blade contact: (2.24, 59.26, -54.57) Vallecula: (--, 25.00, -33.00) Jaw width: 65.4mm Resolve speed: 0.01 Cycles: 82 Cycles at speed: 3 Auto: On Ghosts: Off Cache hit: No Results: 19 Camera position: (122.84, -6.51, 185.88) Camera target: (-3.00, -1.00, -72.00) Camera orientation: 7.4, 1.6, 287.0 ``` | intubation view   ```     Camera position: (20.99, 345.12, -201.12)     Camera target: (0.00, 0.00, 0.00)     Camera orientation: -7.8, 0.5, 400.0 ``` | side view   ```     Camera position: (305.62, 61.30, -59.83)     Camera target: (8.00, 94.00, -41.00)     Camera orientation: 6.2, 1.7, 300.0 ``` | top view   ```     Camera position: (11.26, 6.17, -377.84)     Camera target: (0.00, 110.00, 30.00)     Camera orientation: 4.7, 1.8, 421.0 ``` | above view   ```     Camera position: (-3.01, 315.98, -75.17)     Camera target: (-3.00, -1.00, -72.00)     Camera orientation: 4.7, 0.0, 317.0 ``` | glottis view   ```     Camera position: (122.84, -6.51, 185.88)     Camera target: (-3.00, -1.00, -72.00)     Camera orientation: 7.4, 1.6, 287.0 ``` |
| DLGent Yaw: 5.0° Roll: 0.0°   ``` Scenario: #4 (resolved) Subject: #1 DLGent Yaw: 5.0° Roll: 0.0° Tube lift: -17.486 mm Jaw contact X: 8.853 mm Jaw contact Y: 103.164 mm Tube position: (5.42, 97.97, -37.16) Tube rotation: (54.88, 5.00, 0.00) Upper incisor: (0.00, 107.00, -74.00) Lower incisor: (0.00, 98.00, -60.00) Glottic contact: (-3.00, -17.49, -0.12) Preglottic contact: (-2.51, -14.05, -10.00) Prepreglottic contact: (-1.45, -6.32, -20.00) Maxillary contact: (8.85, 103.16, -73.97) Blade contact: (5.30, 58.78, -57.78) Vallecula: (--, 25.00, -33.00) Jaw width: 65.4mm Resolve speed: 0.01 Cycles: 104 Cycles at speed: 3 Auto: On Ghosts: Off Cache hit: No Results: 20 Camera position: (122.84, -6.51, 185.88) Camera target: (-3.00, -1.00, -72.00) Camera orientation: 7.4, 1.6, 287.0 ``` | intubation view   ```     Camera position: (20.99, 345.12, -201.12)     Camera target: (0.00, 0.00, 0.00)     Camera orientation: -7.8, 0.5, 400.0 ``` | side view   ```     Camera position: (305.62, 61.30, -59.83)     Camera target: (8.00, 94.00, -41.00)     Camera orientation: 6.2, 1.7, 300.0 ``` | top view   ```     Camera position: (11.26, 6.17, -377.84)     Camera target: (0.00, 110.00, 30.00)     Camera orientation: 4.7, 1.8, 421.0 ``` | above view   ```     Camera position: (-3.01, 315.98, -75.17)     Camera target: (-3.00, -1.00, -72.00)     Camera orientation: 4.7, 0.0, 317.0 ``` | glottis view   ```     Camera position: (122.84, -6.51, 185.88)     Camera target: (-3.00, -1.00, -72.00)     Camera orientation: 7.4, 1.6, 287.0 ``` |
| DLGent Yaw: 5.0° Roll: 15.0°   ``` Scenario: #5 (resolved) Subject: #1 DLGent Yaw: 5.0° Roll: 15.0° Tube lift: -17.902 mm Jaw contact X: 13.232 mm Jaw contact Y: 101.514 mm Tube position: (22.06, 95.15, -37.26) Tube rotation: (54.89, 5.00, 15.00) Upper incisor: (0.00, 107.00, -74.00) Lower incisor: (0.00, 98.00, -60.00) Glottic contact: (-2.93, -17.90, -0.12) Preglottic contact: (-3.00, -14.37, -10.00) Prepreglottic contact: (-2.90, -6.43, -20.00) Maxillary contact: (13.23, 101.51, -74.17) Blade contact: (4.30, 58.57, -57.71) Vallecula: (--, 25.00, -33.00) Jaw width: 65.4mm Resolve speed: 0.01 Cycles: 98 Cycles at speed: 2 Auto: On Ghosts: Off Cache hit: No Results: 21 Camera position: (127.22, -6.70, 194.85) Camera target: (-3.00, -1.00, -72.00) Camera orientation: 7.4, 1.6, 297.0 ``` | intubation view   ```     Camera position: (20.99, 345.12, -201.12)     Camera target: (0.00, 0.00, 0.00)     Camera orientation: -7.8, 0.5, 400.0 ``` | side view   ```     Camera position: (305.62, 61.30, -59.83)     Camera target: (8.00, 94.00, -41.00)     Camera orientation: 6.2, 1.7, 300.0 ``` | top view   ```     Camera position: (11.26, 6.17, -377.84)     Camera target: (0.00, 110.00, 30.00)     Camera orientation: 4.7, 1.8, 421.0 ``` | above view   ```     Camera position: (-3.01, 315.98, -75.17)     Camera target: (-3.00, -1.00, -72.00)     Camera orientation: 4.7, 0.0, 317.0 ``` | glottis view   ```     Camera position: (122.84, -6.51, 185.88)     Camera target: (-3.00, -1.00, -72.00)     Camera orientation: 7.4, 1.6, 287.0 ``` |
| DLGent Yaw: 5.0° Roll: 30.0°   ``` Scenario: #6 (resolved) Subject: #1 DLGent Yaw: 5.0° Roll: 30.0° Tube lift: -20.596 mm Jaw contact X: 18.478 mm Jaw contact Y: 98.149 mm Tube position: (38.16, 87.78, -37.51) Tube rotation: (55.22, 5.00, 30.00) Upper incisor: (0.00, 107.00, -74.00) Lower incisor: (0.00, 98.00, -60.00) Glottic contact: (-1.36, -20.60, -0.12) Preglottic contact: (-1.97, -16.74, -10.00) Prepreglottic contact: (-2.79, -8.15, -20.00) Maxillary contact: (18.48, 98.15, -74.63) Blade contact: (5.30, 58.32, -57.62) Vallecula: (--, 25.00, -33.00) Jaw width: 65.4mm Resolve speed: 0.01 Cycles: 90 Cycles at speed: 0 Auto: On Ghosts: Off Cache hit: No Results: 22 Camera position: (122.84, -6.51, 185.88) Camera target: (-3.00, -1.00, -72.00) Camera orientation: 7.4, 1.6, 287.0 ``` | intubation view   ```     Camera position: (20.99, 345.12, -201.12)     Camera target: (0.00, 0.00, 0.00)     Camera orientation: -7.8, 0.5, 400.0 ``` | side view   ```     Camera position: (305.62, 61.30, -59.83)     Camera target: (8.00, 94.00, -41.00)     Camera orientation: 6.2, 1.7, 300.0 ``` | top view   ```     Camera position: (11.26, 6.17, -377.84)     Camera target: (0.00, 110.00, 30.00)     Camera orientation: 4.7, 1.8, 421.0 ``` | above view   ```     Camera position: (-3.01, 315.98, -75.17)     Camera target: (-3.00, -1.00, -72.00)     Camera orientation: 4.7, 0.0, 317.0 ``` | glottis view   ```     Camera position: (122.84, -6.51, 185.88)     Camera target: (-3.00, -1.00, -72.00)     Camera orientation: 7.4, 1.6, 287.0 ``` |
| DLGent Yaw: 5.0° Roll: 45.0°   ``` Scenario: #7 (resolved) Subject: #1 DLGent Yaw: 5.0° Roll: 45.0° Tube lift: -22.634 mm Jaw contact X: 22.482 mm Jaw contact Y: 93.748 mm Tube position: (52.05, 79.20, -38.19) Tube rotation: (55.81, 5.00, 45.00) Upper incisor: (0.00, 107.00, -74.00) Lower incisor: (0.00, 98.00, -60.00) Glottic contact: (0.48, -22.63, -0.12) Preglottic contact: (-0.64, -18.19, -10.00) Prepreglottic contact: (-2.30, -8.49, -20.00) Maxillary contact: (22.48, 93.75, -75.23) Blade contact: (5.47, 57.16, -56.17) Vallecula: (--, 25.00, -33.00) Jaw width: 65.4mm Resolve speed: 0.01 Cycles: 78 Cycles at speed: 3 Auto: On Ghosts: Off Cache hit: No Results: 23 Camera position: (122.84, -6.51, 185.88) Camera target: (-3.00, -1.00, -72.00) Camera orientation: 7.4, 1.6, 287.0 ``` | intubation view   ```     Camera position: (20.99, 345.12, -201.12)     Camera target: (0.00, 0.00, 0.00)     Camera orientation: -7.8, 0.5, 400.0 ``` | side view   ```     Camera position: (305.62, 61.30, -59.83)     Camera target: (8.00, 94.00, -41.00)     Camera orientation: 6.2, 1.7, 300.0 ``` | top view   ```     Camera position: (11.26, 6.17, -377.84)     Camera target: (0.00, 110.00, 30.00)     Camera orientation: 4.7, 1.8, 421.0 ``` | above view   ```     Camera position: (-3.01, 315.98, -75.17)     Camera target: (-3.00, -1.00, -72.00)     Camera orientation: 4.7, 0.0, 317.0 ``` | glottis view   ```     Camera position: (122.84, -6.51, 185.88)     Camera target: (-3.00, -1.00, -72.00)     Camera orientation: 7.4, 1.6, 287.0 ``` |
| DLGent Yaw: 10.0° Roll: 0.0°   ``` Scenario: #8 (resolved) Subject: #1 DLGent Yaw: 10.0° Roll: 0.0° Tube lift: -11.436 mm Jaw contact X: 18.82 mm Jaw contact Y: 98.249 mm Tube position: (13.77, 103.06, -37.59) Tube rotation: (54.02, 10.00, 0.00) Upper incisor: (0.00, 107.00, -74.00) Lower incisor: (0.00, 98.00, -60.00) Glottic contact: (-3.00, -11.44, -0.12) Preglottic contact: (-2.02, -8.14, -10.00) Prepreglottic contact: (0.07, -0.70, -20.00) Maxillary contact: (18.82, 98.25, -74.65) Blade contact: (8.69, 52.64, -55.11) Vallecula: (--, 25.00, -33.00) Jaw width: 65.4mm Resolve speed: 0.01 Cycles: 86 Cycles at speed: 0 Auto: On Ghosts: Off Cache hit: No Results: 24 Camera position: (122.84, -6.51, 185.88) Camera target: (-3.00, -1.00, -72.00) Camera orientation: 7.4, 1.6, 287.0 ``` | intubation view   ```     Camera position: (20.99, 345.12, -201.12)     Camera target: (0.00, 0.00, 0.00)     Camera orientation: -7.8, 0.5, 400.0 ``` | side view   ```     Camera position: (305.62, 61.30, -59.83)     Camera target: (8.00, 94.00, -41.00)     Camera orientation: 6.2, 1.7, 300.0 ``` | top view   ```     Camera position: (11.26, 6.17, -377.84)     Camera target: (0.00, 110.00, 30.00)     Camera orientation: 4.7, 1.8, 421.0 ``` | above view   ```     Camera position: (-3.01, 315.98, -75.17)     Camera target: (-3.00, -1.00, -72.00)     Camera orientation: 4.7, 0.0, 317.0 ``` | glottis view   ```     Camera position: (122.84, -6.51, 185.88)     Camera target: (-3.00, -1.00, -72.00)     Camera orientation: 7.4, 1.6, 287.0 ``` |
| DLGent Yaw: 10.0° Roll: 15.0°   ``` Scenario: #9 (resolved) Subject: #1 DLGent Yaw: 10.0° Roll: 15.0° Tube lift: -12.688 mm Jaw contact X: 21.471 mm Jaw contact Y: 95.278 mm Tube position: (30.19, 98.23, -37.82) Tube rotation: (53.79, 10.00, 15.00) Upper incisor: (0.00, 107.00, -74.00) Lower incisor: (0.00, 98.00, -60.00) Glottic contact: (-3.00, -12.69, -0.12) Preglottic contact: (-2.57, -9.30, -10.00) Prepreglottic contact: (-1.43, -1.72, -20.00) Maxillary contact: (21.47, 95.28, -75.02) Blade contact: (8.43, 54.36, -56.07) Vallecula: (--, 25.00, -33.00) Jaw width: 65.4mm Resolve speed: 0.01 Cycles: 98 Cycles at speed: 2 Auto: On Ghosts: Off Cache hit: No Results: 25 Camera position: (122.84, -6.51, 185.88) Camera target: (-3.00, -1.00, -72.00) Camera orientation: 7.4, 1.6, 287.0 ``` | intubation view   ```     Camera position: (20.99, 345.12, -201.12)     Camera target: (0.00, 0.00, 0.00)     Camera orientation: -7.8, 0.5, 400.0 ``` | side view   ```     Camera position: (305.62, 61.30, -59.83)     Camera target: (8.00, 94.00, -41.00)     Camera orientation: 6.2, 1.7, 300.0 ``` | top view   ```     Camera position: (11.26, 6.17, -377.84)     Camera target: (0.00, 110.00, 30.00)     Camera orientation: 4.7, 1.8, 421.0 ``` | above view   ```     Camera position: (-3.01, 315.98, -75.17)     Camera target: (-3.00, -1.00, -72.00)     Camera orientation: 4.7, 0.0, 317.0 ``` | glottis view   ```     Camera position: (122.84, -6.51, 185.88)     Camera target: (-3.00, -1.00, -72.00)     Camera orientation: 7.4, 1.6, 287.0 ``` |
| DLGent Yaw: 10.0° Roll: 30.0°   ``` Scenario: #10 (resolved) Subject: #1 DLGent Yaw: 10.0° Roll: 30.0° Tube lift: -14.804 mm Jaw contact X: 23.828 mm Jaw contact Y: 91.778 mm Tube position: (45.37, 90.25, -38.26) Tube rotation: (53.85, 10.00, 30.00) Upper incisor: (0.00, 107.00, -74.00) Lower incisor: (0.00, 98.00, -60.00) Glottic contact: (-2.91, -14.80, -0.12) Preglottic contact: (-3.00, -11.13, -10.00) Prepreglottic contact: (-2.72, -2.98, -20.00) Maxillary contact: (23.83, 91.78, -75.47) Blade contact: (8.11, 54.92, -56.39) Vallecula: (--, 25.00, -33.00) Jaw width: 65.4mm Resolve speed: 0.01 Cycles: 89 Cycles at speed: 1 Auto: On Ghosts: Off Cache hit: No Results: 26 Camera position: (122.84, -6.51, 185.88) Camera target: (-3.00, -1.00, -72.00) Camera orientation: 7.4, 1.6, 287.0 ``` | intubation view   ```     Camera position: (20.99, 345.12, -201.12)     Camera target: (0.00, 0.00, 0.00)     Camera orientation: -7.8, 0.5, 400.0 ``` | side view   ```     Camera position: (305.62, 61.30, -59.83)     Camera target: (8.00, 94.00, -41.00)     Camera orientation: 6.2, 1.7, 300.0 ``` | top view   ```     Camera position: (11.26, 6.17, -377.84)     Camera target: (0.00, 110.00, 30.00)     Camera orientation: 4.7, 1.8, 421.0 ``` | above view   ```     Camera position: (-3.01, 315.98, -75.17)     Camera target: (-3.00, -1.00, -72.00)     Camera orientation: 4.7, 0.0, 317.0 ``` | glottis view   ```     Camera position: (122.84, -6.51, 185.88)     Camera target: (-3.00, -1.00, -72.00)     Camera orientation: 7.4, 1.6, 287.0 ``` |
| DLGent Yaw: 10.0° Roll: 45.0°   ``` Scenario: #11 (resolved) Subject: #1 DLGent Yaw: 10.0° Roll: 45.0° Tube lift: -17.547 mm Jaw contact X: 26.2 mm Jaw contact Y: 87.167 mm Tube position: (58.67, 79.75, -39.21) Tube rotation: (54.06, 10.00, 45.00) Upper incisor: (0.00, 107.00, -74.00) Lower incisor: (0.00, 98.00, -60.00) Glottic contact: (-1.96, -17.55, -0.12) Preglottic contact: (-2.51, -13.36, -10.00) Prepreglottic contact: (-2.99, -4.25, -20.00) Maxillary contact: (26.20, 87.17, -76.05) Blade contact: (7.97, 53.59, -55.75) Vallecula: (--, 25.00, -33.00) Jaw width: 65.4mm Resolve speed: 0.01 Cycles: 94 Cycles at speed: 1 Auto: On Ghosts: Off Cache hit: No Results: 27 Camera position: (122.84, -6.51, 185.88) Camera target: (-3.00, -1.00, -72.00) Camera orientation: 7.4, 1.6, 287.0 ``` | intubation view   ```     Camera position: (20.99, 345.12, -201.12)     Camera target: (0.00, 0.00, 0.00)     Camera orientation: -7.8, 0.5, 400.0 ``` | side view   ```     Camera position: (305.62, 61.30, -59.83)     Camera target: (8.00, 94.00, -41.00)     Camera orientation: 6.2, 1.7, 300.0 ``` | top view   ```     Camera position: (11.26, 6.17, -377.84)     Camera target: (0.00, 110.00, 30.00)     Camera orientation: 4.7, 1.8, 421.0 ``` | above view   ```     Camera position: (-3.01, 315.98, -75.17)     Camera target: (-3.00, -1.00, -72.00)     Camera orientation: 4.7, 0.0, 317.0 ``` | glottis view   ```     Camera position: (122.84, -6.51, 185.88)     Camera target: (-3.00, -1.00, -72.00)     Camera orientation: 7.4, 1.6, 287.0 ``` |
| DLGent Yaw: 15.0° Roll: 0.0°   ``` Scenario: #12 (resolved) Subject: #1 DLGent Yaw: 15.0° Roll: 0.0° Tube lift: -2.406 mm Jaw contact X: 25.525 mm Jaw contact Y: 88.895 mm Tube position: (21.99, 110.32, -39.40) Tube rotation: (51.46, 15.00, 0.00) Upper incisor: (0.00, 107.00, -74.00) Lower incisor: (0.00, 98.00, -60.00) Glottic contact: (-3.00, -2.41, -0.12) Preglottic contact: (-1.56, 0.52, -10.00) Prepreglottic contact: (1.50, 7.17, -20.00) Maxillary contact: (25.53, 88.90, -75.83) Blade contact: (7.11, 25.04, -33.07) Vallecula: (--, 25.00, -33.00) Jaw width: 65.4mm Resolve speed: 0.01 Cycles: 92 Cycles at speed: 3 Auto: On Ghosts: Off Cache hit: No Results: 28 Camera position: (122.84, -6.51, 185.88) Camera target: (-3.00, -1.00, -72.00) Camera orientation: 7.4, 1.6, 287.0 ``` | intubation view   ```     Camera position: (20.99, 345.12, -201.12)     Camera target: (0.00, 0.00, 0.00)     Camera orientation: -7.8, 0.5, 400.0 ``` | side view   ```     Camera position: (305.62, 61.30, -59.83)     Camera target: (8.00, 94.00, -41.00)     Camera orientation: 6.2, 1.7, 300.0 ``` | top view   ```     Camera position: (11.26, 6.17, -377.84)     Camera target: (0.00, 110.00, 30.00)     Camera orientation: 4.7, 1.8, 421.0 ``` | above view   ```     Camera position: (-3.01, 315.98, -75.17)     Camera target: (-3.00, -1.00, -72.00)     Camera orientation: 4.7, 0.0, 317.0 ``` | glottis view   ```     Camera position: (122.84, -6.51, 185.88)     Camera target: (-3.00, -1.00, -72.00)     Camera orientation: 7.4, 1.6, 287.0 ``` |
| DLGent Yaw: 15.0° Roll: 15.0°   ``` Scenario: #13 (resolved) Subject: #1 DLGent Yaw: 15.0° Roll: 15.0° Tube lift: -2.662 mm Jaw contact X: 26.469 mm Jaw contact Y: 86.85 mm Tube position: (38.10, 105.39, -39.58) Tube rotation: (51.15, 15.00, 15.00) Upper incisor: (0.00, 107.00, -74.00) Lower incisor: (0.00, 98.00, -60.00) Glottic contact: (-3.00, -2.66, -0.12) Preglottic contact: (-2.10, 0.36, -10.00) Prepreglottic contact: (0.02, 7.15, -20.00) Maxillary contact: (26.47, 86.85, -76.08) Blade contact: (5.84, 35.06, -41.73) Vallecula: (--, 25.00, -33.00) Jaw width: 65.4mm Resolve speed: 0.01 Cycles: 93 Cycles at speed: 3 Auto: On Ghosts: Off Cache hit: No Results: 29 Camera position: (122.84, -6.51, 185.88) Camera target: (-3.00, -1.00, -72.00) Camera orientation: 7.4, 1.6, 287.0 ``` | intubation view   ```     Camera position: (20.99, 345.12, -201.12)     Camera target: (0.00, 0.00, 0.00)     Camera orientation: -7.8, 0.5, 400.0 ``` | side view   ```     Camera position: (305.62, 61.30, -59.83)     Camera target: (8.00, 94.00, -41.00)     Camera orientation: 6.2, 1.7, 300.0 ``` | top view   ```     Camera position: (11.26, 6.17, -377.84)     Camera target: (0.00, 110.00, 30.00)     Camera orientation: 4.7, 1.8, 421.0 ``` | above view   ```     Camera position: (-3.01, 315.98, -75.17)     Camera target: (-3.00, -1.00, -72.00)     Camera orientation: 4.7, 0.0, 317.0 ``` | glottis view   ```     Camera position: (122.84, -6.51, 185.88)     Camera target: (-3.00, -1.00, -72.00)     Camera orientation: 7.4, 1.6, 287.0 ``` |
| DLGent Yaw: 15.0° Roll: 30.0°   ``` Scenario: #14 (resolved) Subject: #1 DLGent Yaw: 15.0° Roll: 30.0° Tube lift: -5.022 mm Jaw contact X: 27.53 mm Jaw contact Y: 84.15 mm Tube position: (52.95, 95.98, -40.03) Tube rotation: (51.06, 15.00, 30.00) Upper incisor: (0.00, 107.00, -74.00) Lower incisor: (0.00, 98.00, -60.00) Glottic contact: (-3.00, -5.02, -0.12) Preglottic contact: (-2.59, -1.72, -10.00) Prepreglottic contact: (-1.30, 5.57, -20.00) Maxillary contact: (27.53, 84.15, -76.42) Blade contact: (8.22, 44.73, -49.93) Vallecula: (--, 25.00, -33.00) Jaw width: 65.4mm Resolve speed: 0.01 Cycles: 82 Cycles at speed: 1 Auto: On Ghosts: Off Cache hit: No Results: 30 Camera position: (122.84, -6.51, 185.88) Camera target: (-3.00, -1.00, -72.00) Camera orientation: 7.4, 1.6, 287.0 ``` | intubation view   ```     Camera position: (20.99, 345.12, -201.12)     Camera target: (0.00, 0.00, 0.00)     Camera orientation: -7.8, 0.5, 400.0 ``` | side view   ```     Camera position: (305.62, 61.30, -59.83)     Camera target: (8.00, 94.00, -41.00)     Camera orientation: 6.2, 1.7, 300.0 ``` | top view   ```     Camera position: (11.26, 6.17, -377.84)     Camera target: (0.00, 110.00, 30.00)     Camera orientation: 4.7, 1.8, 421.0 ``` | above view   ```     Camera position: (-3.01, 315.98, -75.17)     Camera target: (-3.00, -1.00, -72.00)     Camera orientation: 4.7, 0.0, 317.0 ``` | glottis view   ```     Camera position: (122.84, -6.51, 185.88)     Camera target: (-3.00, -1.00, -72.00)     Camera orientation: 7.4, 1.6, 287.0 ``` |
| DLGent Yaw: 15.0° Roll: 45.0°   ``` Scenario: #15 (resolved) Subject: #1 DLGent Yaw: 15.0° Roll: 45.0° Tube lift: -8.217 mm Jaw contact X: 28.613 mm Jaw contact Y: 80.913 mm Tube position: (65.55, 83.79, -41.01) Tube rotation: (51.13, 15.00, 45.00) Upper incisor: (0.00, 107.00, -74.00) Lower incisor: (0.00, 98.00, -60.00) Glottic contact: (-2.97, -8.22, -0.12) Preglottic contact: (-2.99, -4.45, -10.00) Prepreglottic contact: (-2.38, 3.70, -20.00) Maxillary contact: (28.61, 80.91, -76.82) Blade contact: (7.97, 46.21, -51.06) Vallecula: (--, 25.00, -33.00) Jaw width: 65.4mm Resolve speed: 0.01 Cycles: 87 Cycles at speed: 1 Auto: On Ghosts: Off Cache hit: No Results: 31 Camera position: (122.84, -6.51, 185.88) Camera target: (-3.00, -1.00, -72.00) Camera orientation: 7.4, 1.6, 287.0 ``` | intubation view   ```     Camera position: (20.99, 345.12, -201.12)     Camera target: (0.00, 0.00, 0.00)     Camera orientation: -7.8, 0.5, 400.0 ``` | side view   ```     Camera position: (305.62, 61.30, -59.83)     Camera target: (8.00, 94.00, -41.00)     Camera orientation: 6.2, 1.7, 300.0 ``` | top view   ```     Camera position: (11.26, 6.17, -377.84)     Camera target: (0.00, 110.00, 30.00)     Camera orientation: 4.7, 1.8, 421.0 ``` | above view   ```     Camera position: (-3.01, 315.98, -75.17)     Camera target: (-3.00, -1.00, -72.00)     Camera orientation: 4.7, 0.0, 317.0 ``` | glottis view   ```     Camera position: (122.84, -6.51, 185.88)     Camera target: (-3.00, -1.00, -72.00)     Camera orientation: 7.4, 1.6, 287.0 ``` |
| EL50N Yaw: 0.0° Roll: 0.0°   ``` Scenario: #0 (resolved) Subject: #2 EL50N Yaw: 0.0° Roll: 0.0° Tube lift: -0.449 mm Jaw contact X: -2.788 mm Jaw contact Y: 101.038 mm Tube position: (-3.00, 111.34, -51.59) Tube rotation: (41.84, 0.00, 0.00) Upper incisor: (0.00, 102.00, -100.00) Lower incisor: (0.00, 100.00, -82.00) Glottic contact: (-3.00, -0.45, -0.12) Preglottic contact: (-3.00, 1.48, -10.00) Prepreglottic contact: (-3.00, 5.94, -20.00) Maxillary contact: (-2.79, 101.04, -99.79) Blade contact: (-3.00, 49.70, -65.40) Vallecula: (--, 25.00, -33.00) Jaw width: 65.4mm Resolve speed: 0.01 Cycles: 99 Cycles at speed: 3 Auto: On Ghosts: Off Cache hit: No Results: 32 Camera position: (122.84, -6.51, 185.88) Camera target: (-3.00, -1.00, -72.00) Camera orientation: 7.4, 1.6, 287.0 ``` | intubation view   ```     Camera position: (20.99, 345.12, -201.12)     Camera target: (0.00, 0.00, 0.00)     Camera orientation: -7.8, 0.5, 400.0 ``` | side view   ```     Camera position: (305.62, 61.30, -59.83)     Camera target: (8.00, 94.00, -41.00)     Camera orientation: 6.2, 1.7, 300.0 ``` | top view   ```     Camera position: (11.26, 6.17, -377.84)     Camera target: (0.00, 110.00, 30.00)     Camera orientation: 4.7, 1.8, 421.0 ``` | above view   ```     Camera position: (-3.01, 315.98, -75.17)     Camera target: (-3.00, -1.00, -72.00)     Camera orientation: 4.7, 0.0, 317.0 ``` | glottis view   ```     Camera position: (122.84, -6.51, 185.88)     Camera target: (-3.00, -1.00, -72.00)     Camera orientation: 7.4, 1.6, 287.0 ``` |
| EL50N Yaw: 0.0° Roll: 15.0°   ``` Scenario: #1 (resolved) Subject: #2 EL50N Yaw: 0.0° Roll: 15.0° Tube lift: -3.053 mm Jaw contact X: 4.081 mm Jaw contact Y: 100.887 mm Tube position: (15.03, 107.41, -51.48) Tube rotation: (42.29, 0.00, 15.00) Upper incisor: (0.00, 102.00, -100.00) Lower incisor: (0.00, 100.00, -82.00) Glottic contact: (-0.29, -3.05, -0.12) Preglottic contact: (-0.80, -1.01, -10.00) Prepreglottic contact: (-1.67, 3.68, -20.00) Maxillary contact: (4.08, 100.89, -99.80) Blade contact: (-2.25, 49.53, -66.16) Vallecula: (--, 25.00, -33.00) Jaw width: 65.4mm Resolve speed: 0.01 Cycles: 96 Cycles at speed: 3 Auto: On Ghosts: Off Cache hit: No Results: 33 Camera position: (122.84, -6.51, 185.88) Camera target: (-3.00, -1.00, -72.00) Camera orientation: 7.4, 1.6, 287.0 ``` | intubation view   ```     Camera position: (20.99, 345.12, -201.12)     Camera target: (0.00, 0.00, 0.00)     Camera orientation: -7.8, 0.5, 400.0 ``` | side view   ```     Camera position: (305.62, 61.30, -59.83)     Camera target: (8.00, 94.00, -41.00)     Camera orientation: 6.2, 1.7, 300.0 ``` | top view   ```     Camera position: (11.26, 6.17, -377.84)     Camera target: (0.00, 110.00, 30.00)     Camera orientation: 4.7, 1.8, 421.0 ``` | above view   ```     Camera position: (-3.01, 315.98, -75.17)     Camera target: (-3.00, -1.00, -72.00)     Camera orientation: 4.7, 0.0, 317.0 ``` | glottis view   ```     Camera position: (122.84, -6.51, 185.88)     Camera target: (-3.00, -1.00, -72.00)     Camera orientation: 7.4, 1.6, 287.0 ``` |
| EL50N Yaw: 0.0° Roll: 30.0°   ``` Scenario: #2 (resolved) Subject: #2 EL50N Yaw: 0.0° Roll: 30.0° Tube lift: -5.802 mm Jaw contact X: 10.879 mm Jaw contact Y: 99.08 mm Tube position: (31.66, 100.44, -52.05) Tube rotation: (42.93, 0.00, 30.00) Upper incisor: (0.00, 102.00, -100.00) Lower incisor: (0.00, 100.00, -82.00) Glottic contact: (2.24, -5.80, -0.12) Preglottic contact: (1.23, -3.48, -10.00) Prepreglottic contact: (-0.50, 1.74, -20.00) Maxillary contact: (10.88, 99.08, -100.02) Blade contact: (-1.27, 49.61, -66.25) Vallecula: (--, 25.00, -33.00) Jaw width: 65.4mm Resolve speed: 0.01 Cycles: 102 Cycles at speed: 3 Auto: On Ghosts: Off Cache hit: No Results: 34 Camera position: (122.84, -6.51, 185.88) Camera target: (-3.00, -1.00, -72.00) Camera orientation: 7.4, 1.6, 287.0 ``` | intubation view   ```     Camera position: (20.99, 345.12, -201.12)     Camera target: (0.00, 0.00, 0.00)     Camera orientation: -7.8, 0.5, 400.0 ``` | side view   ```     Camera position: (305.62, 61.30, -59.83)     Camera target: (8.00, 94.00, -41.00)     Camera orientation: 6.2, 1.7, 300.0 ``` | top view   ```     Camera position: (11.26, 6.17, -377.84)     Camera target: (0.00, 110.00, 30.00)     Camera orientation: 4.7, 1.8, 421.0 ``` | above view   ```     Camera position: (-3.01, 315.98, -75.17)     Camera target: (-3.00, -1.00, -72.00)     Camera orientation: 4.7, 0.0, 317.0 ``` | glottis view   ```     Camera position: (122.84, -6.51, 185.88)     Camera target: (-3.00, -1.00, -72.00)     Camera orientation: 7.4, 1.6, 287.0 ``` |
| EL50N Yaw: 0.0° Roll: 45.0°   ``` Scenario: #3 (resolved) Subject: #2 EL50N Yaw: 0.0° Roll: 45.0° Tube lift: -9.945 mm Jaw contact X: 16.88 mm Jaw contact Y: 95.557 mm Tube position: (45.76, 89.63, -53.39) Tube rotation: (43.90, 0.00, 45.00) Upper incisor: (0.00, 102.00, -100.00) Lower incisor: (0.00, 100.00, -82.00) Glottic contact: (4.40, -9.94, -0.12) Preglottic contact: (2.92, -7.16, -10.00) Prepreglottic contact: (0.37, -1.05, -20.00) Maxillary contact: (16.88, 95.56, -100.45) Blade contact: (-0.12, 49.53, -66.19) Vallecula: (--, 25.00, -33.00) Jaw width: 65.4mm Resolve speed: 0.01 Cycles: 103 Cycles at speed: 3 Auto: On Ghosts: Off Cache hit: No Results: 35 Camera position: (122.84, -6.51, 185.88) Camera target: (-3.00, -1.00, -72.00) Camera orientation: 7.4, 1.6, 287.0 ``` | intubation view   ```     Camera position: (20.99, 345.12, -201.12)     Camera target: (0.00, 0.00, 0.00)     Camera orientation: -7.8, 0.5, 400.0 ``` | side view   ```     Camera position: (305.62, 61.30, -59.83)     Camera target: (8.00, 94.00, -41.00)     Camera orientation: 6.2, 1.7, 300.0 ``` | top view   ```     Camera position: (11.26, 6.17, -377.84)     Camera target: (0.00, 110.00, 30.00)     Camera orientation: 4.7, 1.8, 421.0 ``` | above view   ```     Camera position: (-3.01, 315.98, -75.17)     Camera target: (-3.00, -1.00, -72.00)     Camera orientation: 4.7, 0.0, 317.0 ``` | glottis view   ```     Camera position: (122.84, -6.51, 185.88)     Camera target: (-3.00, -1.00, -72.00)     Camera orientation: 7.4, 1.6, 287.0 ``` |
| EL50N Yaw: 5.0° Roll: 0.0°   ``` Scenario: #4 (resolved) Subject: #2 EL50N Yaw: 5.0° Roll: 0.0° Tube lift: -1.052 mm Jaw contact X: 8.366 mm Jaw contact Y: 99.939 mm Tube position: (5.42, 110.34, -51.70) Tube rotation: (41.54, 5.00, 0.00) Upper incisor: (0.00, 102.00, -100.00) Lower incisor: (0.00, 100.00, -82.00) Glottic contact: (-3.00, -1.05, -0.12) Preglottic contact: (-2.56, 0.84, -10.00) Prepreglottic contact: (-1.65, 5.22, -20.00) Maxillary contact: (8.37, 99.94, -99.92) Blade contact: (4.00, 49.15, -65.87) Vallecula: (--, 25.00, -33.00) Jaw width: 65.4mm Resolve speed: 0.01 Cycles: 104 Cycles at speed: 3 Auto: On Ghosts: Off Cache hit: No Results: 36 Camera position: (122.84, -6.51, 185.88) Camera target: (-3.00, -1.00, -72.00) Camera orientation: 7.4, 1.6, 287.0 ``` | intubation view   ```     Camera position: (20.99, 345.12, -201.12)     Camera target: (0.00, 0.00, 0.00)     Camera orientation: -7.8, 0.5, 400.0 ``` | side view   ```     Camera position: (305.62, 61.30, -59.83)     Camera target: (8.00, 94.00, -41.00)     Camera orientation: 6.2, 1.7, 300.0 ``` | top view   ```     Camera position: (11.26, 6.17, -377.84)     Camera target: (0.00, 110.00, 30.00)     Camera orientation: 4.7, 1.8, 421.0 ``` | above view   ```     Camera position: (-3.01, 315.98, -75.17)     Camera target: (-3.00, -1.00, -72.00)     Camera orientation: 4.7, 0.0, 317.0 ``` | glottis view   ```     Camera position: (122.84, -6.51, 185.88)     Camera target: (-3.00, -1.00, -72.00)     Camera orientation: 7.4, 1.6, 287.0 ``` |
| EL50N Yaw: 5.0° Roll: 15.0°   ``` Scenario: #5 (resolved) Subject: #2 EL50N Yaw: 5.0° Roll: 15.0° Tube lift: -1.045 mm Jaw contact X: 12.318 mm Jaw contact Y: 98.417 mm Tube position: (22.06, 107.74, -52.00) Tube rotation: (41.37, 5.00, 15.00) Upper incisor: (0.00, 102.00, -100.00) Lower incisor: (0.00, 100.00, -82.00) Glottic contact: (-2.93, -1.05, -0.12) Preglottic contact: (-2.99, 0.91, -10.00) Prepreglottic contact: (-2.95, 5.40, -20.00) Maxillary contact: (12.32, 98.42, -100.10) Blade contact: (2.00, 48.84, -65.62) Vallecula: (--, 25.00, -33.00) Jaw width: 65.4mm Resolve speed: 0.01 Cycles: 91 Cycles at speed: 3 Auto: On Ghosts: Off Cache hit: No Results: 37 Camera position: (122.84, -6.51, 185.88) Camera target: (-3.00, -1.00, -72.00) Camera orientation: 7.4, 1.6, 287.0 ``` | intubation view   ```     Camera position: (20.99, 345.12, -201.12)     Camera target: (0.00, 0.00, 0.00)     Camera orientation: -7.8, 0.5, 400.0 ``` | side view   ```     Camera position: (305.62, 61.30, -59.83)     Camera target: (8.00, 94.00, -41.00)     Camera orientation: 6.2, 1.7, 300.0 ``` | top view   ```     Camera position: (11.26, 6.17, -377.84)     Camera target: (0.00, 110.00, 30.00)     Camera orientation: 4.7, 1.8, 421.0 ``` | above view   ```     Camera position: (-3.01, 315.98, -75.17)     Camera target: (-3.00, -1.00, -72.00)     Camera orientation: 4.7, 0.0, 317.0 ``` | glottis view   ```     Camera position: (122.84, -6.51, 185.88)     Camera target: (-3.00, -1.00, -72.00)     Camera orientation: 7.4, 1.6, 287.0 ``` |
| EL50N Yaw: 5.0° Roll: 30.0°   ``` Scenario: #6 (resolved) Subject: #2 EL50N Yaw: 5.0° Roll: 30.0° Tube lift: -2.478 mm Jaw contact X: 17.113 mm Jaw contact Y: 95.335 mm Tube position: (38.16, 100.82, -52.86) Tube rotation: (41.48, 5.00, 30.00) Upper incisor: (0.00, 102.00, -100.00) Lower incisor: (0.00, 100.00, -82.00) Glottic contact: (-1.36, -2.48, -0.12) Preglottic contact: (-1.90, -0.29, -10.00) Prepreglottic contact: (-2.67, 4.62, -20.00) Maxillary contact: (17.11, 95.33, -100.48) Blade contact: (1.73, 48.39, -65.26) Vallecula: (--, 25.00, -33.00) Jaw width: 65.4mm Resolve speed: 0.01 Cycles: 82 Cycles at speed: 1 Auto: On Ghosts: Off Cache hit: No Results: 38 Camera position: (122.84, -6.51, 185.88) Camera target: (-3.00, -1.00, -72.00) Camera orientation: 7.4, 1.6, 287.0 ``` | intubation view   ```     Camera position: (20.99, 345.12, -201.12)     Camera target: (0.00, 0.00, 0.00)     Camera orientation: -7.8, 0.5, 400.0 ``` | side view   ```     Camera position: (305.62, 61.30, -59.83)     Camera target: (8.00, 94.00, -41.00)     Camera orientation: 6.2, 1.7, 300.0 ``` | top view   ```     Camera position: (11.26, 6.17, -377.84)     Camera target: (0.00, 110.00, 30.00)     Camera orientation: 4.7, 1.8, 421.0 ``` | above view   ```     Camera position: (-3.01, 315.98, -75.17)     Camera target: (-3.00, -1.00, -72.00)     Camera orientation: 4.7, 0.0, 317.0 ``` | glottis view   ```     Camera position: (122.84, -6.51, 185.88)     Camera target: (-3.00, -1.00, -72.00)     Camera orientation: 7.4, 1.6, 287.0 ``` |
| EL50N Yaw: 5.0° Roll: 45.0°   ``` Scenario: #7 (resolved) Subject: #2 EL50N Yaw: 5.0° Roll: 45.0° Tube lift: -5.025 mm Jaw contact X: 21.414 mm Jaw contact Y: 90.774 mm Tube position: (52.05, 90.23, -54.58) Tube rotation: (41.80, 5.00, 45.00) Upper incisor: (0.00, 102.00, -100.00) Lower incisor: (0.00, 100.00, -82.00) Glottic contact: (0.49, -5.03, -0.12) Preglottic contact: (-0.48, -2.45, -10.00) Prepreglottic contact: (-1.98, 3.17, -20.00) Maxillary contact: (21.41, 90.77, -101.03) Blade contact: (2.17, 47.88, -64.85) Vallecula: (--, 25.00, -33.00) Jaw width: 65.4mm Resolve speed: 0.01 Cycles: 96 Cycles at speed: 3 Auto: On Ghosts: Off Cache hit: No Results: 39 Camera position: (122.84, -6.51, 185.88) Camera target: (-3.00, -1.00, -72.00) Camera orientation: 7.4, 1.6, 287.0 ``` | intubation view   ```     Camera position: (20.99, 345.12, -201.12)     Camera target: (0.00, 0.00, 0.00)     Camera orientation: -7.8, 0.5, 400.0 ``` | side view   ```     Camera position: (305.62, 61.30, -59.83)     Camera target: (8.00, 94.00, -41.00)     Camera orientation: 6.2, 1.7, 300.0 ``` | top view   ```     Camera position: (11.26, 6.17, -377.84)     Camera target: (0.00, 110.00, 30.00)     Camera orientation: 4.7, 1.8, 421.0 ``` | above view   ```     Camera position: (-3.01, 315.98, -75.17)     Camera target: (-3.00, -1.00, -72.00)     Camera orientation: 4.7, 0.0, 317.0 ``` | glottis view   ```     Camera position: (122.84, -6.51, 185.88)     Camera target: (-3.00, -1.00, -72.00)     Camera orientation: 7.4, 1.6, 287.0 ``` |
| EL50N Yaw: 10.0° Roll: 0.0°   ``` Scenario: #8 (resolved) Subject: #2 EL50N Yaw: 10.0° Roll: 0.0° Tube lift: 3.318 mm Jaw contact X: 18.251 mm Jaw contact Y: 94.338 mm Tube position: (13.77, 112.79, -53.43) Tube rotation: (39.43, 10.00, 0.00) Upper incisor: (0.00, 102.00, -100.00) Lower incisor: (0.00, 100.00, -82.00) Glottic contact: (-3.00, 3.32, -0.12) Preglottic contact: (-2.13, 4.98, -10.00) Prepreglottic contact: (-0.34, 8.89, -20.00) Maxillary contact: (18.25, 94.34, -100.59) Blade contact: (8.32, 46.23, -63.42) Vallecula: (--, 25.00, -33.00) Jaw width: 65.4mm Resolve speed: 0.01 Cycles: 97 Cycles at speed: 3 Auto: On Ghosts: Off Cache hit: No Results: 40 Camera position: (122.84, -6.51, 185.88) Camera target: (-3.00, -1.00, -72.00) Camera orientation: 7.4, 1.6, 287.0 ``` | intubation view   ```     Camera position: (20.99, 345.12, -201.12)     Camera target: (0.00, 0.00, 0.00)     Camera orientation: -7.8, 0.5, 400.0 ``` | side view   ```     Camera position: (305.62, 61.30, -59.83)     Camera target: (8.00, 94.00, -41.00)     Camera orientation: 6.2, 1.7, 300.0 ``` | top view   ```     Camera position: (11.26, 6.17, -377.84)     Camera target: (0.00, 110.00, 30.00)     Camera orientation: 4.7, 1.8, 421.0 ``` | above view   ```     Camera position: (-3.01, 315.98, -75.17)     Camera target: (-3.00, -1.00, -72.00)     Camera orientation: 4.7, 0.0, 317.0 ``` | glottis view   ```     Camera position: (122.84, -6.51, 185.88)     Camera target: (-3.00, -1.00, -72.00)     Camera orientation: 7.4, 1.6, 287.0 ``` |
| EL50N Yaw: 10.0° Roll: 15.0°   ``` Scenario: #9 (resolved) Subject: #2 EL50N Yaw: 10.0° Roll: 15.0° Tube lift: 4.163 mm Jaw contact X: 20.8 mm Jaw contact Y: 91.556 mm Tube position: (30.19, 109.81, -53.99) Tube rotation: (38.69, 10.00, 15.00) Upper incisor: (0.00, 102.00, -100.00) Lower incisor: (0.00, 100.00, -82.00) Glottic contact: (-3.00, 4.16, -0.12) Preglottic contact: (-2.63, 5.85, -10.00) Prepreglottic contact: (-1.69, 9.76, -20.00) Maxillary contact: (20.80, 91.56, -100.93) Blade contact: (7.06, 46.61, -63.61) Vallecula: (--, 25.00, -33.00) Jaw width: 65.4mm Resolve speed: 0.01 Cycles: 98 Cycles at speed: 2 Auto: On Ghosts: Off Cache hit: No Results: 41 Camera position: (122.84, -6.51, 185.88) Camera target: (-3.00, -1.00, -72.00) Camera orientation: 7.4, 1.6, 287.0 ``` | intubation view   ```     Camera position: (20.99, 345.12, -201.12)     Camera target: (0.00, 0.00, 0.00)     Camera orientation: -7.8, 0.5, 400.0 ``` | side view   ```     Camera position: (305.62, 61.30, -59.83)     Camera target: (8.00, 94.00, -41.00)     Camera orientation: 6.2, 1.7, 300.0 ``` | top view   ```     Camera position: (11.26, 6.17, -377.84)     Camera target: (0.00, 110.00, 30.00)     Camera orientation: 4.7, 1.8, 421.0 ``` | above view   ```     Camera position: (-3.01, 315.98, -75.17)     Camera target: (-3.00, -1.00, -72.00)     Camera orientation: 4.7, 0.0, 317.0 ``` | glottis view   ```     Camera position: (122.84, -6.51, 185.88)     Camera target: (-3.00, -1.00, -72.00)     Camera orientation: 7.4, 1.6, 287.0 ``` |
| EL50N Yaw: 10.0° Roll: 30.0°   ``` Scenario: #10 (resolved) Subject: #2 EL50N Yaw: 10.0° Roll: 30.0° Tube lift: 3.494 mm Jaw contact X: 23.07 mm Jaw contact Y: 88.38 mm Tube position: (45.37, 102.45, -54.93) Tube rotation: (38.40, 10.00, 30.00) Upper incisor: (0.00, 102.00, -100.00) Lower incisor: (0.00, 100.00, -82.00) Glottic contact: (-2.91, 3.49, -0.12) Preglottic contact: (-3.00, 5.37, -10.00) Prepreglottic contact: (-2.84, 9.61, -20.00) Maxillary contact: (23.07, 88.38, -101.31) Blade contact: (5.38, 45.95, -62.98) Vallecula: (--, 25.00, -33.00) Jaw width: 65.4mm Resolve speed: 0.01 Cycles: 90 Cycles at speed: 3 Auto: On Ghosts: Off Cache hit: No Results: 42 Camera position: (122.84, -6.51, 185.88) Camera target: (-3.00, -1.00, -72.00) Camera orientation: 7.4, 1.6, 287.0 ``` | intubation view   ```     Camera position: (20.99, 345.12, -201.12)     Camera target: (0.00, 0.00, 0.00)     Camera orientation: -7.8, 0.5, 400.0 ``` | side view   ```     Camera position: (305.62, 61.30, -59.83)     Camera target: (8.00, 94.00, -41.00)     Camera orientation: 6.2, 1.7, 300.0 ``` | top view   ```     Camera position: (11.26, 6.17, -377.84)     Camera target: (0.00, 110.00, 30.00)     Camera orientation: 4.7, 1.8, 421.0 ``` | above view   ```     Camera position: (-3.01, 315.98, -75.17)     Camera target: (-3.00, -1.00, -72.00)     Camera orientation: 4.7, 0.0, 317.0 ``` | glottis view   ```     Camera position: (122.84, -6.51, 185.88)     Camera target: (-3.00, -1.00, -72.00)     Camera orientation: 7.4, 1.6, 287.0 ``` |
| EL50N Yaw: 10.0° Roll: 45.0°   ``` Scenario: #11 (resolved) Subject: #2 EL50N Yaw: 10.0° Roll: 45.0° Tube lift: 1.882 mm Jaw contact X: 25.491 mm Jaw contact Y: 84.004 mm Tube position: (58.67, 91.44, -56.78) Tube rotation: (38.21, 10.00, 45.00) Upper incisor: (0.00, 102.00, -100.00) Lower incisor: (0.00, 100.00, -82.00) Glottic contact: (-1.95, 1.88, -0.12) Preglottic contact: (-2.44, 4.08, -10.00) Prepreglottic contact: (-2.97, 8.90, -20.00) Maxillary contact: (25.49, 84.00, -101.85) Blade contact: (4.33, 44.94, -61.93) Vallecula: (--, 25.00, -33.00) Jaw width: 65.4mm Resolve speed: 0.01 Cycles: 89 Cycles at speed: 3 Auto: On Ghosts: Off Cache hit: No Results: 43 Camera position: (122.84, -6.51, 185.88) Camera target: (-3.00, -1.00, -72.00) Camera orientation: 7.4, 1.6, 287.0 ``` | intubation view   ```     Camera position: (20.99, 345.12, -201.12)     Camera target: (0.00, 0.00, 0.00)     Camera orientation: -7.8, 0.5, 400.0 ``` | side view   ```     Camera position: (305.62, 61.30, -59.83)     Camera target: (8.00, 94.00, -41.00)     Camera orientation: 6.2, 1.7, 300.0 ``` | top view   ```     Camera position: (11.26, 6.17, -377.84)     Camera target: (0.00, 110.00, 30.00)     Camera orientation: 4.7, 1.8, 421.0 ``` | above view   ```     Camera position: (-3.01, 315.98, -75.17)     Camera target: (-3.00, -1.00, -72.00)     Camera orientation: 4.7, 0.0, 317.0 ``` | glottis view   ```     Camera position: (122.84, -6.51, 185.88)     Camera target: (-3.00, -1.00, -72.00)     Camera orientation: 7.4, 1.6, 287.0 ``` |
| EL50N Yaw: 15.0° Roll: 0.0°   ``` Scenario: #12 (resolved) Subject: #2 EL50N Yaw: 15.0° Roll: 0.0° Tube lift: 10.506 mm Jaw contact X: 25.876 mm Jaw contact Y: 83.23 mm Tube position: (21.99, 115.50, -57.98) Tube rotation: (34.38, 15.00, 0.00) Upper incisor: (0.00, 102.00, -100.00) Lower incisor: (0.00, 100.00, -82.00) Glottic contact: (-3.00, 10.51, -0.12) Preglottic contact: (-1.72, 11.64, -10.00) Prepreglottic contact: (0.92, 14.49, -20.00) Maxillary contact: (25.88, 83.23, -101.93) Blade contact: (5.19, 24.96, -33.17) Vallecula: (--, 25.00, -33.00) Jaw width: 65.4mm Resolve speed: 0.01 Cycles: 98 Cycles at speed: 3 Auto: On Ghosts: Off Cache hit: No Results: 44 Camera position: (122.84, -6.51, 185.88) Camera target: (-3.00, -1.00, -72.00) Camera orientation: 7.4, 1.6, 287.0 ``` | intubation view   ```     Camera position: (20.99, 345.12, -201.12)     Camera target: (0.00, 0.00, 0.00)     Camera orientation: -7.8, 0.5, 400.0 ``` | side view   ```     Camera position: (305.62, 61.30, -59.83)     Camera target: (8.00, 94.00, -41.00)     Camera orientation: 6.2, 1.7, 300.0 ``` | top view   ```     Camera position: (11.26, 6.17, -377.84)     Camera target: (0.00, 110.00, 30.00)     Camera orientation: 4.7, 1.8, 421.0 ``` | above view   ```     Camera position: (-3.01, 315.98, -75.17)     Camera target: (-3.00, -1.00, -72.00)     Camera orientation: 4.7, 0.0, 317.0 ``` | glottis view   ```     Camera position: (122.84, -6.51, 185.88)     Camera target: (-3.00, -1.00, -72.00)     Camera orientation: 7.4, 1.6, 287.0 ``` |
| EL50N Yaw: 15.0° Roll: 15.0°   ``` Scenario: #13 (resolved) Subject: #2 EL50N Yaw: 15.0° Roll: 15.0° Tube lift: 10.74 mm Jaw contact X: 27.342 mm Jaw contact Y: 79.819 mm Tube position: (38.10, 110.90, -58.32) Tube rotation: (33.28, 15.00, 15.00) Upper incisor: (0.00, 102.00, -100.00) Lower incisor: (0.00, 100.00, -82.00) Glottic contact: (-3.00, 10.74, -0.12) Preglottic contact: (-2.21, 11.88, -10.00) Prepreglottic contact: (-0.43, 14.68, -20.00) Maxillary contact: (27.34, 79.82, -102.35) Blade contact: (2.86, 24.96, -33.16) Vallecula: (--, 25.00, -33.00) Jaw width: 65.4mm Resolve speed: 0.01 Cycles: 90 Cycles at speed: 1 Auto: On Ghosts: Off Cache hit: No Results: 45 Camera position: (122.84, -6.51, 185.88) Camera target: (-3.00, -1.00, -72.00) Camera orientation: 7.4, 1.6, 287.0 ``` | intubation view   ```     Camera position: (20.99, 345.12, -201.12)     Camera target: (0.00, 0.00, 0.00)     Camera orientation: -7.8, 0.5, 400.0 ``` | side view   ```     Camera position: (305.62, 61.30, -59.83)     Camera target: (8.00, 94.00, -41.00)     Camera orientation: 6.2, 1.7, 300.0 ``` | top view   ```     Camera position: (11.26, 6.17, -377.84)     Camera target: (0.00, 110.00, 30.00)     Camera orientation: 4.7, 1.8, 421.0 ``` | above view   ```     Camera position: (-3.01, 315.98, -75.17)     Camera target: (-3.00, -1.00, -72.00)     Camera orientation: 4.7, 0.0, 317.0 ``` | glottis view   ```     Camera position: (122.84, -6.51, 185.88)     Camera target: (-3.00, -1.00, -72.00)     Camera orientation: 7.4, 1.6, 287.0 ``` |
| EL50N Yaw: 15.0° Roll: 30.0°   ``` Scenario: #14 (resolved) Subject: #2 EL50N Yaw: 15.0° Roll: 30.0° Tube lift: 10.124 mm Jaw contact X: 28.531 mm Jaw contact Y: 76.512 mm Tube position: (52.95, 102.47, -59.10) Tube rotation: (32.60, 15.00, 30.00) Upper incisor: (0.00, 102.00, -100.00) Lower incisor: (0.00, 100.00, -82.00) Glottic contact: (-3.00, 10.12, -0.12) Preglottic contact: (-2.65, 11.42, -10.00) Prepreglottic contact: (-1.64, 14.48, -20.00) Maxillary contact: (28.53, 76.51, -102.76) Blade contact: (0.81, 24.97, -33.16) Vallecula: (--, 25.00, -33.00) Jaw width: 65.4mm Resolve speed: 0.01 Cycles: 75 Cycles at speed: 1 Auto: On Ghosts: Off Cache hit: No Results: 46 Camera position: (122.84, -6.51, 185.88) Camera target: (-3.00, -1.00, -72.00) Camera orientation: 7.4, 1.6, 287.0 ``` | intubation view   ```     Camera position: (20.99, 345.12, -201.12)     Camera target: (0.00, 0.00, 0.00)     Camera orientation: -7.8, 0.5, 400.0 ``` | side view   ```     Camera position: (305.62, 61.30, -59.83)     Camera target: (8.00, 94.00, -41.00)     Camera orientation: 6.2, 1.7, 300.0 ``` | top view   ```     Camera position: (11.26, 6.17, -377.84)     Camera target: (0.00, 110.00, 30.00)     Camera orientation: 4.7, 1.8, 421.0 ``` | above view   ```     Camera position: (-3.01, 315.98, -75.17)     Camera target: (-3.00, -1.00, -72.00)     Camera orientation: 4.7, 0.0, 317.0 ``` | glottis view   ```     Camera position: (122.84, -6.51, 185.88)     Camera target: (-3.00, -1.00, -72.00)     Camera orientation: 7.4, 1.6, 287.0 ``` |
| EL50N Yaw: 15.0° Roll: 45.0°   ``` Scenario: #15 (resolved) Subject: #2 EL50N Yaw: 15.0° Roll: 45.0° Tube lift: 8.768 mm Jaw contact X: 29.493 mm Jaw contact Y: 73.215 mm Tube position: (65.55, 90.58, -60.48) Tube rotation: (32.24, 15.00, 45.00) Upper incisor: (0.00, 102.00, -100.00) Lower incisor: (0.00, 100.00, -82.00) Glottic contact: (-2.97, 8.77, -0.12) Preglottic contact: (-3.00, 10.36, -10.00) Prepreglottic contact: (-2.62, 13.93, -20.00) Maxillary contact: (29.49, 73.22, -103.17) Blade contact: (-0.83, 24.98, -33.17) Vallecula: (--, 25.00, -33.00) Jaw width: 65.4mm Resolve speed: 0.01 Cycles: 84 Cycles at speed: 1 Auto: On Ghosts: Off Cache hit: No Results: 47 Camera position: (122.84, -6.51, 185.88) Camera target: (-3.00, -1.00, -72.00) Camera orientation: 7.4, 1.6, 287.0 ``` | intubation view   ```     Camera position: (20.99, 345.12, -201.12)     Camera target: (0.00, 0.00, 0.00)     Camera orientation: -7.8, 0.5, 400.0 ``` | side view   ```     Camera position: (305.62, 61.30, -59.83)     Camera target: (8.00, 94.00, -41.00)     Camera orientation: 6.2, 1.7, 300.0 ``` | top view   ```     Camera position: (11.26, 6.17, -377.84)     Camera target: (0.00, 110.00, 30.00)     Camera orientation: 4.7, 1.8, 421.0 ``` | above view   ```     Camera position: (-3.01, 315.98, -75.17)     Camera target: (-3.00, -1.00, -72.00)     Camera orientation: 4.7, 0.0, 317.0 ``` | glottis view   ```     Camera position: (122.84, -6.51, 185.88)     Camera target: (-3.00, -1.00, -72.00)     Camera orientation: 7.4, 1.6, 287.0 ``` |
| DL50N Yaw: 0.0° Roll: 0.0°   ``` Scenario: #0 (resolved) Subject: #3 DL50N Yaw: 0.0° Roll: 0.0° Tube lift: -10.026 mm Jaw contact X: -2.886 mm Jaw contact Y: 106.199 mm Tube position: (-3.00, 105.24, -41.08) Tube rotation: (51.28, 0.00, 0.00) Upper incisor: (0.00, 108.00, -82.00) Lower incisor: (0.00, 97.00, -66.00) Glottic contact: (-3.00, -10.03, -0.12) Preglottic contact: (-3.00, -7.04, -10.00) Prepreglottic contact: (-3.00, -0.33, -20.00) Maxillary contact: (-2.89, 106.20, -81.79) Blade contact: (-3.00, 56.27, -59.80) Vallecula: (--, 25.00, -33.00) Jaw width: 65.4mm Resolve speed: 0.01 Cycles: 104 Cycles at speed: 2 Auto: On Ghosts: Off Cache hit: No Results: 48 Camera position: (122.84, -6.51, 185.88) Camera target: (-3.00, -1.00, -72.00) Camera orientation: 7.4, 1.6, 287.0 ``` | intubation view   ```     Camera position: (20.99, 345.12, -201.12)     Camera target: (0.00, 0.00, 0.00)     Camera orientation: -7.8, 0.5, 400.0 ``` | side view   ```     Camera position: (305.62, 61.30, -59.83)     Camera target: (8.00, 94.00, -41.00)     Camera orientation: 6.2, 1.7, 300.0 ``` | top view   ```     Camera position: (11.26, 6.17, -377.84)     Camera target: (0.00, 110.00, 30.00)     Camera orientation: 4.7, 1.8, 421.0 ``` | above view   ```     Camera position: (-3.01, 315.98, -75.17)     Camera target: (-3.00, -1.00, -72.00)     Camera orientation: 4.7, 0.0, 317.0 ``` | glottis view   ```     Camera position: (122.84, -6.51, 185.88)     Camera target: (-3.00, -1.00, -72.00)     Camera orientation: 7.4, 1.6, 287.0 ``` |
| DL50N Yaw: 0.0° Roll: 15.0°   ``` Scenario: #1 (resolved) Subject: #3 DL50N Yaw: 0.0° Roll: 15.0° Tube lift: -13.117 mm Jaw contact X: 4.54 mm Jaw contact Y: 105.979 mm Tube position: (15.03, 100.88, -41.07) Tube rotation: (51.55, 0.00, 15.00) Upper incisor: (0.00, 108.00, -82.00) Lower incisor: (0.00, 97.00, -66.00) Glottic contact: (-0.28, -13.12, -0.12) Preglottic contact: (-0.84, -10.01, -10.00) Prepreglottic contact: (-1.78, -3.08, -20.00) Maxillary contact: (4.54, 105.98, -81.80) Blade contact: (-1.69, 56.11, -60.46) Vallecula: (--, 25.00, -33.00) Jaw width: 65.4mm Resolve speed: 0.01 Cycles: 99 Cycles at speed: 3 Auto: On Ghosts: Off Cache hit: No Results: 49 Camera position: (122.84, -6.51, 185.88) Camera target: (-3.00, -1.00, -72.00) Camera orientation: 7.4, 1.6, 287.0 ``` | intubation view   ```     Camera position: (20.99, 345.12, -201.12)     Camera target: (0.00, 0.00, 0.00)     Camera orientation: -7.8, 0.5, 400.0 ``` | side view   ```     Camera position: (305.62, 61.30, -59.83)     Camera target: (8.00, 94.00, -41.00)     Camera orientation: 6.2, 1.7, 300.0 ``` | top view   ```     Camera position: (11.26, 6.17, -377.84)     Camera target: (0.00, 110.00, 30.00)     Camera orientation: 4.7, 1.8, 421.0 ``` | above view   ```     Camera position: (-3.01, 315.98, -75.17)     Camera target: (-3.00, -1.00, -72.00)     Camera orientation: 4.7, 0.0, 317.0 ``` | glottis view   ```     Camera position: (122.84, -6.51, 185.88)     Camera target: (-3.00, -1.00, -72.00)     Camera orientation: 7.4, 1.6, 287.0 ``` |
| DL50N Yaw: 0.0° Roll: 30.0°   ``` Scenario: #2 (resolved) Subject: #3 DL50N Yaw: 0.0° Roll: 30.0° Tube lift: -16.632 mm Jaw contact X: 11.96 mm Jaw contact Y: 103.972 mm Tube position: (31.66, 93.62, -41.31) Tube rotation: (52.16, 0.00, 30.00) Upper incisor: (0.00, 108.00, -82.00) Lower incisor: (0.00, 97.00, -66.00) Glottic contact: (2.23, -16.63, -0.12) Preglottic contact: (1.14, -13.22, -10.00) Prepreglottic contact: (-0.73, -5.62, -20.00) Maxillary contact: (11.96, 103.97, -82.08) Blade contact: (-0.08, 56.22, -60.51) Vallecula: (--, 25.00, -33.00) Jaw width: 65.4mm Resolve speed: 0.01 Cycles: 101 Cycles at speed: 3 Auto: On Ghosts: Off Cache hit: No Results: 50 Camera position: (122.84, -6.51, 185.88) Camera target: (-3.00, -1.00, -72.00) Camera orientation: 7.4, 1.6, 287.0 ``` | intubation view   ```     Camera position: (20.99, 345.12, -201.12)     Camera target: (0.00, 0.00, 0.00)     Camera orientation: -7.8, 0.5, 400.0 ``` | side view   ```     Camera position: (305.62, 61.30, -59.83)     Camera target: (8.00, 94.00, -41.00)     Camera orientation: 6.2, 1.7, 300.0 ``` | top view   ```     Camera position: (11.26, 6.17, -377.84)     Camera target: (0.00, 110.00, 30.00)     Camera orientation: 4.7, 1.8, 421.0 ``` | above view   ```     Camera position: (-3.01, 315.98, -75.17)     Camera target: (-3.00, -1.00, -72.00)     Camera orientation: 4.7, 0.0, 317.0 ``` | glottis view   ```     Camera position: (122.84, -6.51, 185.88)     Camera target: (-3.00, -1.00, -72.00)     Camera orientation: 7.4, 1.6, 287.0 ``` |
| DL50N Yaw: 0.0° Roll: 45.0°   ``` Scenario: #3 (resolved) Subject: #3 DL50N Yaw: 0.0° Roll: 45.0° Tube lift: -22.186 mm Jaw contact X: 18.585 mm Jaw contact Y: 99.72 mm Tube position: (45.76, 82.29, -41.99) Tube rotation: (53.15, 0.00, 45.00) Upper incisor: (0.00, 108.00, -82.00) Lower incisor: (0.00, 97.00, -66.00) Glottic contact: (4.40, -22.19, -0.12) Preglottic contact: (2.76, -18.20, -10.00) Prepreglottic contact: (-0.02, -9.45, -20.00) Maxillary contact: (18.58, 99.72, -82.64) Blade contact: (2.07, 56.11, -60.47) Vallecula: (--, 25.00, -33.00) Jaw width: 65.4mm Resolve speed: 0.01 Cycles: 85 Cycles at speed: 1 Auto: On Ghosts: Off Cache hit: No Results: 51 Camera position: (122.84, -6.51, 185.88) Camera target: (-3.00, -1.00, -72.00) Camera orientation: 7.4, 1.6, 287.0 ``` | intubation view   ```     Camera position: (20.99, 345.12, -201.12)     Camera target: (0.00, 0.00, 0.00)     Camera orientation: -7.8, 0.5, 400.0 ``` | side view   ```     Camera position: (305.62, 61.30, -59.83)     Camera target: (8.00, 94.00, -41.00)     Camera orientation: 6.2, 1.7, 300.0 ``` | top view   ```     Camera position: (11.26, 6.17, -377.84)     Camera target: (0.00, 110.00, 30.00)     Camera orientation: 4.7, 1.8, 421.0 ``` | above view   ```     Camera position: (-3.01, 315.98, -75.17)     Camera target: (-3.00, -1.00, -72.00)     Camera orientation: 4.7, 0.0, 317.0 ``` | glottis view   ```     Camera position: (122.84, -6.51, 185.88)     Camera target: (-3.00, -1.00, -72.00)     Camera orientation: 7.4, 1.6, 287.0 ``` |
| DL50N Yaw: 5.0° Roll: 0.0°   ``` Scenario: #4 (resolved) Subject: #3 DL50N Yaw: 5.0° Roll: 0.0° Tube lift: -11.043 mm Jaw contact X: 8.708 mm Jaw contact Y: 105.138 mm Tube position: (5.42, 103.89, -41.14) Tube rotation: (51.06, 5.00, 0.00) Upper incisor: (0.00, 108.00, -82.00) Lower incisor: (0.00, 97.00, -66.00) Glottic contact: (-3.00, -11.04, -0.12) Preglottic contact: (-2.53, -8.10, -10.00) Prepreglottic contact: (-1.53, -1.46, -20.00) Maxillary contact: (8.71, 105.14, -81.92) Blade contact: (4.75, 55.84, -60.33) Vallecula: (--, 25.00, -33.00) Jaw width: 65.4mm Resolve speed: 0.01 Cycles: 96 Cycles at speed: 1 Auto: On Ghosts: Off Cache hit: No Results: 52 Camera position: (122.84, -6.51, 185.88) Camera target: (-3.00, -1.00, -72.00) Camera orientation: 7.4, 1.6, 287.0 ``` | intubation view   ```     Camera position: (20.99, 345.12, -201.12)     Camera target: (0.00, 0.00, 0.00)     Camera orientation: -7.8, 0.5, 400.0 ``` | side view   ```     Camera position: (305.62, 61.30, -59.83)     Camera target: (8.00, 94.00, -41.00)     Camera orientation: 6.2, 1.7, 300.0 ``` | top view   ```     Camera position: (11.26, 6.17, -377.84)     Camera target: (0.00, 110.00, 30.00)     Camera orientation: 4.7, 1.8, 421.0 ``` | above view   ```     Camera position: (-3.01, 315.98, -75.17)     Camera target: (-3.00, -1.00, -72.00)     Camera orientation: 4.7, 0.0, 317.0 ``` | glottis view   ```     Camera position: (122.84, -6.51, 185.88)     Camera target: (-3.00, -1.00, -72.00)     Camera orientation: 7.4, 1.6, 287.0 ``` |
| DL50N Yaw: 5.0° Roll: 15.0°   ``` Scenario: #5 (resolved) Subject: #3 DL50N Yaw: 5.0° Roll: 15.0° Tube lift: -11.292 mm Jaw contact X: 12.97 mm Jaw contact Y: 103.462 mm Tube position: (22.06, 101.20, -41.32) Tube rotation: (51.02, 5.00, 15.00) Upper incisor: (0.00, 108.00, -82.00) Lower incisor: (0.00, 97.00, -66.00) Glottic contact: (-2.92, -11.29, -0.12) Preglottic contact: (-2.99, -8.24, -10.00) Prepreglottic contact: (-2.92, -1.44, -20.00) Maxillary contact: (12.97, 103.46, -82.13) Blade contact: (3.25, 55.62, -60.23) Vallecula: (--, 25.00, -33.00) Jaw width: 65.4mm Resolve speed: 0.01 Cycles: 94 Cycles at speed: 2 Auto: On Ghosts: Off Cache hit: No Results: 53 Camera position: (122.84, -6.51, 185.88) Camera target: (-3.00, -1.00, -72.00) Camera orientation: 7.4, 1.6, 287.0 ``` | intubation view   ```     Camera position: (20.99, 345.12, -201.12)     Camera target: (0.00, 0.00, 0.00)     Camera orientation: -7.8, 0.5, 400.0 ``` | side view   ```     Camera position: (305.62, 61.30, -59.83)     Camera target: (8.00, 94.00, -41.00)     Camera orientation: 6.2, 1.7, 300.0 ``` | top view   ```     Camera position: (11.26, 6.17, -377.84)     Camera target: (0.00, 110.00, 30.00)     Camera orientation: 4.7, 1.8, 421.0 ``` | above view   ```     Camera position: (-3.01, 315.98, -75.17)     Camera target: (-3.00, -1.00, -72.00)     Camera orientation: 4.7, 0.0, 317.0 ``` | glottis view   ```     Camera position: (122.84, -6.51, 185.88)     Camera target: (-3.00, -1.00, -72.00)     Camera orientation: 7.4, 1.6, 287.0 ``` |
| DL50N Yaw: 5.0° Roll: 30.0°   ``` Scenario: #6 (resolved) Subject: #3 DL50N Yaw: 5.0° Roll: 30.0° Tube lift: -13.514 mm Jaw contact X: 18.115 mm Jaw contact Y: 100.048 mm Tube position: (38.16, 94.05, -41.76) Tube rotation: (51.29, 5.00, 30.00) Upper incisor: (0.00, 108.00, -82.00) Lower incisor: (0.00, 97.00, -66.00) Glottic contact: (-1.37, -13.51, -0.12) Preglottic contact: (-1.95, -10.18, -10.00) Prepreglottic contact: (-2.75, -2.81, -20.00) Maxillary contact: (18.12, 100.05, -82.58) Blade contact: (3.67, 55.35, -60.09) Vallecula: (--, 25.00, -33.00) Jaw width: 65.4mm Resolve speed: 0.01 Cycles: 93 Cycles at speed: 2 Auto: On Ghosts: Off Cache hit: No Results: 54 Camera position: (122.84, -6.51, 185.88) Camera target: (-3.00, -1.00, -72.00) Camera orientation: 7.4, 1.6, 287.0 ``` | intubation view   ```     Camera position: (20.99, 345.12, -201.12)     Camera target: (0.00, 0.00, 0.00)     Camera orientation: -7.8, 0.5, 400.0 ``` | side view   ```     Camera position: (305.62, 61.30, -59.83)     Camera target: (8.00, 94.00, -41.00)     Camera orientation: 6.2, 1.7, 300.0 ``` | top view   ```     Camera position: (11.26, 6.17, -377.84)     Camera target: (0.00, 110.00, 30.00)     Camera orientation: 4.7, 1.8, 421.0 ``` | above view   ```     Camera position: (-3.01, 315.98, -75.17)     Camera target: (-3.00, -1.00, -72.00)     Camera orientation: 4.7, 0.0, 317.0 ``` | glottis view   ```     Camera position: (122.84, -6.51, 185.88)     Camera target: (-3.00, -1.00, -72.00)     Camera orientation: 7.4, 1.6, 287.0 ``` |
| DL50N Yaw: 5.0° Roll: 45.0°   ``` Scenario: #7 (resolved) Subject: #3 DL50N Yaw: 5.0° Roll: 45.0° Tube lift: -17.169 mm Jaw contact X: 22.611 mm Jaw contact Y: 94.855 mm Tube position: (52.05, 83.40, -42.78) Tube rotation: (51.82, 5.00, 45.00) Upper incisor: (0.00, 108.00, -82.00) Lower incisor: (0.00, 97.00, -66.00) Glottic contact: (0.49, -17.17, -0.12) Preglottic contact: (-0.58, -13.34, -10.00) Prepreglottic contact: (-2.19, -4.98, -20.00) Maxillary contact: (22.61, 94.85, -83.24) Blade contact: (5.00, 54.70, -59.78) Vallecula: (--, 25.00, -33.00) Jaw width: 65.4mm Resolve speed: 0.01 Cycles: 88 Cycles at speed: 2 Auto: On Ghosts: Off Cache hit: No Results: 55 Camera position: (122.84, -6.51, 185.88) Camera target: (-3.00, -1.00, -72.00) Camera orientation: 7.4, 1.6, 287.0 ``` | intubation view   ```     Camera position: (20.99, 345.12, -201.12)     Camera target: (0.00, 0.00, 0.00)     Camera orientation: -7.8, 0.5, 400.0 ``` | side view   ```     Camera position: (305.62, 61.30, -59.83)     Camera target: (8.00, 94.00, -41.00)     Camera orientation: 6.2, 1.7, 300.0 ``` | top view   ```     Camera position: (11.26, 6.17, -377.84)     Camera target: (0.00, 110.00, 30.00)     Camera orientation: 4.7, 1.8, 421.0 ``` | above view   ```     Camera position: (-3.01, 315.98, -75.17)     Camera target: (-3.00, -1.00, -72.00)     Camera orientation: 4.7, 0.0, 317.0 ``` | glottis view   ```     Camera position: (122.84, -6.51, 185.88)     Camera target: (-3.00, -1.00, -72.00)     Camera orientation: 7.4, 1.6, 287.0 ``` |
| DL50N Yaw: 10.0° Roll: 0.0°   ``` Scenario: #8 (resolved) Subject: #3 DL50N Yaw: 10.0° Roll: 0.0° Tube lift: -6.229 mm Jaw contact X: 18.749 mm Jaw contact Y: 99.56 mm Tube position: (13.77, 107.59, -41.95) Tube rotation: (49.82, 10.00, 0.00) Upper incisor: (0.00, 108.00, -82.00) Lower incisor: (0.00, 97.00, -66.00) Glottic contact: (-3.00, -6.23, -0.12) Preglottic contact: (-2.06, -3.45, -10.00) Prepreglottic contact: (-0.08, 2.83, -20.00) Maxillary contact: (18.75, 99.56, -82.64) Blade contact: (8.59, 51.22, -57.81) Vallecula: (--, 25.00, -33.00) Jaw width: 65.4mm Resolve speed: 0.01 Cycles: 97 Cycles at speed: 3 Auto: On Ghosts: Off Cache hit: No Results: 56 Camera position: (122.84, -6.51, 185.88) Camera target: (-3.00, -1.00, -72.00) Camera orientation: 7.4, 1.6, 287.0 ``` | intubation view   ```     Camera position: (20.99, 345.12, -201.12)     Camera target: (0.00, 0.00, 0.00)     Camera orientation: -7.8, 0.5, 400.0 ``` | side view   ```     Camera position: (305.62, 61.30, -59.83)     Camera target: (8.00, 94.00, -41.00)     Camera orientation: 6.2, 1.7, 300.0 ``` | top view   ```     Camera position: (11.26, 6.17, -377.84)     Camera target: (0.00, 110.00, 30.00)     Camera orientation: 4.7, 1.8, 421.0 ``` | above view   ```     Camera position: (-3.01, 315.98, -75.17)     Camera target: (-3.00, -1.00, -72.00)     Camera orientation: 4.7, 0.0, 317.0 ``` | glottis view   ```     Camera position: (122.84, -6.51, 185.88)     Camera target: (-3.00, -1.00, -72.00)     Camera orientation: 7.4, 1.6, 287.0 ``` |
| DL50N Yaw: 10.0° Roll: 15.0°   ``` Scenario: #9 (resolved) Subject: #3 DL50N Yaw: 10.0° Roll: 15.0° Tube lift: -6.631 mm Jaw contact X: 21.389 mm Jaw contact Y: 96.646 mm Tube position: (30.19, 103.58, -42.28) Tube rotation: (49.48, 10.00, 15.00) Upper incisor: (0.00, 108.00, -82.00) Lower incisor: (0.00, 97.00, -66.00) Glottic contact: (-2.99, -6.63, -0.12) Preglottic contact: (-2.59, -3.77, -10.00) Prepreglottic contact: (-1.53, 2.61, -20.00) Maxillary contact: (21.39, 96.65, -83.02) Blade contact: (8.06, 52.99, -58.95) Vallecula: (--, 25.00, -33.00) Jaw width: 65.4mm Resolve speed: 0.01 Cycles: 89 Cycles at speed: 1 Auto: On Ghosts: Off Cache hit: No Results: 57 Camera position: (122.84, -6.51, 185.88) Camera target: (-3.00, -1.00, -72.00) Camera orientation: 7.4, 1.6, 287.0 ``` | intubation view   ```     Camera position: (20.99, 345.12, -201.12)     Camera target: (0.00, 0.00, 0.00)     Camera orientation: -7.8, 0.5, 400.0 ``` | side view   ```     Camera position: (305.62, 61.30, -59.83)     Camera target: (8.00, 94.00, -41.00)     Camera orientation: 6.2, 1.7, 300.0 ``` | top view   ```     Camera position: (11.26, 6.17, -377.84)     Camera target: (0.00, 110.00, 30.00)     Camera orientation: 4.7, 1.8, 421.0 ``` | above view   ```     Camera position: (-3.01, 315.98, -75.17)     Camera target: (-3.00, -1.00, -72.00)     Camera orientation: 4.7, 0.0, 317.0 ``` | glottis view   ```     Camera position: (122.84, -6.51, 185.88)     Camera target: (-3.00, -1.00, -72.00)     Camera orientation: 7.4, 1.6, 287.0 ``` |
| DL50N Yaw: 10.0° Roll: 30.0°   ``` Scenario: #10 (resolved) Subject: #3 DL50N Yaw: 10.0° Roll: 30.0° Tube lift: -7.969 mm Jaw contact X: 23.642 mm Jaw contact Y: 93.314 mm Tube position: (45.37, 96.13, -42.89) Tube rotation: (49.45, 10.00, 30.00) Upper incisor: (0.00, 108.00, -82.00) Lower incisor: (0.00, 97.00, -66.00) Glottic contact: (-2.92, -7.97, -0.12) Preglottic contact: (-3.00, -4.86, -10.00) Prepreglottic contact: (-2.77, 2.00, -20.00) Maxillary contact: (23.64, 93.31, -83.43) Blade contact: (7.22, 53.19, -58.96) Vallecula: (--, 25.00, -33.00) Jaw width: 65.4mm Resolve speed: 0.01 Cycles: 90 Cycles at speed: 3 Auto: On Ghosts: Off Cache hit: No Results: 58 Camera position: (122.84, -6.51, 185.88) Camera target: (-3.00, -1.00, -72.00) Camera orientation: 7.4, 1.6, 287.0 ``` | intubation view   ```     Camera position: (20.99, 345.12, -201.12)     Camera target: (0.00, 0.00, 0.00)     Camera orientation: -7.8, 0.5, 400.0 ``` | side view   ```     Camera position: (305.62, 61.30, -59.83)     Camera target: (8.00, 94.00, -41.00)     Camera orientation: 6.2, 1.7, 300.0 ``` | top view   ```     Camera position: (11.26, 6.17, -377.84)     Camera target: (0.00, 110.00, 30.00)     Camera orientation: 4.7, 1.8, 421.0 ``` | above view   ```     Camera position: (-3.01, 315.98, -75.17)     Camera target: (-3.00, -1.00, -72.00)     Camera orientation: 4.7, 0.0, 317.0 ``` | glottis view   ```     Camera position: (122.84, -6.51, 185.88)     Camera target: (-3.00, -1.00, -72.00)     Camera orientation: 7.4, 1.6, 287.0 ``` |
| DL50N Yaw: 10.0° Roll: 45.0°   ``` Scenario: #11 (resolved) Subject: #3 DL50N Yaw: 10.0° Roll: 45.0° Tube lift: -10.397 mm Jaw contact X: 26.032 mm Jaw contact Y: 88.746 mm Tube position: (58.67, 85.44, -44.14) Tube rotation: (49.57, 10.00, 45.00) Upper incisor: (0.00, 108.00, -82.00) Lower incisor: (0.00, 97.00, -66.00) Glottic contact: (-1.96, -10.40, -0.12) Preglottic contact: (-2.49, -6.86, -10.00) Prepreglottic contact: (-2.99, 0.83, -20.00) Maxillary contact: (26.03, 88.75, -83.99) Blade contact: (6.91, 52.22, -58.37) Vallecula: (--, 25.00, -33.00) Jaw width: 65.4mm Resolve speed: 0.01 Cycles: 96 Cycles at speed: 2 Auto: On Ghosts: Off Cache hit: No Results: 59 Camera position: (122.84, -6.51, 185.88) Camera target: (-3.00, -1.00, -72.00) Camera orientation: 7.4, 1.6, 287.0 ``` | intubation view   ```     Camera position: (20.99, 345.12, -201.12)     Camera target: (0.00, 0.00, 0.00)     Camera orientation: -7.8, 0.5, 400.0 ``` | side view   ```     Camera position: (305.62, 61.30, -59.83)     Camera target: (8.00, 94.00, -41.00)     Camera orientation: 6.2, 1.7, 300.0 ``` | top view   ```     Camera position: (11.26, 6.17, -377.84)     Camera target: (0.00, 110.00, 30.00)     Camera orientation: 4.7, 1.8, 421.0 ``` | above view   ```     Camera position: (-3.01, 315.98, -75.17)     Camera target: (-3.00, -1.00, -72.00)     Camera orientation: 4.7, 0.0, 317.0 ``` | glottis view   ```     Camera position: (122.84, -6.51, 185.88)     Camera target: (-3.00, -1.00, -72.00)     Camera orientation: 7.4, 1.6, 287.0 ``` |
| DL50N Yaw: 15.0° Roll: 0.0°   ``` Scenario: #12 (resolved) Subject: #3 DL50N Yaw: 15.0° Roll: 0.0° Tube lift: 1.982 mm Jaw contact X: 25.75 mm Jaw contact Y: 89.449 mm Tube position: (21.99, 113.52, -44.48) Tube rotation: (46.57, 15.00, 0.00) Upper incisor: (0.00, 108.00, -82.00) Lower incisor: (0.00, 97.00, -66.00) Glottic contact: (-3.00, 1.98, -0.12) Preglottic contact: (-1.62, 4.35, -10.00) Prepreglottic contact: (1.27, 9.77, -20.00) Maxillary contact: (25.75, 89.45, -83.89) Blade contact: (6.48, 25.02, -33.09) Vallecula: (--, 25.00, -33.00) Jaw width: 65.4mm Resolve speed: 0.01 Cycles: 90 Cycles at speed: 1 Auto: On Ghosts: Off Cache hit: No Results: 60 Camera position: (122.84, -6.51, 185.88) Camera target: (-3.00, -1.00, -72.00) Camera orientation: 7.4, 1.6, 287.0 ``` | intubation view   ```     Camera position: (20.99, 345.12, -201.12)     Camera target: (0.00, 0.00, 0.00)     Camera orientation: -7.8, 0.5, 400.0 ``` | side view   ```     Camera position: (305.62, 61.30, -59.83)     Camera target: (8.00, 94.00, -41.00)     Camera orientation: 6.2, 1.7, 300.0 ``` | top view   ```     Camera position: (11.26, 6.17, -377.84)     Camera target: (0.00, 110.00, 30.00)     Camera orientation: 4.7, 1.8, 421.0 ``` | above view   ```     Camera position: (-3.01, 315.98, -75.17)     Camera target: (-3.00, -1.00, -72.00)     Camera orientation: 4.7, 0.0, 317.0 ``` | glottis view   ```     Camera position: (122.84, -6.51, 185.88)     Camera target: (-3.00, -1.00, -72.00)     Camera orientation: 7.4, 1.6, 287.0 ``` |
| DL50N Yaw: 15.0° Roll: 15.0°   ``` Scenario: #13 (resolved) Subject: #3 DL50N Yaw: 15.0° Roll: 15.0° Tube lift: 1.876 mm Jaw contact X: 26.84 mm Jaw contact Y: 87.02 mm Tube position: (38.10, 108.75, -44.73) Tube rotation: (46.07, 15.00, 15.00) Upper incisor: (0.00, 108.00, -82.00) Lower incisor: (0.00, 97.00, -66.00) Glottic contact: (-3.00, 1.88, -0.12) Preglottic contact: (-2.14, 4.31, -10.00) Prepreglottic contact: (-0.16, 9.82, -20.00) Maxillary contact: (26.84, 87.02, -84.19) Blade contact: (5.47, 33.69, -42.06) Vallecula: (--, 25.00, -33.00) Jaw width: 65.4mm Resolve speed: 0.01 Cycles: 84 Cycles at speed: 1 Auto: On Ghosts: Off Cache hit: No Results: 61 Camera position: (122.84, -6.51, 185.88) Camera target: (-3.00, -1.00, -72.00) Camera orientation: 7.4, 1.6, 287.0 ``` | intubation view   ```     Camera position: (20.99, 345.12, -201.12)     Camera target: (0.00, 0.00, 0.00)     Camera orientation: -7.8, 0.5, 400.0 ``` | side view   ```     Camera position: (305.62, 61.30, -59.83)     Camera target: (8.00, 94.00, -41.00)     Camera orientation: 6.2, 1.7, 300.0 ``` | top view   ```     Camera position: (11.26, 6.17, -377.84)     Camera target: (0.00, 110.00, 30.00)     Camera orientation: 4.7, 1.8, 421.0 ``` | above view   ```     Camera position: (-3.01, 315.98, -75.17)     Camera target: (-3.00, -1.00, -72.00)     Camera orientation: 4.7, 0.0, 317.0 ``` | glottis view   ```     Camera position: (122.84, -6.51, 185.88)     Camera target: (-3.00, -1.00, -72.00)     Camera orientation: 7.4, 1.6, 287.0 ``` |
| DL50N Yaw: 15.0° Roll: 30.0°   ``` Scenario: #14 (resolved) Subject: #3 DL50N Yaw: 15.0° Roll: 30.0° Tube lift: 0.71 mm Jaw contact X: 27.813 mm Jaw contact Y: 84.489 mm Tube position: (52.95, 100.31, -45.33) Tube rotation: (45.85, 15.00, 30.00) Upper incisor: (0.00, 108.00, -82.00) Lower incisor: (0.00, 97.00, -66.00) Glottic contact: (-3.01, 0.71, -0.12) Preglottic contact: (-2.62, 3.38, -10.00) Prepreglottic contact: (-1.43, 9.31, -20.00) Maxillary contact: (27.81, 84.49, -84.50) Blade contact: (4.46, 36.90, -44.81) Vallecula: (--, 25.00, -33.00) Jaw width: 65.4mm Resolve speed: 0.01 Cycles: 93 Cycles at speed: 2 Auto: On Ghosts: Off Cache hit: No Results: 62 Camera position: (122.84, -6.51, 185.88) Camera target: (-3.00, -1.00, -72.00) Camera orientation: 7.4, 1.6, 287.0 ``` | intubation view   ```     Camera position: (20.99, 345.12, -201.12)     Camera target: (0.00, 0.00, 0.00)     Camera orientation: -7.8, 0.5, 400.0 ``` | side view   ```     Camera position: (305.62, 61.30, -59.83)     Camera target: (8.00, 94.00, -41.00)     Camera orientation: 6.2, 1.7, 300.0 ``` | top view   ```     Camera position: (11.26, 6.17, -377.84)     Camera target: (0.00, 110.00, 30.00)     Camera orientation: 4.7, 1.8, 421.0 ``` | above view   ```     Camera position: (-3.01, 315.98, -75.17)     Camera target: (-3.00, -1.00, -72.00)     Camera orientation: 4.7, 0.0, 317.0 ``` | glottis view   ```     Camera position: (122.84, -6.51, 185.88)     Camera target: (-3.00, -1.00, -72.00)     Camera orientation: 7.4, 1.6, 287.0 ``` |
| DL50N Yaw: 15.0° Roll: 45.0°   ``` Scenario: #15 (resolved) Subject: #3 DL50N Yaw: 15.0° Roll: 45.0° Tube lift: -1.804 mm Jaw contact X: 28.824 mm Jaw contact Y: 81.367 mm Tube position: (65.55, 88.33, -46.50) Tube rotation: (45.85, 15.00, 45.00) Upper incisor: (0.00, 108.00, -82.00) Lower incisor: (0.00, 97.00, -66.00) Glottic contact: (-2.97, -1.80, -0.12) Preglottic contact: (-3.00, 1.27, -10.00) Prepreglottic contact: (-2.48, 7.92, -20.00) Maxillary contact: (28.82, 81.37, -84.90) Blade contact: (4.39, 37.73, -45.62) Vallecula: (--, 25.00, -33.00) Jaw width: 65.4mm Resolve speed: 0.01 Cycles: 91 Cycles at speed: 2 Auto: On Ghosts: Off Cache hit: No Results: 63 Camera position: (122.84, -6.51, 185.88) Camera target: (-3.00, -1.00, -72.00) Camera orientation: 7.4, 1.6, 287.0 ``` | intubation view   ```     Camera position: (20.99, 345.12, -201.12)     Camera target: (0.00, 0.00, 0.00)     Camera orientation: -7.8, 0.5, 400.0 ``` | side view   ```     Camera position: (305.62, 61.30, -59.83)     Camera target: (8.00, 94.00, -41.00)     Camera orientation: 6.2, 1.7, 300.0 ``` | top view   ```     Camera position: (11.26, 6.17, -377.84)     Camera target: (0.00, 110.00, 30.00)     Camera orientation: 4.7, 1.8, 421.0 ``` | above view   ```     Camera position: (-3.01, 315.98, -75.17)     Camera target: (-3.00, -1.00, -72.00)     Camera orientation: 4.7, 0.0, 317.0 ``` | glottis view   ```     Camera position: (122.84, -6.51, 185.88)     Camera target: (-3.00, -1.00, -72.00)     Camera orientation: 7.4, 1.6, 287.0 ``` |
